# Challenges and opportunities in reforming the planning system for England: a rapid review of literature and lessons for the future of planning for health

**DOI:** 10.1101/2025.04.15.25325746

**Authors:** Michael Chang, Neil Carhart, Michael Cook, Charlotte Fox, Warren Lever, Lourdes Madigasekera-Elliott, Ellen Reith

## Abstract

Land use planning is primarily undertaken at the local level through local plans and development management decisions by local authorities. Planning includes objectives around promoting healthy and safe communities so public health considerations and involvement are necessary. A rapid review was undertaken of reports specific to the planning system between 2010 and 2024 and based on methods focused on gathering professional views. The reports were from academic papers, professional practice reports and parliamentary inquiries. There is a need to take a systems thinking approach to health focusing on the connections across the system, context and capability. This can help identify connected themes and gaps. In this context, the analysis found consistent themes for discussion: uncertainty in the reform process, clarity in national policy, complexity in plan-making and the local plan, delays in decision-making, politics in planning, reduced resourcing in local government, decreasing capability and skills, and concluding with reflections on improving public health consideration and involvement to drive local planning for health. The implications for public health involvement and consideration of health and wellbeing issues in planning as a key influence of the wider determinants of health can be profound with challenges and opportunities presented for future policy and practice.

## 1. Introduction

The planning system manages the use and development of land and is an important legal determinant of health-promoting policy development and decision-making at local levels (Gostin et al., 2019, Chang et al., 2022b). Changes or improvements to the planning system are usually introduced by national governments as a result of a combination of either changing ideologies or evidence about removing those barriers and challenges to achieving important priorities such as housing delivery (Shepherd et al., 2024). Delivering on the wider determinants of health through the built and natural environment must operate within the parameters of any planning system. Therefore, having a systematic understanding of the common but recurring challenges and opportunities that underpin any planning reform is important to ensure wider public health involvement consistently adds value and supports the planning process (Chang, 2017).

The planning system has been increasingly seen as a barrier to housing growth, competition and quality environments by UK Conservative governments. Such thinking and hypotheses have been influenced by policy think tanks such as the conservative Policy Exchange describing the ‘failed and failing’ system (Morton, 2011, Shepherd et al., 2024). The planning system was reforming following a change in government in 2010 from Labour to a Coalition Government of Conservative and Liberal Democrat parties. The Conservative Party 2010 election manifesto identified the planning system as a barrier to growth and committed to reforms such as creating a “*simple and consolidated national planning framework*” (The Conservative Party, 2010). The 2024 general election resulted in the formation of a new Labour government. Its election manifesto committed to an “*immediately update the National Policy Planning Framework to undo damaging Conservative changes*” (The Labour Party, 2024). The Prime Minister’s speech in December 2024 referred to planning as “*a blockage in our economy that is so big*” (Starmer, 2024). This signifies the planning system as a politically-charged lever to the extent that many planning commentators have suggested de-politicising planning to address society needs on housing and the environment (Bhalla and Payne, 2023).

One of the main reforms in the English planning system in the early 2010s as set out in the Coalition Agreement was the introduction of the National Planning Policy Framework (NPPF) in 2012 to replace the suite of Planning Policy Statements followed by the Planning Practice Guidance (PPG) in 2014. There have been several reviews of the planning system to examine and make recommendations to the government of the day on what changes might be helpful in ensuring the efficient and effective function of planning. These reviews have been supplemented by published academic and professional research examining planning barriers which this paper will seek to highlight. Planning commentators observe that since 2010, the English planning system has been in a constant reform and the implications have had and continue to have on both the operation of the system and outcomes it seeks to achieve (Raynsford Review, 2018, Sturzaker and Sykes, 2024).

Prior to 2010, under Labour governments, there were several reports that preclude this research. Notably these are the Barker Review of land use planning in 2006, Lyons inquiry into local government in 2007 and the Killian Pretty Review of the planning applications process in 2008. The Barker Review set out a number of recommendations to ensure that the planning system promotes a positive planning culture within the plan-led system, ensures sufficient resources for planning, enhances efficiencies in processing applications and improves skills and training (Barker, 2006). The Lyons Inquiry highlighted the need to reduce the complexity and detail of central control and local government’s place-shaping role through land use planning (Lyons, 2007). The Killian Pretty Review made recommendations to make the application planning process swifter, more efficient and more effective (The Killian Pretty Review, 2008).

At the same time, there has been an emergence and reunifying of effort to bring public health issues back into planning, especially given the early roots of planning in improving public health (McKinnon et al., 2020). During the initial period of planning reform of the Coalition Government in 2010, that was an initial window of opportunity. The Health and Social Care Act 2012 that saw health and social care reforms move public health functions from the NHS into local authorities, and also replacement of primary care trusts and strategic health authorities with clinical commissioning groups. Local authorities in upper tier and unitary areas are now responsible for planning, transport and public health as functions become integrated into a single administrative local government structure (Buck, 2020). Opportunities still exist in lower tier district authorities in utilising planning to shape to shape healthy places and impact the wider determinants of health (Local Government Association and Town & Country Planning Association, 2018, Local Government Association and District Councils Network, 2019).

Understanding of the reform process through systems and change management thinking can be particularly helpful. Systems thinking takes an integrative way to view a complex issue as part of a system of elements that function together as a whole. Taking a systems view in governance of public policy and complex issues such as planning system reform is not only necessary for understanding, but can also help identify points of leverage, facilitate successful decision-making, and help avoid unintended consequences (Meadows, 1999, Stroh, 2015, Bolton, 2022). Efforts have been taken to identify three perspectives – the system, context and capability, that have informed the analysis and discussion in this paper (Chang et al., Under review).

Change management is a complex process that results from identifying the motivational factors and problems of the current state that are influencing the case for change (Sewerin et al., 2022, John, 2018). An effective change management process requires different behaviours from those involved. In doing so, it can help us better plan for the degree of this change, and ultimately be successful in achieving reform objectives. We have made the conscious decision to frame the literature review in this way to highlight the range of recurring issues informing planning reform. The planning system and reforms to it cannot be seen from a singular issue given its multiplicity of inputs, throughputs and outputs. Without adopting as systems framing, we would not be able to fully comprehend and appreciate any connections between the issues.

The purpose of the study is to provide a systematic and efficient way to understand what the literature says about the case for change and reform to planning during a specified period of the 2010-2024 Conservative governments. The aim of conducting the literature review was to systematically identify the range of these reviews on the operation of the English planning system and summarise their findings on barriers, challenges and opportunities in an integrative way. It is the objective of the review to systematically identify relevant literature and understand their collective findings and recommendations informing any calls for change or improvement to the planning system. The outcomes will help us understand the relevance of these findings and to discuss lessons for future local planning for health in which public health involvement can add value, not burden, to the process. The main review questions were:

● What challenges and opportunities have been identified in relation to the operation of the planning system in England that have informed past and current programmes of planning reform?
● How are these issues relevant and appropriate to apply to improving our understanding of maximising consideration of health and wellbeing outcomes and enabling public health involvement in local planning?

## 2. Methods

A rapid review methodology was adopted with integrative review for analysis and discussion. A rapid review is a type of knowledge synthesis in which components of the systematic review process are simplified or omitted to produce information in a short period of time (Smela et al., 2023). This review type was preferred given its usefulness for policy assessment, flexibility to address the expected diversity of literature and timeliness while following the same methods as a systematic review. An integrative review provides a broader summary of the literature and includes findings from a range of research designs. It gathers and synthesises both empirical and theoretical evidence relevant to a clearly defined problem (Broome, 2000) and adapting the characteristics of a reflexive thematic analysis approach (Braun and Clarke, 2022) to find patterns within these reports and to present themes and patterns for others to inspect (Maykut and Morehouse, 1994).

Figure 1 outlines the process of undertaking the literature review including review team forming, search strategy, coding and analysis.

**Figure 1.**
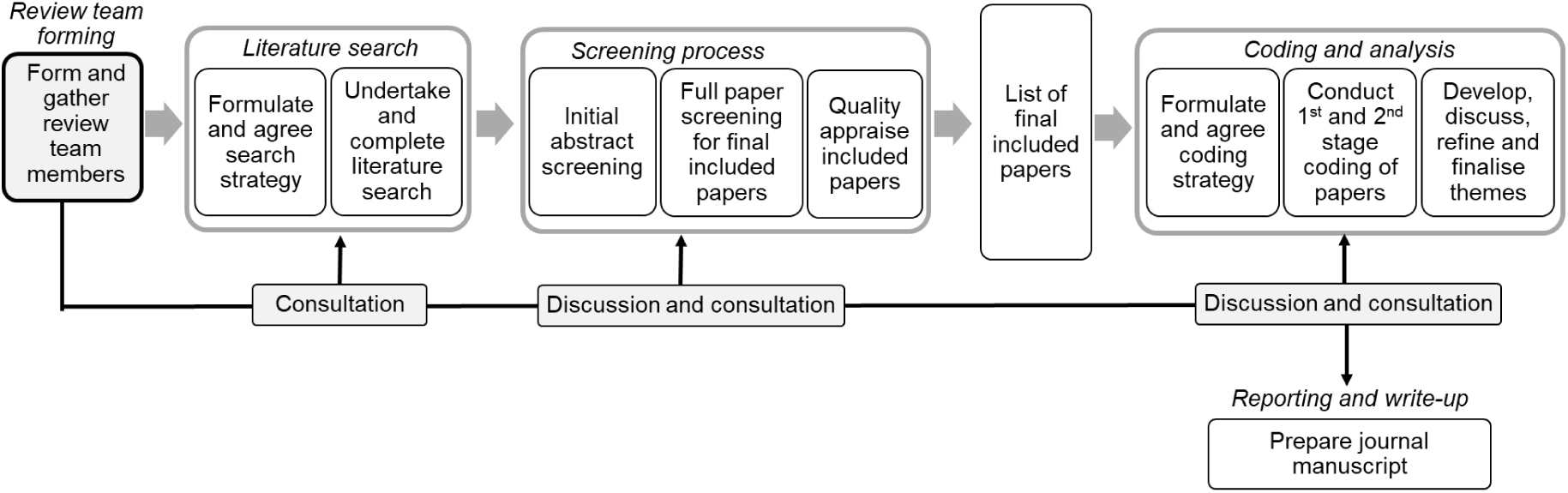
Literature review and analysis process.

### 2.1 Review team forming

A review team was formed to undertake the literature review. In addition to the main author, it consisted of four local government planning and public health professionals, a knowledge and evidence specialist from a national public health agency and an academic. The main tasks carried out by the review team members were literature search, screening and data extraction, and coding and analysis. Virtual meetings took place during key stages of the process to discuss issues and agree on decisions.

### 2.2 Literature search and screening process

Full details of the search strategy are available in supplementary material and a summary is described here. An initial scoping search was run on OVID Medline & Scopus (May 25, 2024) based on key terms (such as planning system review, planning reform, barriers and challenges) and key reports (such as from the Local Plans Expert Group) shared by the main author. Key terms were also identified from the titles, abstracts & data records of the scoping search result. Search terms were also identified and checked using the PubMed PubReMiner word frequency analysis tool. Given the exploratory nature of the scope a traditional PICO (Population, Intervention, Comparison, Outcome) strategy development was deemed not effective. Themes informing the initial search terms were informed from the research question: English planning system and issues that may impact its operation, and health/public health impacts. During the scoping phase, it was decided to remove the health/public health impacts as one of the primary search terms and use frequency operators around planning terms due to the ubiquitous and homographic nature of planning and planner within the evidence base. This approach was agreed at review stage by the review team.

The search was validated by testing whether it retrieved the shared key reports and 2 systematic reviews identified during the scoping phase. The reports indexed in OVID Medline were found and the non-Medline indexed reports were found within the Scopus strategy. The OVID Medline search strategy was peer reviewed by a UKHSA Knowledge & Evidence Specialist which involved proofreading the syntax and spelling used within the search and the overall structure but did not make use of the PRESS Checklist with agreed upon changes being made across all database search strategies. Grey Literature search strategies were based on the key free text terms used in the validated searches. Given the scope was focused on England’s planning system only, a validated UK only search filter was used on OVID Medline & OVID Embase with translated unvalidated versions used on Web of Science & Scopus. The search date range was limited to between 1 Apr 2010 and 31 Mar 2024.

Electronic searches for eligible results were conducted on the 21 Jul 2024 on the following databases: Ovid MEDLINE(R) ALL, Ovid Embase, Scopus, OVID Social Policy & Practice, and Web of Science. Additionally, a grey literature search of Google Scholar, Google, and TRIP Database Pro identified a further 378 results. Citation searching using Citationchaser identified an additional 306 unique results. All 6391 records were imported into Endnote Library. Deduplication was done using Dedudclick and Endnote. After de-duplication there were 5461 unique results.

Following de-duplication, records were imported into Rayyan Systematic Review software. After title and abstract screening according to a set of inclusion/ exclusion criteria agreed among the review team, 63 unique records remained, of which 61 were retrieved for full paper eligibility screening. A total of 46 reports was included in the final data extraction and analysis stage. Figure 2 summarises the outcomes of the search according to PRISMA.

**Figure 2.**
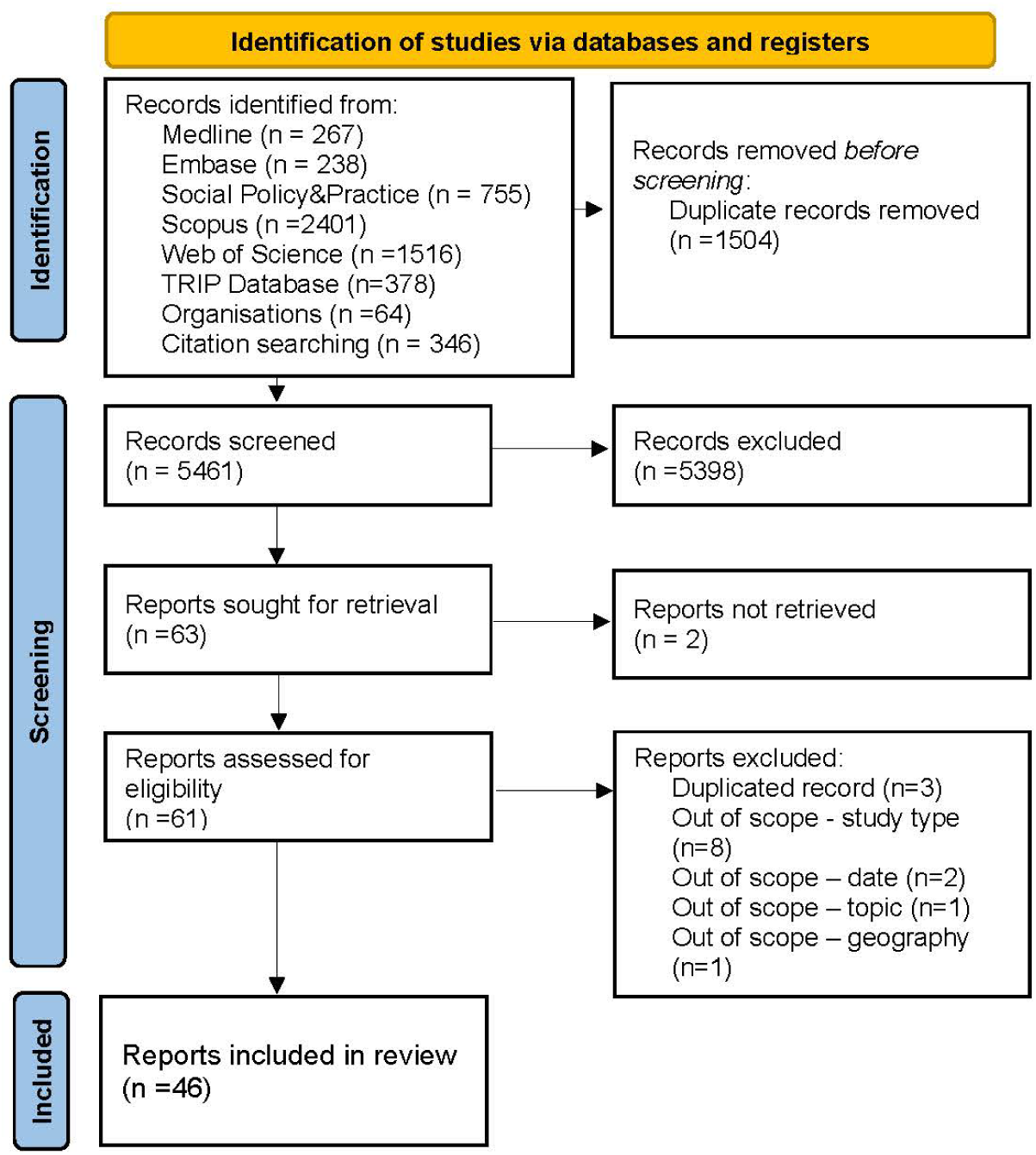
PRISMA diagram of inclusion/ exclusion reports.

Each paper was quality appraised using the CASP checklist for qualitative research (Critical Appraisal Skills Programme, 2018). The checklist had nine questions and responses to the questions had a total maximum possible score of 18. See supplementary materials for the table of scored attributed to each paper. We did not exclude reports on the basis of a low score, but the scores provided the review team with an indication of the quality of the research methods as set out in each paper.

### 2.3 Coding and analysis

Data was initially extracted from final included reports using a Microsoft Excel template provided by MC to members of the review team involved in this stage. The need for the creation and use of a template for the review team was in response to mitigate challenges associated with a greater number of collaborators and larger volumes of materials (Stemler, 2000, Krippendorff, 2013). Clear guidance and parameters during data extraction was needed when reviewing the diverse scope and type of reports eligible for inclusion. A template was created to help the review team to extract consistent data on:

- type of publication and study
- population on which research were conducted on
- research context, aims and objectives relating to the planning system
- findings, results, conclusions and recommendations
- any comments or observations

A coding strategy was developed and refined, and the coding process was undertaken through four separate cycles – first cycle of initial categorisation after familiarisation with the data, two cycles of coding and an intermediate cycle of refining the coding categorisation. (See Supplementary for the coding framework). The analysis process was undertaken to be consistent with six phases of the reflexive thematic analysis approach (Braun and Clarke, 2022).

- **Phase 1 Familiarisation:** Familiarising with the dataset was done as part of the screening and data extraction process during which the review team met virtually to discuss and agree approaches (June and July 2024). In addition to the data extraction activity, it involved MC tidying up the fulfilled template completed by different review team members into a singular Microsoft Word document in preparation for coding. As part of this process, initial ideas were noted and helped shape the coding strategy as a live and evolving document.
- **Phase 2 Coding:** This involved breaking down the data into ‘units of meaning’ (Maykut and Morehouse, 1994) and coding them into building blocks to inform theme generation. Code categorisation was informed *a priori* by a framework of ideas on planning for health systems thinking: the system, the context and capability (Chang et al., Under review). The first round of coding generated first-level codes under each node. Highlighted text and aligned codes were then extracted to a separate word document to complete the second round of coding which generated further distinct codes to aid understanding of the texts.
- **Phase 3 Theme generation:** Generating themes involved initially organising the first level and second level codes in phase 2 into potential categories.
- **Phase 4 Theme development:** Developing the themes involved consolidating and refining codes by grouping related codes and start to organise them according to the coding strategy for further the analysis.
- **Phase 5 Theme refinement:** Final naming of the themes was conducted and informed by the framework of codes in previous phases. This phase involved the review team meeting again virtually (October 2024).
- **Phase 6 Writing:** The writing up involved the final output in the form of this paper as well as the iterative documentation of comments and observations throughout both the coding and thematic analysis process.

### 2.4 Limitations

The diversity of initial literature presented issues for the eligibility screening process as they included study methodology (surveys, interviews and roundtables), publication type (academic, professional reports not published in academic papers and parliamentary or government commissioned reports) and publication sources (third sector organisations, governmental bodies and private consultancies). This made the literature research challenging to capture all the possible relevant reports. This challenge was recognised earlier on which informed the selection of the literature review approach and data extraction template. However, we acknowledged that there may be a number of reports that may not have been identified for example those not published and searchable in academic databases.

The use of qualitative analysis software was considered early in the process with an initial decision and preference for NVivo with its helpful functions to support analytical strategies (Kalpokas and Radivojevic, 2022). A technical hardware issue experienced during the process meant NVivo would not be available for use. As a result there was a period of reflection and awareness of literature precedent to set out the advantages and disadvantages of manual coding which included more engagement with the data and nuanced understanding needed (Isangula et al., 2024). There was a recognition of not accessing novel functionality available on NVivo through automated processes such as presentation of coded text and highlighting thematic relationships. Critics discussed the over-complication of such software for straight forward data organisation and structure in early career researchers (Ose, 2016). The impact of the problem on efficiency and quality was minimised by the coding process being undertaken only by the main author (MC) and the manageable quantity of included reports to code (n=46).

## 3. Results

A summary of the included reports and their key attributes are set out in Table 1. The main observations about the included reports are:

- 50% (n=23) of the reports are from academic sources, 39% (n=18) are from professional practice and 11% (n=5) from parliamentary sources. Reports are classified as academic if they were conducted and published in academic journals, professional practice if they were from professional and third sector organisations, and parliamentary if they were from governmental or parliamentary reports such as a select committee.
- Findings from the majority of the reports were obtained through interview methods (43%, n=20) followed by surveys or questionnaires (28%, n=13) and other mixed and non-defined methods such as parliamentary calls for evidence (26%, n=12). Documentary analysis of local plans (17%, n=8), practitioner engagement through workshops, roundtables or seminars (13%, n=6) and literature reviews (11%, n=8) were methods also employed.
- While the main literature search parameter was on the planning system, the reports covered a wider range of specific topics within the planning system. General planning for health was the most prominent (33%, n=15) followed by housing and planning (24%, n=11) and general planning (15%, n=7). Other topics that were covered included obesity, affordable housing, national and local planning, racial inequalities, sustainability and climate change.
- 9% (n=4) of reports were published during 2010-14, 37% (n=17) during 2015-2019) and 54% (n=25) during 2020-2024.
- Using the CASP checklist, the average quality score across the forty-six reports was 12.5 out of maximum score of 18. Most of the twelve reports that scored below the average score were ‘Professional Practice’ (n=7) while most of thirty reports above the average score were ‘Academic’ (n=20) although some ‘Professional Practice’ reports also scored above average (n=8).

**Table 1.**
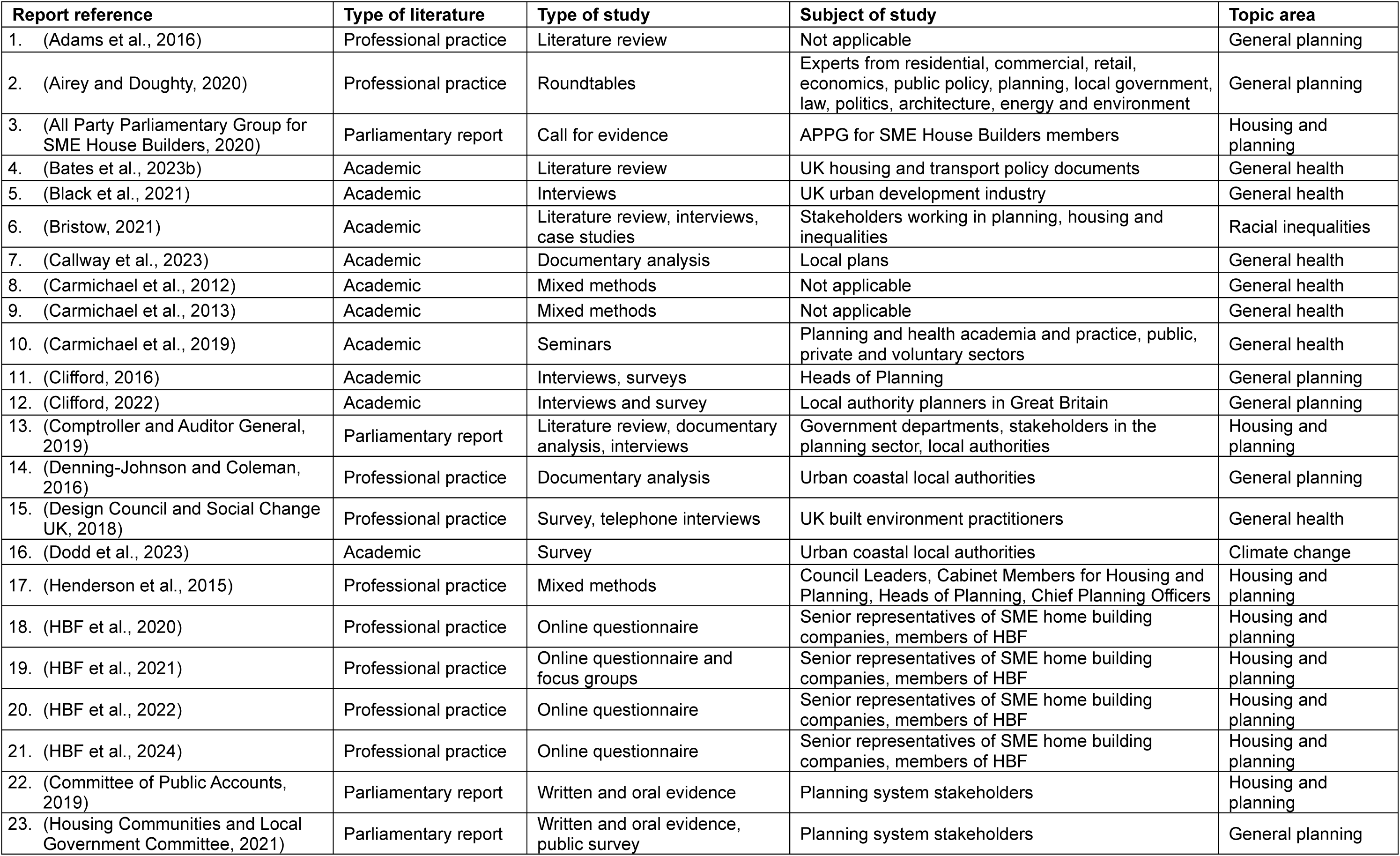
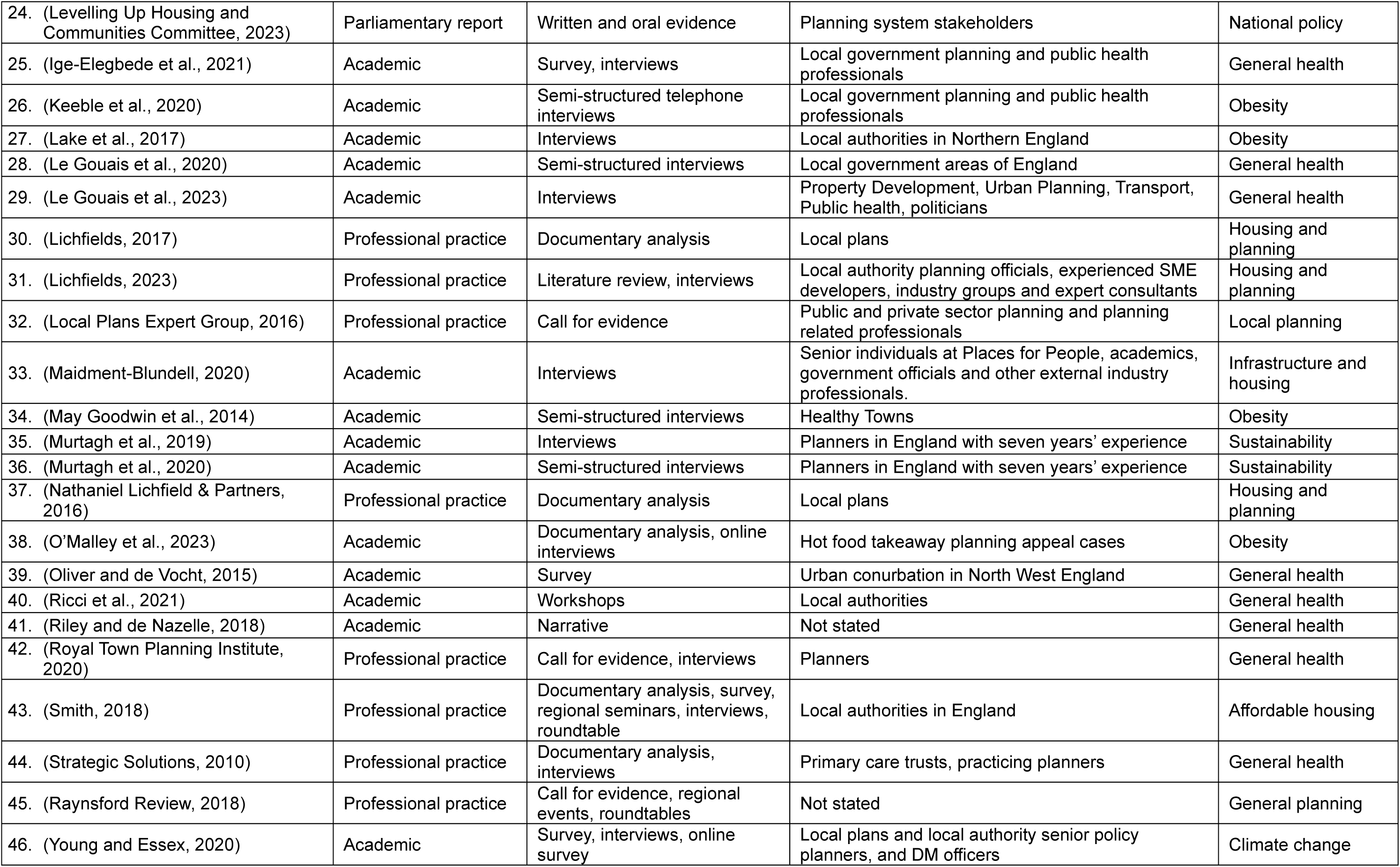
Summary description and literature characteristics of included reports.

## 4. Discussion of themes

The thematic analysis yielded important insight into the range of issues identified from the coding process. We observed issues that have been consistently highlighted throughout the literature published in academic papers, professional practice and parliamentary reports during the continuous period of planning reform programme.

From the systems perspective which framed the coding strategy and thematic analysis, we collated these issues into the following themes for discussion with numbered references to reports from the literature review in Table 1. While presented separately, the themes provided inter-connected and rich observations that reflect the complexity of the planning system.

- uncertainty in the reform process,
- clarity in national policy,
- complexity in plan-making and the local plan,
- delays in decision-making,
- politics in planning,
- resourcing in local government,
- capability and skills, and
- concluding with issues relating to improving public health consideration and involvement in the local planning system.

### 4.1 Uncertainty in the reform process

Parliamentarians voiced their concerns in 2023 by concluding “*stop-start reform over several years… has regrettably resulted in uncertainty among local authorities and across the planning sector*” (24, pp.9). Uncertainty had been a recurring keyword throughout the literature, appearing 139 times in 32 reports, and this was also evidence in planning commentaries such as from the Pegasus Group on the 2023 update of the NPPF that “*the Government originally planned to publish the update in Spring 2023 and the delays to its publication have added to the already significant uncertainty created*” (Hamilton-Foyn and Clarke, 2023). While many reports noted the need for the planning system to be reformed, the uncertainty experienced by the sector struggling with reform meant the “*cumulative impact on both policy and practice is a loss of confidence in planning for affordable housing*” (43, pp.45). After the initial reforms were put in place by the Coalition Government of 2010 with the new NPPF in 2012 and the new PPG, the Local Plans Expert Group was calling for “*a period of stability*” (32, pp.5).

Reports which sought widespread practitioner responses through surveys provided useful insight. For example, the majority of respondents to one such survey (43) suggested that specific change to the NPPF on viability testing of local policies have had impacted on councils not securing enough affordable housing. Proposed changes not yet implemented such as the introduction of a new Infrastructure Levy to replace Section 106 and Community Infrastructure Levy, can also have an effect with councils responding that the change would see affordable housing decrease or that they ‘don’t know’ in a 2022 survey (20). Four years after the Levy was first proposed in the 2020 Planning White Paper (Ministry of Housing Communities and Local Government, 2020) and one year after the technical details were consulted in 2023 (Department for Levelling Up Housing and Communities, 2023c), in August 2024 the new Labour Government confirmed it will not implement the new Levy mechanism (Ministry of Housing Communities & Local Government, 2024b).

Uncertainty and continuing changes to planning reforms have had a tangible impact on the day-to-day operation of the planning system with one paper from 2017, five years after the introduction of the NPPF, finding that “*over 60% of LPAs are still without a Local Plan tested and found sound*” (30, pp.2). In a 2023 parliamentary inquiry into the reform package, the select committee cited the reason many councils are delaying their local plans was that they were waiting for clarity and “*waiting to see the implications of the proposed NPPF changes for their local plans*” (24, pp.9).

Accepted practice in public policy making dictates some evaluation should or would have taken place so that politicians and policy makers can understand if and how a policy works for the public (Wond and Macaulay, 2010, Davies, 2012). It is not possible to find out whether successive governments have systematically undertaken and published standard evaluations that monitored and reviewed outcomes from the overall reform programme, with the exception of access to one evaluation of changes to permitted development rights for office to residential change of use (Clifford et al., 2020). In the UK context, this means following guidance in the Magenta Book on evaluation or the Green Book that states monitoring and evaluation of proposals should be included as an integral part of proposed interventions (HM Treasury, 2022, Evaluation Task Force, 2023).

One paper from a parliamentary inquiry noted their concerns “*about the lack of detail in respect of the proposed reforms to the planning system, which has made it very difficult to assess the possible practical implications of many of the reforms*” (Housing Communities and Local Government Committee, 2021). This follows a similar conclusion in 2018 that “*a period of near-continuous change in the planning system over the past decade has compounded rather than resolved the problems these so-called reforms were designed to remedy*” (45, pp.3). This effect can exacerbate unhelpful protection of professional interests as they make sense of the reforms rather than helping to achieve the wider aspirational objectives and integration intended by Government with one paper suggesting that “a*mbiguities in reform agendas open up spaces for institutional entrepreneurs to protect and further their own interests*” (12, pp.100).

The complexity of the planning system and any improvements to it will inevitably make it challenging to plan and predict (Phillips and Klein, 2023). While it takes time to implement and bed down change that may or may not be justified, it does not take away the temporal disruption from uncertainty and their compounding effect on clarity in national and local policy and practice.

### 4.2 Clarity in national policy

Successive governments have made and proposed improvements to simplify and streamline the planning system. The complete transformation and reconfiguration of national policy and guidance was one notable achievement. Prior to the first NPPF published in March 2012, national planning policies were articulated in a series of planning policy statements. The 2012 NPPF and the supporting PPG, a web-based guidance launched two years later in March 2014, replaced 7,000 plus pages of government guidance (Taylor, 2012).

But the effect of replacing an established complex web of national policy and guidance may have had the opposite effect and exacerbated uncertainty in the planning system. This was highlighted in 2016 by the Local Plans Expert Group stating, “*additional policy statements, new legislation and rapid changes to the NPPG, all of which may be beneficial in their own right but which do have the unintended consequences of destabilising plan making*” (32, pp.4). There have also been legal cases brought about to clarify the legal relationship between the NPPF and PPG for example in R (Mead and Redrow) v SoS LUHC [2024] EWHC 279 (Admin) on flood risk policy.

Although the NPPF is not named in planning legislation or constitutes a statutory instrument, it is an important legal determinant of land use policy. In its introduction, it states the NPPF must be taken into account in preparing development plans and is a material consideration in planning decisions (Department for Levelling Up Housing and Communities, 2023b). The application of the NPPF to support plan-making and inform planning decisions can only be achieved through greater specificity, consistency and intent which is in effect avoiding uncertainty. But in a parliamentary inquiry into the draft NPPF concluded it “*does not achieve clarity by its brevity; critical wording has been lost and what remains is often unhelpfully vague. its drafting must be more precise and consistent, and sufficiently to enable local authorities to write their own Local Plans*” (House of Commons Communities and Local Government Committee, 2011). One paper suggested that including tackling racial inequalities and meeting the housing needs of Black, Asian and Minority Ethnic (BAME) groups as a core NPPF purpose “*would compel local authorities to better address these aims in every aspect of decision-making*” (6, pp.51). Positively, another paper in relation to climate change noted “*initiation of such policies was driven largely by central government guidance, namely, the NPPF and NPPG (National Planning Policy Guidance). The response suggests that a strong top-down influence drives climate change policy formulation in plan making*” (46, pp.923-924).

The literature highlighted examples in certain policy areas of the importance of ensuring clarity and specificity in the NPPF. For example, practitioners commented that absence of “*social rent from the definition of affordable housing in the draft revised NPPF was particularly significant for the outcomes that could be secured from the planning system for those in greatest housing need*” (43, pp.28). The definition of affordable housing in the 2018 NPPF was ultimately included in the final published version. Another example illustrated by literature related to the imposition of national permitted development rights (PDR) for change of use from office to residential in 2013 which delivered the majority (87%) of the 102,830 new homes built since 2015/16 (Rankl, 2024). This PDR was highlighted as the cause of compromising local housing quality and ability of local authorities in securing affordable housing (14, 22, 43).

Too much specificity can have the opposite effect with one paper noting that “*planning regulations were perceived by some.to be inflexible*” and hinder consideration of key outcomes (9, pp.263). The literature also highlighted that “*a consistent approach is needed and necessary across all LPAs and this should continue to be provided by National Planning Policy Framework*” (3, pp.21). This juxtaposition of local plan policies and decisions in conformity to national policy is proving a challenge to make planning reform satisfy all users.

### 4.3 Complexity in plan-making and the local plan

There have been changes to the local plan in the years preceding the timeframe of this literature review. Most notably, this included Planning Policy Statement 12 on local spatial planning in 2008 stating every LPA should produce a ‘core strategy’ as part of a local development framework of documents aimed to break up and drive efficiency in developing the suite of documents. Substantial shift in plan production in the 2012 NPPF stated every local planning authority should have one ‘local plan’ document. Planning researchers have commented on the evolving nature of planning from land use to spatial planning and the need for planners to diversify knowledge and navigate increasing complexity in the process of creating local policies (Ferm, 2018, Parker and Street, 2021b).

Local planning is the main pillar of the planning system where strategic directions are set over a minimum of a 15-year period and policy requirements such as housing, environment and socio-economic outcomes are articulated to inform land use decisions in the community. Much of the changes to local planning have been framed and driven by a focus on housing delivery and the changing methodology for calculating housing needs which continues to be the source of dispute. This is supported by one industry explanation of increasing delays to plan-making that following examination in public scrutiny, “*there is often a necessary ‘patching-up’ of evidence which results in different housing requirement figures from that originally proposed*” (30, pp.5). The increasing scope and complexity of local plan evidence requirements was cited as a challenge when justifying policies during the examination in public process as well as part of the planning applications process.

With explicit reference to how housing needs are calculated and tested in relation to local environmental constraints to which the Local Plans Expert Group (LPEG) highlighted “*which have become one of the most burdensome, complex and controversial components of plan making*” (32, pp.2). The LEPG also highlighted local plan evidence complexities but equally framed them as burdens and unnecessary namely Sustainability Appraisal and soundness requirements that was corroborated by other reports i.e. in Lichfields (29).

Notwithstanding the unintended impact on local planning highlighted in Section 4.2 of this paper, the focus on striving to perfect process improvements in local planning had been a constant theme highlighted in literature. In reflecting stakeholder feedback, a parliamentary report highlighted “*there were allegations of a fixation with process, and widespread complaints that the system was too complex, obscure and slow*” (23, pp.11). There were genuine concerns highlighted across the sector about the speed of the plan preparation process and that “*it is necessary to use one of several “carrots and sticks” to incentivise local authorities to generate timely local plans or local plan reviews*” (32, pp.4). Similar concerns about the local plan-making process have been identified in the earlier reforms brought about in 2004 by the Planning and Compulsory Purchase Act (Shaw and Lord, 2009).

The 2020 Planning for the Future White Paper proposed measures intended to clarify a new role for Local Plans and a new process for simplifying their contents as it recognised that a poor planning process leads to poor outcomes (Ministry of Housing Communities and Local Government, 2020). While there is recognition that complexity in local plan contents was inevitable given the need to reconcile competing local priorities, others suggested that “*a looser binary zonal system should be introduced instead*” (2, pp.71). The White Paper outlines the introduction of new National Development Management Policies (NDMP) to make local plan contents faster to produce and avoid repetition. As the NDMP has not yet been created at the time of writing, the impact is yet to be made on local planning. There was “*concern at the lack of detail in the Bill, including on NDMPs. and this had also led stakeholders to hypothesise as to what might be enacted rather than respond to firm policy proposals*” (24, pp.32). The White Paper was an important milestone in planning reform at the start of 2020s as it influenced the Levelling-up and Regeneration Act 2023 but many changes to local planning are still yet to be introduced or bedded down. In a plan-led system, having an up to date local plan is often used as the litmus test of quality of the local authority planning function, and ultimately justification for state intervention (Ministry of Housing Communities and Local Government, 2020).

### 4.4 Delays in decision-making

At the sharp end of the planning system, decisions are made through the planning applications process or development management based on the local plan and other material considerations. The quality and speed of decisions and greater standardisation of information requirements were used in making the case for the current planning reform (Ministry of Housing Communities and Local Government, 2020). Previously, a 2008 review of the planning applications process by Joanna Killian and David Pretty had already identified five areas of concern from stakeholder feedback which were proportionality, process, engagement, culture and complexity (The Killian Pretty Review, 2008). A similar study focused on Wales for the Welsh Government was published in 2010.

The planning system for England operates a discretionary system which leaves room for interpreting policy requirements unlike a zonal system such as in the United States, Germany and New Zealand where decisions are made in accordance with detailed criteria. The differences between discretionary and zoning systems and their strengths and weaknesses have been set out by a Royal Town Planning Institute research paper (Schulze Bäing and Webb, 2020). One paper concluded that while “*the unusual level of flexibility. creates room to negotiate how proposed development might be improved. Equality, however, this discretion is often viewed as a cause of uncertainty by both developers and communities*” (1, pp.52).

The literature from the development industry have consistently identified decision-making in planning as a barrier. In a series of annual surveys of small housebuilders there was a marked increase in response rates to the causes of delays in obtaining planning permission. In 2020, 83% of survey responded points to “*delays in securing planning permission or discharging of planning conditions by local authorities as a major barrier to delivery*” (18, pp.5). In 2021, the figure increased to 94% in 2021 (19, pp.8), 93% in 2022 (20, pp.10) and again in 2023/24 (21, pp. pp.8). In the introduction reporting on the latest survey, it highlighted “*the consistency of the results, particularly across the past three years of the survey, points to a lack of progress made by Government and LPAs in tackling these issues*” (21, pp.9). The effect of delay was compounded by “*developers are also having to absorb increases in the costs of achieving an implementable planning*” (21, pp. 11) and cost of building homes increasing as “*any additional costs arising from higher requirements in the building regulations will likely increase construction costs*” (3, pp.19).

With the complexity of the planning application process, the literature had sought to have a better understanding of those factors underpinning the specific challenges and barriers experienced by users of the planning system or in achieving planning objectives. One factor was the amount of information requirements needed by LPAs to validate and determine a planning application. Such requirements are set out nationally by Government in regulations, for example design and access statement, and locally in accordance with NPPF criteria by LPAs to help meet certain policy or statutory responsibilities, for example air quality assessment, health impact assessment or daylight access (Department for Levelling Up Housing and Communities, 2023b). Notwithstanding changes that were implemented following the Killian Pretty Review recommendations, depending on the perspective, having requirements for certain assessments to accompany a planning application can be seen to be necessary to secure both quality of application and the development proposal. Requirements can also be seen as bureaucracy, adding to costs and it was suggested that one of the top three actions for supporting SMEs in the planning process was it “*specifically remove red tape*” (19, pp.21) to which the planning profession responded that “*to enable us to build more homes and better places, we need to broaden the focus beyond. less red tape*” (1, pp.6). Planning researchers have suggested the lasting impact of such policy dismantling as opposed to policy making in local government responsibilities (Eckersley and Tobin, 2019).

Another factor was the requirement for consulting or involving other users of the planning system such as the Planning Inspectorate (PINS) and statutory consultees. On PINS, it was highlighted in the evidence received in a parliamentary inquiry that they “*had become more risk averse, for instance through demanding more documentation and rejecting more planning proposals at appeal*” (23, pp.12). But others including the Raynsford Review expressed the opposite concern of a “*system no longer functioning as a positive public interest framework for decision-making. The very high level of successful appeals during the last five years for major housing development is one signal*” (45, pp.51). It was stated that PINS had been slow in determining appeals but that it “*acknowledges its performance is unacceptable*” (13, pp.9). Further insight can be obtained by referring to a government-commissioned Rosewell inquiry into planning appeals in 2018 which was not included in this literature review (Rosewell, 2018).

Planning regulations specify the requirement to consult a list of 27 statutory consultees for applications of certain developments, for example consulting Sport England for developments affecting playing fields plus any non-statutory consultees that may have an interest. The PPG affords LPAs discretion to consider whether there are planning reasons to engage other consultees who have an interest, such as public and environmental health. Some literature supported the involvement of consultees that for example “*enables them to articulate the future infrastructure needs of the residents in new developments*” (33, pp.34) or “*to limit the potential for catastrophic infrastructure failure and consequential harm to people and property*” (23, pp.25). But others suggested a prominent characteristic that they “*were often very slow to engage with developers. This reflects a long-standing complaint*” (23, pp.25) or “*length of time which many statutory consultees took to respond during the plan making process*” (32, pp.37). In December 2023, former Secretary of State the Rt Hon Michael Gove MP, appointed Sam Richards, Chief Executive of campaign NGO – Britain Remade, to conduct a rapid review of statutory consultees. According to Sam Richards the report remained “*as yet unpublished but sat on the desk of the new Secretary of State*” (Richards, 2024) following the July 2024 election, adding to further uncertainty and lack of clarity in the system.

At the sharp end of the local planning system, any delays to decision-making risk reducing the ability of the system to deliver priorities such as housing and environmental protection, sustain public trust and secure private sector investment. This often adds to and resulting in the emergence of a tone in the discourse in planning that centres on politicians and politics.

### 4.5 Politics in local planning

The politicisation in and of the planning system has been subject to much discourse. Planning is beyond a technical activity but one that delivers society’s complex, often changing and competing, needs through creating policies and making land use decisions. At the same time, it operates paradoxically as a plan-led system in a market-oriented real estate economy (Allmendinger and Dunse, 2005, Adams and Tiesdell, 2010, Adams et al., 2016). It is inevitable the planning system attracts what planning researchers describe as interest-based planning, political oversight and political gaming (Parker and Street, 2021a).

Improvements to the planning system have sought ways to better mediate competing interests but ultimately the determination is a political one. One paper acknowledged the task of weighing up interests of various stakeholder groups but “*in reality decisions are often made on political grounds”* (41, pp.644). Others saw galvanising support to mitigate the impact of politicisation of competing issues such as climate change where “*what progress there has been. has come in response to public pressure to act*” (16, pp.12).

The role of local politicians and the political institution of the local authority is a key pillar in the planning process to democratically legitimise decisions made on developments on behalf of local communities and achieve the corporate mandate of the local authority. When leveraging positive politics to maximise value from the planning system, the literature noted that “*when they achieve the right level of political support, plans and strategies have the power to generate shared frames of understanding that can shape the subsequent expectations and behaviour of a wide range of actors*” (1, pp.49). Reports often associated “*councils that have a strong institutional and corporate commitment to securing social and affordable housing. have greater levels of success*” (43, pp.30) in delivering vision and outcome-led aspirations such as on affordable housing.

However, there were also opposing perspectives depending on political ideologies on certain policy issues such as on health with one paper suggesting “*local politicians sometimes considered planning policy adoption to be a “nanny-state” approach*” (26, pp.6). On climate change or sustainability, it was suggested that “*the issue was not a big enough political priority to motivate the elected members to “drive” policy formulation in that area*” (46, pp.925). Impact of loss of strategic planning on issues that cross administrative boundaries was highlighted where “*tension was perceived between neighbouring councils with different priorities, with suggestions that councillors may not care about the health impacts of urban development for residents living outside their administrative area*” (29, pp.5). With calls that “*there needed to be ‘better co-ordination between districts’ in some areas. of both planning for housing and the management of housing stock*” 17, pp.28).

Involvement of local politicians in the process was cited as a barrier and cause of delay. One paper reflected stakeholder views on plan-making that “*the absence of a Local Plan is clearly a major issue for housing delivery and appears to be related to both resources, political consensus*” (17, pp.28). Gathered views from housebuilders would often raise the impact of politics as a challenge and cause of delay or ultimately even refusal of planning permission. One paper highlighted cases where elected members often refuse without justification and suggested they “*are involved in too many planning applications. More efforts need to be made that (decisions) should be delegated to Officers*.” (3, pp.16).

The operation of the local planning system is necessarily reliant on local politicians as democratically accountable to local communities on planning decisions made. At the time of writing this paper, the Government is consulting on introducing mandatory training for elected members on planning committees and clearer requirements for delegation of decisions to officers (Ministry of Housing Communities & Local Government, 2024a). The consultation paper highlighted the insufficient understanding among planning committees of planning principles and law which can inhibit their ability to make decisions. Part of the problem and solution lies in the trend in local government resourcing of the planning department.

### 4.6 Resourcing in local government

The complexity and diversity of local authority planning functions in plan-making and decision-making on land use development includes managing complex negotiations, meeting statutory responsibilities and securing sustainable development. Carrying out these functions require funding for human resources against the backdrop of increasing population and meeting housing targets. Local government financing has tightened over the years with increasing inequality across and within councils (Gray and Barford, 2018). The Institute of Government calculated that there was a 15.8% reduction in spending on local authority planning services in 2022/23 since 2009/10 (Hoddinott et al., 2023) and the Royal Town Planning Institute (RTPI) estimated a loss of 1,000 planners between 2009 and 2018 (Patterson-Waterston et al., 2020).

The term ‘resourcing’ had been mentioned 574 times in the reports from our literature review. In 2016, the year in which resourcing was first mentioned by the RTPI with “*concern about the gradual residualisation of planning services and the profession within local government*” (1, pp.49) and by private consultancy about “*resource challenges planning departments are facing in getting Local Plans adopted*” (37, pp.15). In the same year in a seminal government-commissioned report on local plans was completed by the Local Plans Expert Group, the only reference to resourcing was not about local government but a recommendation to Government to undertake “*a review of PINs resources in the light of the full scale of recommendations set out in this Report*” (32, pp.49) to address capacity to examine many new local plans. Reduction in resourcing of other key users in the local planning process was also highlighted on one paper that stated the “*reduction in the capacity of local planning authorities and statutory consultees. has made it more difficult for LPA planners to make timely and proportionate decisions effectively*” (31, pp.21).

The reports were consistent in their identification and framing of challenges associated resourcing of local planning authorities. This was particularly visible in the Home Builders Federation surveys which saw 73% of respondents believe a lack of resource in local authority planning departments is a major barrier in 2020 rising to 90% of respondents in 2023/24 and overall, there was “*little change at the very top of the table in comparison with other years, with staff resourcing/ shortages being seen as the number one major constraint*” (21, pp.13).

Reports agreed about the impact of reduced capacity on efficiency of process and quality of outcomes. They illustrated these impacts that included inefficient use of resources where “*most LPAs, who now seem to be dependent upon planning consultants in lieu of permanent planning officers*” (3, pp.10), delays caused by staff turnover where “*applicants are often forced to consistently ‘start again’ on demonstrating the case for development, often in the face of inconsistent feedback on proposals*” (31, pp.20), implementation from “*frequently lack resources and capacity to ensure that effective proactive enforcement can be operationalised*” (10, pp.159), impact on knowledge retention such as “*erosion of knowledge relating to climate change in their team with impacts upon Local Plan development*” (46, pp.925), and even associating capacity shortage’s “*cumulative impact on both policy and practice is a loss of confidence in planning for affordable housing*” (43, pp.34).

To remedy the challenges, in July 2023, the previous Government published guidance and committed a £24 million Planning Skills Delivery Fund over the next 2 years to tackle backlogs in planning applications and boost capacity (Department for Levelling Up Housing and Communities, 2023a). Labour restated their election pledge in July 2024 to employ 300 new local planning officers (Reeves, 2024). There are 326 LPAs in England. Figure 3 indicates the years in which reports had substantive content discussing local government resourcing and suggests that the rise in the issue post 2017 could be a result of cumulative lack of action by successive governments until recently. This suggestion was consistent with our observation, beyond identifying the challenge, that reports did not recommend tangible and specific measures to counteract the challenge and improve on the status quo. We also note there have been several reports published in 2024 that have highlighted challenges under the resourcing and capability themes. They were not reflected because publication was outside of the literature search period.

**Figure 3.**
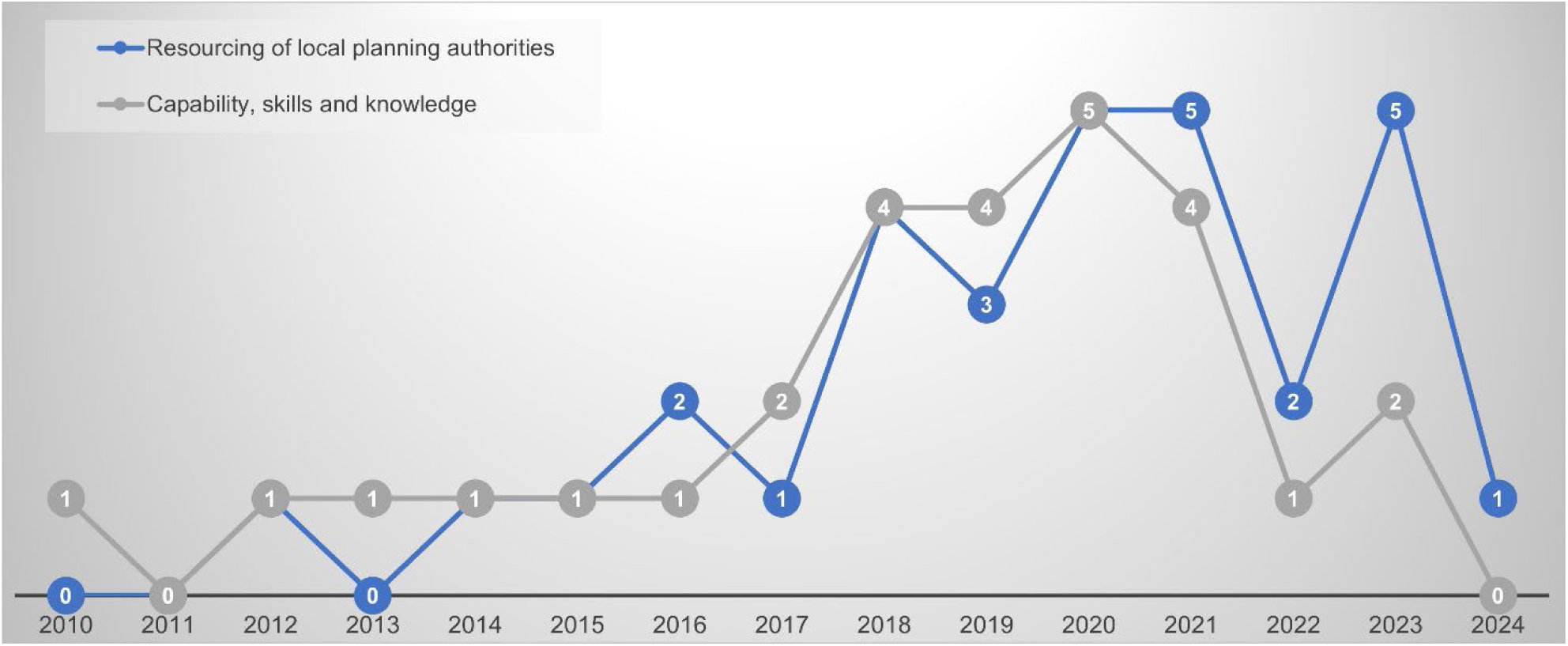
Timeline of when resourcing and capability issues were identified.

### 4.7 Capability and skills

While some of the literature highlights capacity and capability jointly and interchangeably, we have chosen to de-couple the two themes because they have distinctive defining impacts on planning in terms of quantity and quality. The term ‘skills’ had been mentioned 269 times in 32 reports in this literature review. Much of the issues raised here can be framed in terms of planning researchers citing the culture change was needed as part of understanding and implementing changes to planning system (Shaw and Lord, 2009). Figure 3 indicates the years in which reports had substantive content discussing skills and capability challenges, corresponding to the rise in interest in resourcing post 2016.

An increasing complex society with complex needs requires planners to acquire additional skills. The literature overall reflected contemporary concerns about the skills gap. Some of these skills relate to the renewed emphasis on design and the ability to commission or interpret technical assessments such as on housing or environmental. One paper expressed concerns from the sector “*about a ‘lack of resources’ and ‘specialist skills’ needed to undertake these assessments*” (17, pp.28). A parliamentary report noted the gradual loss of specialist staff in local government from 2006 such as on design, architecture, heritage and ecology, and concluded that “*Government’s reforms require an increase in planning staff, especially those with specific specialist skills, such as design. These skills gaps will need to be filled*” (23, pp.80). The report highlighted the RTPI’s evidence that part of the *“£500 million over four years was needed in additional funding. would be a specific design element*” (23, pp.79). The increasing use of digital technology and the digitalising of the planning process was highlighted as a reform solution, but the skills gap was also highlighted.

Delays in the planning system have also been attributed to the loss of experience and qualified capability. This attribution was consistently identified through engagement with the public and private sectors. The conclusion of a parliamentary report representing housebuilders suggested *“(lack of) experience and expertise are increasingly the reason for delays. Well qualified, quality planners would address this widespread blockage, which is evident across all Local Authorities*” (3, pp.21). From practitioner interviews, the Raynsford Review conceded a situation of arithmetic where “*some local authorities have fallen below a critical mass of both capacity and the skill to do the job in an effective and timely way*” (45, pp.61).

The literature had associated the capability and skills challenge with the increasing complexity of the planning in addressing socio-economic and environmental objectives beyond the traditional land use planning. One paper questioned whether “*planning processes are able to cope with the required level of experimentation, flexibility and iterative learning*” (10, pp.161). While another suggested that greater agency should be attributed to experienced planners where they are able to “*influence the very neoliberal reforms they are enacting might thus be the best guardian against the complete reduction of planning* (11, pp.398).

The above raises the question about the quality and responsiveness of the educational system to modern and future challenges. Many reports recommended solutions including a closer public-private collaboration with “*an academy-led model, run by a nationally recognised planning consultancy, would allow for a university course/module. to allow students access to the working world*” (3, pp.23). Other reports raised more fundamental concerns about recruitment and retention reflecting a survey by the Local Government Association reporting “*58% of all councils said that they were experiencing difficulties in recruiting planning officers, which rose to 83% of county councils*” (24, pp.24). The National Audit Office suggested while the Government was aware of the issue in 2017, it did not “*understand the extent of skills shortages in planning. the Department does not collate comprehensive data on the extent of this shortage*” (13, pp.11). The concern also extended to PINS.

In July 2023, the Government announced the development of a capacity and capability programme to support local planning authorities to attract, retain and develop skilled planners, as well as encouraging a new pipeline of planners to enter the profession (Department for Levelling Up Housing and Communities, 2023a). This included support for apprenticeships and postgraduate bursaries, and commissioning Public Practice to conduct nationwide recruitment and skills surveys in 2023 and 2024 which consistently identified the difficulty in attracting appropriately qualified or skilled candidates as the main issue (Public Practice, 2024).

### 4.8 Implications for public health in local planning

The challenges associated with the earlier themes have had and will continue to have consequences for local planning for health. The planning system, the local plan and its institutions constitute legal rules and frameworks that exert a powerful force on the social determinants of health, positively and negatively (Gostin et al., 2019).

Much of the literature was the result of efforts by researchers to better understand the dynamic evolution of the planning system since 2010 and the role of public health professionals and considerations within this process.

Planning reform during the early years of the 2010 Government coincided with the Health and Social Care Act 2012 reforms to the health and care system which moved public health responsibilities back into local authorities from NHS’s administrative bodies. Planning and health researchers in 2013 marked these health reforms as an opportunity and important milestone for strengthening the planning and health relationship in local government and noted “*it will be interesting, in time, to examine whether the new system offers better opportunities for planners to consider health outcomes of their decisions or if it will prevent progress being made*” (9, pp.264). The challenges remained across the two professions in the ensuing years. A parliamentary report in 2016 concluded the current planning process continues to be a major impediment to health (House of Commons Health Select Committee, 2016) followed by another paper in 2017 observing *“(directors of public health) DsPH also did not notice a major change in planners’ responsibilities as a result of the restructuring*” (27, pp.188), and another in 2021 reporting results from workshops conceded “*there is still a big disconnect between the worlds – public health commissioners are not equipped with the knowledge or language*” (40, pp.18).

Acknowledging the framing of politics and role of politicians in planning is a prerequisite for securing policy buy-in as planners have “*expressed their concern that.. the political nature of planning and the realities of day-to-day practice and pressure inherent in the system are not addressed*” (10, pp.157). Others expressed concerns about political support negatively where “*a lack of political support at the local level makes it difficult to influence local (health) policies*” (25, pp.667). Also, positively when local politicians “*provided impetus for Planning professionals to explore regulation*” (26, pp.6) especially where evidence exist to suggest a relationship between political majority and policies on the food environment (Keeble et al., 2019, Chang and Hobbs, 2024). Ultimately the literature was consistent in highlighting the need for “*very brave politicians to support healthy development*” (29, pp.4), and ultimately “*unless there is real political prioritisation of health outcomes among members, a council’s approach to health in spatial planning is likely to be little more than tokenism*” (44, pp.vii).

The politics of planning is played out when determining proposals when planners assess a proposal against the local plan and other material considerations. The discretionary nature of the English planning system has been highlighted as an opportunity for other issues such as health to be taken into considerations from traditional planning matters but that this discretionary “*is often viewed as a cause of uncertainty by both developers and communities*” (2, pp.52). Reports reflected practitioner views over the years for health on this uncertainty and inconsistency across decision making and called for health to be better embedded as a material consideration to “*remove these doubts and also enable evidence of health impact to be handled on the basis of reasonable probability rather than absolute proof*” (41, pp.viii). In the first update of the NPPF since 2012, the 2018 version revised paragraph 91 under the Promoting Healthy and Safe Communities chapter stated “Planning policies and decisions should aim to achieve healthy, inclusive and safe places” which in effect signals decisions to be made considering health.

Planning activities are informed by evidence about the economic, social and environmental characteristic of the area including those about public health needs and priorities. Planning reform has sought to streamline volume of evidence requirements and reduce their burden on applicant and planners. Aside from volume, the literature consistently raised the challenge about the relevance and usefulness of public health evidence. Evidence used to create land use policies usually comes in the form of site-specific assessments, and in the planning decision-making process, expert judgements are also highly weighted. In reflecting practitioner feedback, one paper noted “*local practitioners are often unclear about how they can apply evidence of intervention outcomes from academic studies”* (6, pp.157) and “*these types of evidence were useful for describing problems but did not provide solutions useful for decision actors*” (35, pp.114). Researchers noted the complexity of the system and healthy places which requires access to different information and such critical health evidence from the traditional evidence hierarchy may be under-represented (Tate, 2020, Chang et al., 2022b, Bates et al., 2023a).

The implications for public health involvement and consideration in the planning process are profound if there is a fundamental professional disconnect. Learning from a national programme of healthy new towns between 2016 and 2019 identified “*a learning curve that must be appreciated to aid understanding of different roles, the barriers that can exist, and how collaborative working can move forward to support healthy communities*” (34, pp.123). A contributing factor was due to “*impacts of reduced local authority budgets on the availability of resources and on the skillset needed to support collaborative work between public health and planning*” (25, pp.661). There was a suggestion in relation to the practical application of health data where there “*is also a need to build the capacity and skills of planners to effectively use and integrate the information derived from local JSNAs and other health data as well as the evidence from research and guidance*” (10, pp.158).

Skills and learning of planning and public health professions remained an issue in relation to education where “*it was also mentioned that the lack of health content in planning degrees*” (40, pp.18), and conversely “*there is a need for health practitioners to better understand the regulatory systems within which planners work. in order to have realistic expectations about how planning can impact on community health*” (25, pp.669).

The literature highlighted good practice despite the challenges experienced by local authorities where “*successful projects have been driven by planning professionals who initiated and developed collaborative, innovative and ‘integrated’ approaches to healthy placemaking*” (42, pp.34). A move towards strong inter-disciplinary and sectoral collaboration with “*academic-practice links and collaboration has also been emphasised in Bristol for instance to develop and implement the concept of healthy planning*” (10, pp.161). There was a notable increased recognition in the literature on the emergent benefits of different models of providing dedicated and specialist capacity for example “*the use of “broker” agency such as the London’s Healthy Urban Development Unit (HUDU)… dedicated officer with explicit health and planning responsibilities*” (9, pp.264). Capacity was found to “*break down silos and assist in building understanding, trust and rapport, therefore, encouraging collaboration. Such institutional changes can encourage the integration of health into decision making*” (41, pp.648). Such benefits have been verified in wider literature (Chang et al., 2022a, Coombes et al., 2024) with the government-commissioned 2014 Farrell Review of architecture and built environment going further to conclude embedding the knowledge and skills as early as possible in school education (The Farrell Review, 2014).

## 5. Conclusion and implications for policy and practice

The discourse around the planning system and agreeable ways to improve its operation have long been an area of interest by politicians, professionals, business interests, the general public, and researchers. As such it has also become an easy target for both criticism and ripe territory for political debate, research and professional reflection. At the same time, those with external interests of the planning system such as public health professionals, have continued to explore its potential to deliver wider societal priorities. The findings and discussion in this paper provide useful insight and avenues for future research, as well as contributing to learning lessons for policy and practice.

The themes we have selected for discussion suggested that there are systemic connections between them that require further unpicking. For example, the inability of local authorities to keep their local plans up to date despite various governments putting in place incentives and targets on the plan-making process had been attributed to a combination of uncertainty and reduced resourcing. But that reduced resourcing had been attributed to lower levels of decision-making on applications that have resulted in lower levels of housebuilding and therefore investment arising from growth and development. The pressure of the planning system to help address the increasing complexity of societal issues had contributed to lack of skills development in key policy areas including urban design, development economics and health, which in turn have affected decision-making. What is certain, however, is that a sense of stability and reduction in uncertainty is not likely in the short-term as the new government kicks of a wider programme of change. Therefore, practitioners need to learn to work flexibility and adapt to an uncertain environment.

In this paper, we have presented a selection of themes arising from our literature review, but it is clear from the analysis there are many more such as use of health in impact assessments and balancing economic considerations, which can help a more complete picture of understanding. Some themes exhibit a virtuous cycle of cause and effect. The literature review has been an important process of understanding the context and literature that have underpinned the discourse around planning reform over the years. A key observation from the literature was the challenge of associating their respective findings and recommendations to any subsequent implementation and follow-through of planning reform initiatives.

We can conclude there was a political undertone to the literature where government-commissioned and parliamentary reports that sought to identify reform measures to improve, modernise, simplify and speed up the planning system which has been a standing feature during the tenure of any administration. Most of the reports published during the Conservative governments between 2010-2024 have been included in this study. Such reports and desire to reform the planning system are particularly acute for incoming administrations with different political ideologies as evident with planning reform featuring prominently in the Labour Manifesto as one of the actions within 100 days in office (The Labour Party, 2024). There are several useful published analysis and commentary for further interest not least because we have excluded in our literature review, by design, reforms specific to regional planning abolition and the new neighbourhood planning system, see for examples (Tait and Inch, 2016, Riddell, 2020, Parker et al., 2023).

The issues raised are not novel but have been repetitive over the years, for example the Confederation of British Industry echoed views from professional practice reports included in this study published that the planning system is the biggest hurdle to housing delivery and investment (Confederation of British Industry, 2024).

Contemporary literature also increasingly highlights the relationship with building blocks ealth such as housing, access to greenspaces and active travel. For example in reports published as recent as the Radix Big Tent Housing Commission highlighted the health impact of not delivering more homes (Radix Big Tent, 2024). Another report published after the completion of our literature review by the Competition & Markets Authority highlighted the nature and operation of the planning system in relation to impact on housebuilding on people’s health outcomes (Competition and Markets Authority, 2024). Despite the challenges and threats presented in this paper, there were signs of positivity within local authorities to better use the planning system to achieve shared goals and priorities such as public health by valuing their professional judgement and supporting professional development (Murtagh et al., 2019).

However, these shared goals and priorities will not be achieved if challenges associated with the chronic resourcing, skills and capability in local government and the planning profession are not addressed. A survey of LPAs commissioned by the Ministry of Housing, Communities and Local Government in 2023 and published in early 2025 found that almost all planning departments reported planning skills gaps and relied heavily on external expertise to fill these gaps. Roles in public and environmental health featured high on the list on par with heritage and conservation and even higher than highways and transport (Verian and Ministry of Housing Communities & Local Government, 2025). With the planning profession increasingly recognised as part of the wider public health workforce such as by the World Health Organization and the Royal Society for Public Health, addressing the learning deficit of health in planning education has become a necessity (Chang et al., 2022a).

Aligning the planning and public health systems, especially in the wake of reforms is important because when asked how engaged the following stakeholders were in Health and Wellbeing Boards’ priority setting, planning emerged as the area within local government with the largest gap between current involvement and ambition for future involvement (Kuznetsova, 2012). Across health in local government responsibilities, the planning system had and continues to be a source of optimism as a net contributor to the new government’s prevention agenda (House of Commons Health and Social Care Committee, 2024). There is acknowledgement that planning and health reforms can help overcome some of the barriers to achieving healthy places but that planning for health will not happen by default (Ross and Chang, 2012, Chang et al., 2022b). Starting to address and connect some of the underlying issues in a systems approach can begin to shift the momentum from reactive to proactive local planning for health.

## 6. Author contributions

Relevant contributions have been attributed to individual authors in the submission version with authors details.

## 7. Declaration of interests

The authors declare that they have no known competing financial interests or personal relationships that could have appeared to influence the work reported in this paper.

## 8. Funding sources

This research did not receive any specific grant from funding agencies in the public, commercial, or not-for-profit sectors.

## Supporting information

Supplemental_Coded Text

Supplemental_Coding Framework

Supplemental_Search Strategy

Supplemental_CASP scores

## Data Availability

All data produced in the present study are available upon reasonable request to the authors.

## Notes

### Competing Interest Statement

The authors have declared no competing interest.

### Funding Statement

This study did not receive any funding.

