## Supplemental_Coded Text for "Challenges and opportunities in reforming the planning system for England: a rapid review of literature and lessons for the future of planning for health"

**Coding**

Notes:

- During the second coding process, I have identified First Level codes from the initial list to add to the extracted data, indicated in **Red** text.
- Second level codes formulated by deducting from the extracted data and therefore do not directly correspond or need to link to the First level codes.
- Second level codes provide further categorisation of understanding the data in relation to the First level codes
- The relationships and connections among the data, first and second level codes are being visualised separately in a network relationship diagram.

| **Data Page** | **Extracted data** | **First level code** | **Second level code (if needed)** |
| --- | --- | --- | --- |
| 2 | 2023 | Resourcing of local planning authorities | - |
| 2 | Governance was generally good, with local priorities reflected in policy and active encouragement of inward investment. However, all UCLPAAs were heavily constrained financially in economic development and regeneration functions, both in terms of investment capital and in terms of staff resources, which affected their ability to drive development. | Resourcing of local planning authorities | Impact on implementation |
| 2 | implementation being subject in all cases to externalities (either private investment or government funding opportunities) that were not aligned to local priorities | Complexity of planning regulatory framework | - External influence - Lack of alignment |
| 2 | under-resourcing of local authorities to properly resource plan implementation | Resourcing of local planning authorities | Impact on implementation |
| 2 | National planning policy for coastal tourism and leisure is a necessity | Strength of national policy | Clarity in national policy |
| 2 | The development of a standard methodology for analysing economic plan performance prior to the development of new policies is essential to allow local planning authorities to ensure that policies are delivering as expected, | - Use of impact assessment - Local plan content and plan-making process | - Impact on implementation - Frontloading/ upstream |
| 2 | Class E to residential permitted development rights should be repealed, and regulation of short-term holiday lets should be increased. | - Strength of national policy - Use of (local) planning powers | - Increase regulations - Repeal regulations (Permitted Development Rights) |
| 2 | The use of evaluative practice will allow for better plan-making, support governance | - Monitoring and evaluation - Local plan content and plan-making process | Governance |
| 2 | Greater emphasis should be placed on the use of local planning powers such as Local Development Orders | Use of (local) planning powers | Better of local levers (Local Development Orders) |
| 3 | 2016 | Capability, skills and knowledge | - |
| 3 | planners must have the confidence to focus more on how and why these will change spatial patterns of development from what might otherwise have occurred if markets had been left to their own devices, and less on the various statutory procedures that need to be signed off before such plans can be officially adopted. | - Visioning and outcomes-focused - Capability, skills and knowledge | - Confidence - Less focus on process - Frontloading/ upstream |
| 3 | National and local government need to consider the particular powers, resources and expertise | - Use of (local) planning powers - Capability, skills and knowledge |  |
| 3 | the value of planning needs to be analysed not on the basis of narrow measures of ‘efficiency’, but on the extent to which planning is delivering the economic, social and environmental value it so demonstrably can and how this might be maximised | Visioning and outcomes-focused | - Deliver wider value and benefits - Less focus on process |
| 3 | When they achieve the right level of political support, plans and strategies have the power to generate shared frames of understanding that can shape the subsequent expectations and behaviour of a wide range of actors across the public and private sectors and in civil society | - Impact of politics and politisation in planning - Visioning and outcomes-focused. | Impact on implementation |
| 3 | Generating local political agreement around the goals of a plan or strategy can be time-consuming and difficult. Planners, politicians, developers and the public have all regularly expressed frustration that the legal-bureaucratic complexity of aspects of the plan-making process makes the task even harder. | Complexity of planning regulatory framework | Impact on implementation  Political commitment |
| 3 | This has led the RTPI and others to express concern about the gradual residualisation of planning services and the profession within local government | - Complexity of planning regulatory framework - Resourcing of local planning authorities | Professionalism and impact on profession |
| 3 | Development control powers are reactive and rely on the discretion to determine whether proposals fit with a given local planning authority’s agreed definition of where the public interest lies in the use and development of land. The unusual level of flexibility created by the fact that “other material considerations” can outweigh the development plan allows local authorities to take into account a wide range of considerations, and to respond to new challenges and opportunities. It also creates room to negotiate how proposed development might be improved. Equally however, this discretion is often viewed as a cause of uncertainty by both developers and communities. | Decision-making process | - Uncertainty - Impact on implementation - External influences |
| 3 | Here the South Worcestershire case study is instructive. It is important however to begin to demonstrate the value of planning regulation in this way. The protection of key place qualities, for example, is often highly prized but not fully appreciated until it is threatened. In other ways too, a commitment to positive regulation can play a key role in ensuring high quality development. | Impact of economic focus, viability and costs | - Increase regulations - Deliver wider value and benefits |
| 4 | Regulation needs to be proportional and efficient, but it also needs to be re-valued for how effectively it can deliver the outcomes agreed in plans. All of this is easier to achieve in a context where there is a political commitment to resourcing regulation to deliver public value | - Impact of politics and politisation in planning - Visioning and outcomes-focused. - Resourcing of local planning authorities | - Increase regulations - Deliver wider value and benefits - Political commitment |
| 4 | The power to leverage private investment from public action: R12  An alternative mode of leverage involves using public sector land, assets or finance to draw private investment into areas where market-demand is low. A variety of regeneration initiatives in the 1990s and 2000s used public resources in this way to target support to deprived areas. In principle, public investment can help to reduce risk and increase incentives for private developers. | Stakeholders and statutory consultees | Role of private sector  Private sector investment |
| 4 | The power to leverage public value from private investment: A third and related means by which planning processes seek to deliver public value is by capturing a share of the value of private development to pay for key public infrastructure or services. This has become a vital means of ensuring delivery of the new infrastructure required to mitigate the impacts of development and to ensure that it is properly serviced.  An alternative mode of leverage involves using public sector land, assets or finance to draw private investment into areas where market-demand is low. A variety of regeneration initiatives in the 1990s and 2000s used public resources in this way to target support to deprived areas. In principle, public investment can help to reduce risk and increase incentives for private developers. There has however been some criticism of the terms under which such ‘partnership’ agreements have operated directly or indirectly to subsidise the private sector without offering guaranteed or meaningful returns on the levels of public investment involved. It is also important that potential unintended consequences of such intervention are acknowledged, including the displacement of existing uses and users if land values increase | Impact of economic focus, viability and costs | Private sector investment  Funding |
| 4 | Running through each of the powers described above is the argument that planners could deliver substantially better outcomes through a better understanding of the ways in which planning can and do shape markets | Visioning and outcomes-focused |  |
| 3 | Thinking about places first, by bringing together agencies, government bodies and service providers to identify and deliver the best long-term outcomes across different policy areas | - Visioning and outcomes-focused - Partnership, collaboration, relationships, engagement |  |
| 3 | In particular, the focus is on targeting resources to achieve the best long-term outcomes, rather than managing resources to minimise current spending. This focuses attention towards an investment approach, in which spending now yields long-term benefits, or at least minimises later problems from arising | Visioning and outcomes-focused | - Deliver wider value and benefits - Frontloading/ upstream - Funding |
| 3 | Making local plans and other strategies genuinely long-term visions, by using tools such as horizon scanning | Visioning and outcomes-focused |  |
| 3 | Learning the lessons from Urban Regeneration Companies, Urban Development Corporations, Enterprise Zones and other private-led partnerships, which demonstrate some of the ways in which barriers to development can be removed to facilitate development. | - Knowledge transfer, good practices - Stakeholders and statutory consultees | - Impact on implementation - Private sector investment |
| 3 | Opening-up large-scale developments to market forces within the context of appropriate masterplanning frameworks. On these sites, private developers could be given the freedom to deliver new homes | Stakeholders and statutory consultees | - Private sector investment - External influence |
| 3 | Building on the tradition of local community efforts to stimulate growth, manage development and create new settlements, through models such as Community Land Trusts | Stakeholders and statutory consultees | Role of local communities |
| 5 | 2020 | Resourcing of local planning authorities  Capability, skills and knowledge |  |
| 5 | Change the inefficient and costly approach of most LPAs, who now seem to be dependent upon planning consultants in lieu of permanent planning officers | Resourcing of local planning authorities | Role of private sector |
| 5 | the training of planners and other related professionals. Staff shortages, lack of people, experience and expertise are increasingly the reason for delays | - Resourcing of local planning authorities - Capability, skills and knowledge - Training, learning and development | Impact on implementation |
| 5 | Well qualified, quality planners would address this widespread blockage | Capability, skills and knowledge | - Professional qualification - Impact on implementation |
| 5 | More certainty and consistency in the planning system - SME housebuilders felt that in terms of the future of planning and improvement to be made, more certainty in the planning system both in terms of what the likely outcome will be and, how long it will take were the two key goals. A consistent approach is needed and necessary across all LPAs and this should continue to be provided by National Planning Policy Framework and guidance which is essential to the delivery of this consistency | - Impact of planning reform processes - Strength of national policy | - Uncertainty - Consistency - Clarity in national policy |
| 5 | SME housebuilders feel Design Reviews should take place at a very early stage in the process and not late on in the process where any benefit is likely to be small and the costs, in terms of delay, and associated costs of changing plans, road layouts, landscape and drainage details, as a result, for example, are very significant | - Use of (local) planning powers | - Better of local levers (Design Review) - Impact on implementation - Cost and delay - Frontloading/ upstream |
| 5 | Elected Members are involved in too many planning applications. More efforts need to be made that they should be delegated to Officers to determine in accordance with planning policy, whether that be national or local policies contained within the Local Plan | - Impact of politics and politisation in planning - Decision-making process | - Role of elected members - Impact on implementation |
| 5 | Currently, whilst determination timeframes and costs are lower in terms of application, the complexity involved in bringing through marginal sites and lack of planning balance to offset constraints and impacts means that determination is often slower, leading to delay to return and impact on cash flow | Decision-making process | - Impact on implementation - Cost and delay |
| 5 | Links between private consultancies and Local Authorities should be encouraged to help upskill junior planners via LPA internships | - Training, learning and development - Capability, skills and knowledge | Role of private sector |
| 5 | The Government needs to review and amend its Planning Guarantee (to determine applications within 26 weeks, with a further 26 weeks for an appeal if necessary), as it is rarely used as LPAs insist on applicants to agree an extension of time before the 13- or 26-week time period is up | Decision-making process | - Impact on implementation |
| 5 | Upfront fees to Local Authorities - Consideration was also given to the issue of upfront fees | Resourcing of local planning authorities | - Planning fees - Frontloading/ upstream |
| 5 | Training for Elected Members about local policy and development management should be meaningful | Training, learning and development | Role of elected members |
| 6 | New environmental initiatives, though often valid, also do come at an extra cost | Impact of economic focus, viability and costs | Cost and delay |
| 6 | There should be a legal duty on local authorities to pay special attention to tackling the housing supply issue, as is the case with the protection of listed buildings | Strength of national policy | - Increased regulations (housing) |
| 6 | Training for future planning professionals - Key to overturning current practices is to ensure that students are provided with high- quality, industry-led thinking that allows them to challenge the views of academics and gatekeepers and provides them with the skills to understand the requirements to look at planning via the three central tenets of the NPPF – Economic, Environmental and Social benefits | Training, learning and development | - Deliver wider value and benefits - Professional qualification |
| 6 | An academy-led model, run by a nationally-recognised planning consultancy, would allow for a university course/module, perhaps over one of the three years of study (the last would make sense), to allow students access to the working world, as well as the resource and expertise of some of the UK’s best and brightest on key topics and projects | - Training, learning and development - Capability, skills and knowledge | - Role of private sector - Externalised expertise |
| 7 | 2021 | Resourcing of local planning authorities  Capability, skills and knowledge |  |
| 7 | All bar one of the interviewees agreed that health is not being taken into account sufficiently when making decisions in urban development, and they would support the types of valuation mechanisms presented to them by UPSTREAM team: they (a) understood the uncertainties inherent in this type of approach, (b) were less interested in it as a tool for precise market cost-benefit appraisal and (c) more interested in it as an enabler for prioritising issues, understanding orders of magnitude and communicating value | Impact of economic focus, viability and costs | - Health consideration - Impact on implementation - Uncertainty - Deliver wider value and benefits - Innovative practices |
| 7 | In London and other major cities, there is also a substantial reliance on foreign investment, particularly from the USA and China, to provide longer-term financing. Despite the economic downturn since 2008, there was a broad sense that money is available as long as the conditions are otherwise right. A further challenge presented was that higher density development requires greater up-front investment, higher profit margins, and a cash flow model that is markedly different from plot-by-plot sales. The role of finance in provision of community infrastructure (e.g. public transport, local amenities and public realm) was raised repeatedly, and public sector investment was seen as crucial for the forward funding of infrastructure (e.g. paying up front for city tram lines, which won’t immediately recoup their losses), not least due to their ability to borrow more cheaply. It was also acknowledged however that local government is constrained in its lending, borrowing or otherwise take on risk, which shifts responsibility to the private sector and is more expensive . | Impact of economic focus, viability and costs | - External influence - Private sector investment |
| 8 | The theme of partnership was notable for two main reasons: firstly, for the level of importance given to it by interviewees (and how it underpins other core areas of risk, land and finance) and, secondly, for the multiplicity of characteristics and mechanisms that were suggested make up successful partnership: e.g. trust, shared values and vision, track record and parity of control. In other words, there was a clear lack of shared definition of what is meant by the term ‘partnership’ | Partnership, collaboration, relationships, engagement | - Risk management - Deliver wider value and benefits |
| 8 | On the role of the private sector, it was acknowledged that there is a wide range in quality between different private sector developers that the quality of the developer makes a significant difference to quality of environment and that partnering with higher quality developers was not always possible | Stakeholders and statutory consultees | - Role of private sector - Deliver wider value and benefits |
| 8 | Questions were raised about the relative merits of government intervention to support healthier development and for various reasons, not only its efficacy but also the (potentially significant) gap between policy and implementation | - Strength of national policy - Decision-making process - Monitoring and evaluation | Impact on implementation |
| 8 | A current and significant lack of resource and capacity within the public sector was clearly articulated in the interviews (i.e. capital budgets, number of staff, level of expertise), not just by the public sector interviewees (e.g. low salaries, lack of opportunity, lack of resource to create change) but also by the private sector (e.g. protracted negotiations, delays). | - Resourcing of local planning authorities - Capability, skills and knowledge | - Salary level - Cost and delay - Funding |
| 7 | 4. Balanced partnership: Establish what constitutes the optimal balance in partnership—between public, private and community—including determining the right balance (of power and resource) between parties. | - Partnership, collaboration, relationships, engagement |  |
| 7 | Evaluate and develop innovations in developer models, and what being a good partner means (factoring in: agenda setting and prioritization, trust, track record, shared vision, time horizons | - Knowledge transfer, good practices - Partnership, collaboration, relationships, engagement | Innovative practices |
| 7 | Investigate the role of national government policy, legislation and regulation in areas outside of planning policy (e.g. short-termism in finance, land control and disposal), and their potential impact on healthy urban development | Strength of national policy | Non-planning regulations |
| 9 | 2013 | Capability, skills and knowledge |  |
| 9 | As far as EIA is concerned, evidence demonstrates that the EIA process is generally effective in considering and assessing environmental health issues such as air quality, noise pollution, but other key health issues such as levels of physical activity, mental well-being and health equity are rarely considered. | Use of impact assessment | Environmental Impact Assessment  Health consideration |
| 9 | The fact that some authorities perform quite impressively in relation to health, while others do poorly as we will illustrate below highlights the important conclusion that it is not primarily the planning system which inhibits health-integrated plans | Local plan content and plan-making process | - External influence - Innovative practices |
| 9 | The attitudes, resources and knowledge of the key players and the rationale for the difference between the exemplary local authorities and others are not fully explained by the evidence available | - Capability, skills and knowledge - Stakeholders and statutory consultees | Professionalism and impact on profession |
| 9 | Planning regulations were perceived by some authors to be inflexible, and failing to highlight health in appraisal processes | - Complexity of planning regulatory framework - Use of impact assessment | Health consideration |
| 9 | Concerns were also raised about gaps in the quality and range of the local evidence base supposed to underpin the ‘soundness’ of plans and allowing planning permission, as well as inadequate scoping processes in plans, resulting in the exclusion of health and well-being as objectives | Evidence base | - Quality of evidence - Scope of evidence - Health consideration - Soundness - Frontloading/ upstream |
| 10 | Our literature review analysis suggests that those responsible for decisions on, and assessments of, planning proposals often view health in narrow terms, focussing on physical environment concerns such as air quality, rather than recognising the role of the social environment and other broader determinants of health. | Decision-making process | Health consideration |
| 10 | This narrow focus is seen to be primarily a result of a lack of engagement between health and planning professionals, coupled with the rigid boundaries around the development of knowledge between these two professions, different cultures between the various stakeholders, with differing terminologies and languages, priorities and structures (UWE, 2011c). Lack of understanding of the roles that different organisations and individuals hold is also factor in that respect (Tewdwr-Jones, 2011 | - Partnership, collaboration, relationships, engagement - Stakeholders and statutory consultees | - Lack of engagement - Professional qualification |
| 10 | Furthermore case study research in England shows that spatial planners have a weak knowledge of how they can influence the determinants of health (NICE, 2011). | Capability, skills and knowledge |  |
| 10 | A number of good practice examples have nevertheless emerged involving the use of “broker” agency such as the London’s Healthy Urban Development Unit (HUDU) advising local authorities and health agencies on the planning of health facilities through the use of legal planning agreements, extracting financial support from developers and bridging the divide between the two sectors. Evidence review and case study research (in the cases of GLA, HUDU, Plymouth, Glasgow and Bristol) also suggests that broader collaboration can be made effective through a range of methods. The Bristol and Plymouth case studies (UWE, 2011d; NICE, 2011) stress the value of joint appointments between health authority and local authority | - Stakeholders and statutory consultees - Knowledge transfer, good practices | - Externalised expertise - Dedicated posts and capacity - Innovative practices |
| 10 | The Bristol and Plymouth case studies (UWE, 2011d; NICE, 2011) stress the value of joint appointments between health authority and local authority. This has been found to break down silo barriers and greatly assist the integration of health into planning policy and decisions. It can take the form of a jointly appointed director of public health, and a dedicated officer with explicit health and planning responsibilities | Capability, skills and knowledge | Dedicated posts and capacity |
| 10 | According to the Manchester, Bristol as well as Plymouth case studies, this should also apply to major developments: the public health authority can influence the nature of the initial advice given to applicants. Changes to move the public health function into local authorities in England may be beneficial in this respect | Decision-making process | - Impact on implementation - Impact of public health reforms - Frontloading/ upstream |
| 10 | Critical to success is the political and professional commitment at local level. Is there political commitment to health and well-being all along the decision-making process? | Impact of politics and politisation in planning | - Political commitment - Professionalism and impact on profession |
| 10 | Further research is needed to examine how the rhetoric of plans and strategies is implemented on the ground, how planning and other departments prioritise health when defining the purpose and scope of plans and projects | - Evidence base - Local plan content and plan-making process | - Health consideration - Impact on implementation |
| 10 | We found in Manchester, Plymouth and Bristol that a pre-application HIA, with the health and planning authorities helping with scoping, can enable key issues to be addressed in advance and mitigation incorporated at the outset when it is likely to be much more effective. HIA has been identified as a trigger for mutual learning. | - Use of impact assessment - Training, learning and development | - Health impact assessment - Frontloading/ upstream |
| 10 | At the time we carried out our research, the English planning system did not contain any specific planning policy guidance or planning policy statement on health. It has some excellent nonstatutory healthy environment guidance in the fields of urban design, sustainable building design, local transport and street design, open space, green-space and recreation, for instance By Design, the Manual for Streets or the Code for sustainable homes, but the potential health benefits are not always sufficiently explicit and major gaps in official guidance (except at the broad policy level) occur in relation to, for example, accessibility, social inclusion and strategic policy. This is likely to reduce the ability of local authorities to plan healthy urban environments | Strength of national policy | - Clarity in national policy - Impact on implementation |
| 11 | On one hand, as the UK government promotes more ‘localism’ it might be argued that national guidelines and guidance are no longer appropriate. Even before the new UK 2010 Coalition Government, UK planning regulations required evidence backing standards to be locally based where possible | Use of (local) planning powers | Role of local communities |
| 11 | The NPPF does, though, lack precision on how to interpret healthy planning and how to monitor achievements on the ground. The National Indicators (used until 2011) to assess and compare local authority performance, included targets relevant to health and well-being. Although they have been abolished, it could be argued that they were useful tools to promote healthy environments, and had important health implications. They should they continue to be used by local authorities for local monitoring. | - Strength of national policy - Monitoring and evaluation | - Clarity in national policy - Impact on implementation |
| 11 | Similarly, until the 2011 planning reform, the annual monitoring of progress against a wide range of indicators in the English planning system (similar but not identical in the devolved administrations in Scotland and Wales) offered an important and systematic mechanism for promoting healthier environments even if the discretionary elements within the annual monitoring meant that some authorities used more and better health-related indicators than others. Annual reports also carefully monitored progress in achieving healthier environments where specific official guidelines to assist that aim did not exist. Whilst the need to submit these annual monitoring reports to the Government has been ended, local authorities still need to inform the local community of progress. They can therefore continue to be a useful tool for local authorities to review policy and its implementation. To give an example, Plymouth, one of the local authorities that scored highly in our research in terms of mainstreaming of health into its planning documents used their annual report to highlight strengths and weaknesses and show how the weaknesses were to be addressed. We would therefore recommend local authorities to continue on using them. | Monitoring and evaluation | Better of local levers |
| 11 | The move of the public health function into local authorities in 2013 in England offers new opportunities in this respect (DoH, 2010). It will be interesting in time to examine if this policy ambition has the desired results in producing healthier environments. | Impact of planning reform processes | Impact of public health reforms |
| 12 | 2019 | - Capability, skills and knowledge - Resourcing of local planning authorities |  |
| 12 | Challenges in the current English context: In the context of planning policy, a number of issues emerged, linked to the need to continue to develop a robust evidence base to inform policy development. Many seminar participants were aware of the evidence that is currently available, and which may be used in planning and place-making. There was a consensus that currently the evidence base is not joined-up and that much of the required evidence is not currently available in a format that is useful for decision-making – especially at a local level. Practitioners expressed their concern that academic studies were often not well enough informed by the types of questions that practice and policymaking require; that the political nature of planning and the realities of day to day practice and pressure inherent in the system are not addressed. Furthermore, academic researchers do not necessarily present their findings in ways that facilitate decision-making. An alternative view put forward by public health stakeholders is that the evidence base is sufficient, but that decision makers are often not able, or not always willing to act in accordance with evidence-based recommendations | - Evidence base - Impact of politics and politisation in planning | - Type of evidence - Use of evidence base - Impact on implementation |
| 12 | Research showed that much of the available evidence is either not readily usable, not politically feasible, or contravenes legislation. Local practitioners are often unclear about how they can apply evidence of intervention outcomes from academic studies (including health outcomes). They frequently discount academic research as insufficiently relevant to their local area and practice, and instead use a range of other information sources including local routine data, expert consensus and policy reports | Evidence base | - Use of evidence base - Relevance of evidence - Type of evidence base |
| 12 | practitioners highlight their struggle with the continuous emergence of a new evidence base. This reflects past research that questioned whether planning processes are able to cope with the required level of experimentation, flexibility and iterative learning | Capability, skills and knowledge | - Use of evidence base - Innovative practices - Uncertainty |
| 12 | Planning officers stated their need for a usable local evidence base. In order to achieve this, however, change in local policy and practice is required. Local public health strategies, including Directors of Public Health Annual Public Health Reports and the mechanisms to assess local population needs (i.e. Joint Strategic Needs Assessments (JSNAs) and Health and Wellbeing Board Strategies (HWBSs) are key for aligning these agendas. However, policy integration is difficult, according to the stakeholders consulted. Another participant stated that in addition, local JSNAs and local HWBSs often do not reference the importance of the built environment at all, for instance referencing Local Development Plans | Use of (local) planning powers | - Type of evidence base - Lack of alignment - Non planning regulations |
| 12 | In practice, however, there is also a need to build the capacity and skills of planners to effectively use and integrate the information derived from local JSNAs and other health data as well as the evidence from research and guidance from a variety of stakeholders – including those from built environment organisations | Capability, skills and knowledge | - Use of evidence base - External influences |
| 13 | Inconsistencies in national planning policy: The NPPF and the PPG acknowledge the built environment as a determinant of health and planning as an instrument for creating healthy communities. Core “healthy” planning principles are listed in national policy and include quality design, affordability, reduction of pollution, empowerment, mixed-use, heritage, public transport, high quality open spaces, and high quality homes. NPPF and PPG also encourage cross-working between local public health and planning teams, both, at plan and project levels. However, a number of legal issues in the land development process impede the pursuit of this higher objective | - Partnership, collaboration, relationships, engagement - Strength of national policy | - Consistency - Clarity in national policy - Impact on implementation |
| 13 | Opportunities and good practice in spatial policy and tools for health: Within the national policy context, it might be that opportunities for planners to enable health comes from good practice at local level, despite a recurrent issue raised in the seminars, namely that a great deal of ‘reinventing the wheel’ was going on in local authorities, with few mechanisms for effective learning of good practice from each other highlighted | Knowledge transfer, good practices | Innovation |
| 13 | Some local authorities have found ways to integrate public health into planning policy, either at strategic level, in core strategies (i.e. the strategic element of their Local Plan) or at urban development project level in their Site Allocations and Development Management Policies. Good practice in Stoke on Trent, Bristol and Stockport shows how this might be realised by integrating health and planning agendas. The importance of aligning local authority agendas on place, poverty, inequality and the economy is seen as essential by stakeholders consulted to achieving integrated policy-making. Stoke on Trent, Bristol and Stockport have embedded healthy planning principles into their core strategies. The three authorities have followed through their strategies at development management level by developing standards helping to shape new urban developments. | - Use of (local) planning powers - Local plan content and plan-making process | - Impact on implementation - Innovation |
| 13 | Seminars also identified the potential of Impact Assessments to mainstream consideration of health evidence into planning practice. England has not adopted a national Health in All Policy approach, though the NPPF recommends use of HIA for large planning applications and consultation with public health teams. Some local authorities have already started using HIA as part of their healthy planning tools | Use of impact assessment | - Health impact assessment - Clarity in national policy |
| 13 | A key challenge in the post 2012 context remains under-resourcing of the planning system and a heavy reliance on a small number of large construction firms to deliver housing. Local authorities now frequently lack resources and capacity to ensure that effective proactive enforcement can be operationalised, including monitoring and compliance functions. This was supported by many comments made at the seminars | - Resourcing of local planning authorities - Monitoring and evaluation | - Impact on implementation - Role of private sector |
| 14 | The public sector has no significant resources to deliver housing targets and relies on the private sector with short term financial objectives rather than long term health and sustainability ambitions. The challenge is to convince private developers that they can too contribute to shaping healthy communities by endorsing design standards which promote active living, mixed use and mixed income developments without damaging their financial interests. | - Stakeholders and statutory consultees - Impact of economic focus, viability and costs | - Role of private sector - Funding |
| 14 | However, the role of planning should not be underestimated, even in market driven environments. As one developer participant put it “we do not do health because no one is asking us to”. | Strength of national policy | Role of private sector |
| 14 | One thing which is certain is that the key challenges for evidence, partnership working and strong policy integration by practitioners at national and local level will remain. Policy integration such as the one described in section 5 requires an effective, tried and tested cross-sector collaboration between planning and public health and other relevant teams. Pre-2012 research showed that best practice in England depended not so much on the planning system per se, as on leadership, commitment, knowledge of politicians and practitioners involved. | - Partnership, collaboration, relationships, engagement - Leadership - Impact of politics and politisation in planning |  |
| 12 | Local planning policies can restrict urban features promoting unhealthy behaviour, e.g. restricting hot-food takeaways in close proximity to schools and youth facilities. However, indicators offer more comprehensive guidance to plan healthy developments and inform planning policies (e.g. fast food takeaways) if they cross reference local public health policies and targets. For instance, Gateshead’s use of childhood obesity targets in their Supplementary Planning Guidance provides a much firmer footing on which to make planning decisions and fight appeals, than more generic targets of for example preventing fast food outlets with 400m of schools (FUSE, 2018) | Use of (local) planning powers | Non-planning regulations  Impact on implementation |
| 12 | The priority given to short term viability as well as systems construct challenges could work to counteract the focus on sustainable, healthy communities promulgated in the rest of the NPPF | Impact of economic focus, viability and costs |  |
| 12 | However, the whole development process and its culture need to evolve and will only do so with the buy-in from the key stakeholders and actors of development. | Stakeholders and statutory consultees | Role of private sector |
| 12 | The research suggests that in those authorities that have made real progress with integration, this is driven by key individuals able to bridge the public health-planning gap, understand sectoral priorities and how institutions work to break through silos and promote joint agendas, tools and practice. The case studies of Bristol, Stoke-on-Trent, Stockport have demonstrated this point. The importance of academic-practice links and collaboration has also been emphasised in Bristol for instance to develop and implement the concept of healthy planning. | - Capability, skills and knowledge - Partnership, collaboration, relationships, engagement | - Governance - Externalised expertise (academia) |
| 13 | In this context, impact assessments (IAs; in particular HIA and health inclusive SEA and EIA) can play a key role. This can enable local authorities to build institutional capacity, create processes, policies, lines of accountability and engage with communities. Yet, the practice of IA (HIA and health inclusive SEAs / EIAs) needs to be revisited as it is currently not effectively used by local planning authorities consistently across England in order to support healthy planning. There is currently a lack of capacity to conduct IAs effectively. | - Use of impact assessment - Capability, skills and knowledge | - Environmental impact assessment - Health impact assessment - Health considerations |
| 13 | Local planning authorities can start by promoting a consortium approach to place-based, pro-active planning and design. | - Visioning and outcomes-focused - Partnership, collaboration, relationships, engagement | Governance |
| 15 | 2022 | Resourcing of local planning authorities |  |
| 15 | A lack of detail in reform initiatives, or resource to support their implementation, as well as potential conflicts between different government policies – the inherent complexity and contradictoriness of state institutions – means that frontline local authority planners must make sense of policies from central government as they’re put into practice | - Resourcing of local planning authorities - Complexity of planning regulatory framework | - Uncertainty - Impact on implementation |
| 15 | Ambiguities in reform agendas open up spaces for institutional entrepreneurs to protect and further their own interests | Impact of planning reform processes | - Uncertainty - Role of private sector |
| 15 | Local authority planners must utilise legislation and policy instruments introduced by central government (even whilst sometimes exploiting ambiguities in them) and must face the materiality of austerity-driven grant cuts leading to fewer planners (even whilst sometimes using changed financial contexts to justify their role). This is compounded by the powerful notion that part of being a ‘good planner’ includes following policy and legislation | - Use of (local) planning powers - Resourcing of local planning authorities | - Better use of local levers - Funding - Professionalism and impact on profession |
| 16 | 2015 | - Resourcing of local planning authorities - Capability, skills and knowledge |  |
| 16 | Political leadership - Many of the local authorities who responded to the survey and all of those featured in the case studies (see Part 2) have a strong commitment to meeting the housing challenge. However, concerns were raised that there needs to be consistent, national prioritisation of affordable and social housing by the UK Government, Scottish Government and Welsh Government. Many councils identified leadership and business skills as a key barrier to bringing forward innovative new models of housing | - Leadership - Capability, skills and knowledge | - Clarity in national policy - Commercial and development economics skills |
| 16 | The key issues raised around planning were getting a Local Plan in place that demonstrates | Local plan content and plan-making process | Up to date local plan |
| 16 | Of the respondents surveyed just over half (53%) reported to have an up-to-date plan with an adopted five year land supply. The five year land supply requirement was a key concern of many of the local authorities who responded, due to environmental and planning designations such as Green Belt and economic constraints such as viability. 41% of the 111 councils who responded to this particular question did not have an ‘up-to-date’ local plan with an adopted five year land supply; a further six local authorities did not know whether they did or not. The absence of a Local Plan is clearly a major issue for housing delivery and appears to be related to both resources, political consensus and the workability of national government policy. | - Local plan content and plan-making process - Impact of economic focus, viability and costs - Resourcing of local planning authorities - Impact of politics and politisation in planning - Strength of national policy | - Up to date local plan - Clarity in national policy |
| 16 | The plan preparation and review process should allow authorities to use a solid evidence base to take a holistic and long-term view and consider the most sustainable options for the scale and location of future growth and regeneration. This requires a range of assessments, including evaluations of housing requirements, urban capacity (for example Strategic Housing Market Assessments), employment, the economy, flood risk, transport, biodiversity, landscape, and energy production needs and capacity | - Use of impact assessment - Evidence base | - |
| 16 | Survey respondents expressed concern about a ‘lack of resources’ and ‘specialist skills’ needed to undertake these assessments and in particular the need for Government to set out ‘clearer guidance on Strategic Housing Market Assessments’ | - Resourcing of local planning authorities - Capability, skills and knowledge |  |
| 16 | “duty to cooperate was not designed to a proxy for strategic planning at a wider sub-regional or regional scale.” While there is evidence that the duty to cooperate has driven usual cooperation, there was a particular issue in high demand areas where local authorities who failed to comply with the duty are left without a sound legal framework for the delivery of housing need through planning | Impact of strategic planning practices |  |
| 17 | Some county council respondents suggested that there needed to be ‘better co-ordination between districts’ in some areas, in terms of both planning for housing and the management of housing stock. One respondent recommended that ‘County Structure Plans’ are re-introduced and others suggested that a return to some form of ‘strategic planning’ is needed. There was a powerful consensus at the roundtable discussion that to be effective, plans need to reflect functional housing market areas. | Impact of strategic planning practices | - |
| 17 | In England the National Planning Policy Framework (NPPF) introduced a new policy on viability testing. The test is based on a straight forward residual valuation, but it is framed narrowly to “provide a competitive return to willing developers and land owners”. In response to the question ‘has the viability test, as set out in the NPPF, helped or hindered your local authorities ability to secure sufficient social and affordable housing to meet local needs?’ over half (54%) of the 95 councils that responded to this question said it had hindered, | - Impact of politics and politisation in planning - Impact of economic focus, viability and costs - Impact of planning reform processes | - Impact on implementation |
| 17 | Construction skills and capacity - At the roundtable discussion a representative from APSE rightly highlighted, that in addition to “…developing policies, liberating local authorities, encouraging investors and identifying land, it is essential that the construction industry has the capacity to be able to build because without them the problem will not be addressed.” It was recognised that the Government needed to provide support to the supply chain, promoting and providing further funding for training and encouraging technological change within the sector | - Capability, skills and knowledge - Training, learning and development | - Funding - Innovative practices |
| 18 | 2024 | Resourcing of local planning authorities |  |
| 18 | For the fourth consecutive year, SME home builders have consistently identified planning process challenges as the top two obstacles to their growth | Complexity of planning regulatory framework | - |
| 18 | Taking the top spot was ‘delays in securing planning permission or discharging conditions’ which was considered a major barrier by 93% of SME developers | Decision-making process | Cost and delay |
| 18 | This was followed closely by a ‘lack of resources in Local Planning Authorities (LPAs)’ which was declared a major barrier by 90% of respondents | Resourcing of local planning authorities | - |
| 18 | PLANNING COSTS - At a time when the planning service is performing poorly, developers are also having to absorb increases in the costs of achieving an implementable planning increase. This is compounding an already difficult situation further, particularly for SMEs. When asked whether the costs of obtaining implementable planning permission have changed in the past three years, 97% of respondents believed the costs had increased | Decision-making process | Cost and delay |
| 18 | POLITICAL DECISION MAKING- Some of the most significant challenges faced by developers with regards to planning extend beyond practical matters such as resourcing. This year’s survey results show that political decision making is a significant barrier to growth for SME home builders, and that SME developers are dissatisfied with the current Government’s approach. The Government’s approach was also considered to be positive for those who oppose the building of new homes | Impact of politics and politisation in planning | - |
| 19 | PLANNING CONSTRAINTS - In addition to being asked about the major barriers to development generally, respondents were also asked to specifically rank the biggest constraints in the planning process. There was little change at the very top of the table in comparison with other years, with staff resourcing/shortages being seen as the number one major constraint (68%) | Resourcing of local planning authorities | - |
| 20 | 2022 | Resourcing of local planning authorities |  |
| 20 | 92% said a lack of resources in Local Planning Authorities (LPAs) was a major barrier to growth (up from 90% in 2021) | Resourcing of local planning authorities | - |
| 20 | This is supported by the anecdotal comments received from developers of all sizes, across the country, who broadly consider the current state of the planning process to be the worst it’s ever been. While this situation is frustrating for all developers, it is acutely challenging for SME builders, who by their very nature are less well-equipped to mitigate these issues. For example, if a large developer faces delays on a particular site, it will at least have multiple other sites that it can progress in the interim. SMEs, on the other hand, may have their capital tied up in just one or two projects at a time. As a result, lengthy delays can bring their business grinding to a halt. | Complexity of planning regulatory framework | Cost and delay |
| 20 | While questions over the detail of the Government’s planning policy remain, it is clear that the obstacles in the planning process are varied and vast. In response to a deep dive question about the biggest constraints in the planning process, red tape, political decision making and associated services, such as highways approvals, all emerged as further constraints that need to be addressed. | - Impact of politics and politisation in planning - Impact of planning reform processes - Complexity of planning regulatory framework | - |
| 20 | Respondents were also asked for their views about the impact the forthcoming infrastructure levy is likely to have on the provision of affordable housing. 39% of respondents felt affordable housing would decrease, a third (33%) did not know and 23% felt the provision would remain the same. Only 5% believed affordable housing provision would increase. | Impact of planning reform processes | - |
| 20 | Ensure planning departments are adequately resourced and funded | Resourcing of local planning authorities | Funding |
| 21 | 2021 | Resourcing of local planning authorities |  |
| 21 | The survey results showed that the planning process was, again, the most significant challenge that SMEs face and that the problems with the process have worsened over the last 12 months | Complexity of planning regulatory framework | - |
| 21 | 94% of respondents see delays in securing planning permission or discharging conditions as a major barrier to housing delivery. This is up from 83% in 2020 | Decision making | - Impact on implementation - Cost and delay |
| 21 | 90% of respondents believe a lack of resource in local authority planning departments is a major barrier. This is up from 73% in 2020 | Resourcing of local planning authorities | - |
| 21 | In particular, the length of time it takes for some planning applications to reach fruition was repeatedly raised – most respondents could give instances of a development that had taken years for a planning resolution | Decision-making process | - |
| 21 | highlighting the planning process as a major barrier in this most recent survey, it is of ever increasing importance that these are addressed by the reforms that have long been promised by the Government | Impact of planning reform processes | - |
| 21 | The top three suggestions as to how the Government can help SMEs were:  1. REFORM THE PLANNING PROCESS, SPECIFICALLY REMOVE RED TAPE AND BETTER EQUIP PLANNING DEPARTMENTS | - Complexity of planning regulatory framework - Resourcing of local planning authorities | - |
| 21 | 2020 | Resourcing of local planning authorities |  |
| 21 | The survey results indicate that one of these challenges, planning, continues to be of particular concern. When respondents were asked about the challenges to increasing housing delivery over the next twelve months, planning issues were cited as the major barrier. | Complexity of planning regulatory framework | - |
| 21 | 83% of respondents see delays in securing planning permission or discharging of planning conditions by local authorities as a major barrier to delivery | Decision making | - Impact on implementation - Cost and delay |
| 21 | 73% of respondents believe a lack of resource in local authority planning departments is a major barrier | Resourcing of local planning authorities | - |
| 21 | SMEs are unable to effectively mitigate these setbacks as they may have their capital tied up in just one or two projects at a time. As a result, lengthy delays can bring their business to a halt. On 6 August, the Government unveiled its Planning for the Future white paper, containing a range of measures that it intends to streamline the planning process and enable new homes to be built faster. While we are hopeful that the reforms will deliver positive changes for home builders and particularly SME developers, the scale of the proposed changes means that the transition will take time. As such, it is imperative over the coming months that Local Authorities do not use this period of flux to halt the development of Local Plans. | - Complexity of planning regulatory framework - Impact of planning reform processes - Local plan content and plan-making process | Cost and delay |
| 21 | THE TOP THREE SUGGESTIONS WERE:  3. REFORM THE PLANNING SYSTEM" | Impact of planning reform processes | - |
| 22 | 2020 | - Capability, skills and knowledge - Resourcing local planning authorities |  |
| 22 | Planning professionals felt empowered to regulate takeaway food outlets because of “tools” in place. The planning system was considered unique because of the ability to target takeaway food outlets, and expand, rather than limit, choice. There was a perceived obligation to act because planning power had been provided to local government areas and should therefore be used | Use of (local) planning powers | Better use of local levers |
| 22 | Takeaway food outlet regulation was not always considered a priority. Resource shortages, competing economic priorities, political pressure, and the need for housing provision prevented prioritisation | - Impact of politics and politisation in planning - Impact of economic focus, viability and costs - Resourcing of local planning authorities | - |
| 22 | Local evidence in the form of data and statistics were also discussed. Evidence needed to be relevant to a local government area and robust to help justify planning policy adoption.  Peer-reviewed, scientific evidence was considered to support planning policy adoption. However, when discussing health focused approaches, the ability of this evidence to demonstrate takeaway food outlet impact was questioned.  Whilst the importance of peer-reviewed scientific evidence was understood there was a difference in opinion on how it could be used. Some Planning professionals questioned whether causal relationships could be drawn from correlational evidence. Public Health professionals seemed more open to results of this nature. | Evidence base | - Use of evidence base - Relevance of evidence - Type of evidence |
| 23 | 3.5. Internal stakeholder co-operation - Planning and Public Health professionals recognised that cross-department collaboration facilitated adoption. Professionals in each department had unique roles. Public Health professionals ensured a health focus was embedded within planning policies and provided supporting local data and evidence. Planning professionals drafted policies that complied with the National Planning Policy Framework. Devolution of responsibility for public health to local government in 2013 was recognised as an important milestone in building this relationship. | Stakeholders and statutory consultees | - Health consideration - Professionalism and impact on profession - Impact of public health reforms |
| 23 | The need for shared understanding of professions, priorities and goals to help develop applicable and realistic policies was recognised. New roles could link often siloed departments | Capability, skills and knowledge | - Impact of public health reforms - Dedicated posts and capacity |
| 23 | Planning policy adoption was further supported by local politicians. They act as leaders, raise concerns, promote regulatory need, and endorse planning policy adoption. | - Impact of politics and politisation in planning - Leadership |  |
| 23 | Challenge to planning policy - Objections to planning policy adoption by national and international fast-food chain representatives could undermine efforts and lead to amended planning policies. This was a recognised concern.  Local government areas initially determine the acceptability of planning applications. Where permission is refused, applicants may appeal, and a final decision is made by the National Planning Inspectorate. One participant described how a decision to refuse planning permission for a new takeaway food outlet was overturned. This caused confusion, frustration, and a feeling that national guidance had been contradicted | Decision making | - External influence - Impact on implementation - Planning Inspectorate |
| 23 | There could also be internal challenges. Local politicians sometimes considered planning policy adoption to be a “nanny-state” approach. This perspective, without an appropriate response from professionals in practice, could create a barrier towards adoption | Impact of politics and politisation in planning | - Role of elected members - External influence |
| 24 | 2021 | - Capability, skills and knowledge - Resourcing local planning authorities |  |
| 24 | The top three organizations/professionals perceived by survey respondents to impede the integration of spatial planning and health evidence at the local level were private developers, private sector consultants and planners in Planning Inspectorates | - Evidence base - Stakeholders and statutory consultees | - Role of private sector - Planning Inspectorate |
| 24 | there was a difference between public health and planning professions in their understanding and the use of evidence that was highlighted as a key barrier to collaborative working between public health and planning professionals. Planning professionals emphasized that policy and national standards are the most important sources of evidence, whilst public health professionals cited research evidence as most important. | Evidence base | - Use of evidence base - Type of evidence base - Professionalism and impact on profession |
| 24 | economic arguments with developers were seen as a key barrier, with practitioners noting that developers would consider the statutory obligations but are less concerned with intangibles such as health that can impact on their profit margin | Impact of economic focus, viability and costs | Health considerations |
| 24 | some practitioners expressed concern that a lack of political support at the local level makes it difficult to influence local policies that ensure health is appropriately integrated into spatial planning | Impact of politics and politisation in planning | Health considerations |
| 24 | Practitioners also argued that existing legislation is not strong enough to see substantial improvements in healthy place-making and that stronger legislation with explicit links to health integration is needed to engage with developers | Strength of national policy | - Increase regulations - Clarity in national policy - Health considerations |
| 24 | issues of resource and capacity at local authority level were identified, with concerns raised about the impacts of reduced local authority budgets on the availability of resources and on the skillset needed to support collaborative work between public health and planning | - Resourcing of local planning authorities - Capability, skills and knowledge | - Lack of engagement |
| 24 | Nine out of 10 respondents agreed that a lack of evidence that can be translated to practice at the local level is an important barrier to health integration into spatial planning at the local level | Evidence base | - Health considerations |
| 24 | 89% of respondents considered the reduced capacity to be a major barrier (Table 3). | Resourcing of local planning authorities |  |
| 24 | building relationships with developers was seen as important to promote values of healthy place-making | - Partnership, collaboration, relationships, engagement | - Role of private sector - Deliver wider value and benefits |
| 24 | articulating the wider benefits for multiple stakeholders: practitioners identified that an important step to addressing siloed working across various sectors is to articulate the wider benefit of integrating health into planning to multiple stakeholders including developers, local authority, the NHS and other sectors. | - Visioning and outcomes-focused | - Deliver wider value and benefits - Role of private sector |
| 24 | both public health and planning professionals agreed that simplifying the presentation of evidence in terms of the language and accessibility to both fields enables collaborative working | Evidence base | - Relevance of evidence base - Access to evidence base - Lack of engagement |
| 24 | Nearly all respondents (96%) agreed that integrating health into the Local Plan is an important facilitator of healthy spatial planning | Local plan content and plan-making processes | - Health considerations |
| 25 | The findings from this study also demonstrated that difficulties in translating evidence, for different audiences with differing needs, were a key barrier in getting evidence into practice at the local level | Evidence base | - Relevance of evidence base |
| 25 | A lack of resource and skillset to support collaborative work between public health and planning was reported as the second most important barrier facing local professionals | - Resourcing of local planning authorities - Capability, skills and knowledge | - Lack of engagement - Access to evidence base |
| 25 | There is a need to integrate local health and well-being needs and priorities into the Local Plan and decision-making process | - Local plan content and plan-making process - Decision-making process | - Health considerations |
| 25 | Heads of Planning play an essential role in ensuring that Local Plans are up to date and meet not only the generic health and well-being requirements in the NPPF and the National Planning Practice Guidance8 but also link to local needs as outlined in Joint Strategic Needs Assessments (JSNAs). This also requires that Directors of Public Health and their teams should ensure that all health and well-being strategies (and healthcare strategies) refer to the environmental aspects of disease causation and how the built environment can be modified to support health and well-being | - Leadership - Local plan content and plan-making process | - Up to date local plan - Health considerations - Better use of local levers |
| 25 | A joint basic understanding of the impacts of the built environment on health and the systems and processes which are used by both built environment and public health professionals through a local training programme is pivotal in addressing cultural gaps.4,17 Training could be jointly delivered with key partners such as the professional institutes and universities and targeted across the spectrum of the career path from undergraduate modules to professional continuing professional development | - Training, learning and development | - Professional qualification |
| 25 | Political support is essential to ensure that improvements to health underpin all planning decisions at the local level. Political support from elected members and clear corporate priorities were identified as crucial determinants of the extent to which health is integrated into spatial planning | - Impact of policies and politicisation in planning - Decision-making | Health considerations |
| 25 | There was a perceived need to improve access to existing wealth of knowledge with strong support for a central repository of good practice for sharing good practice across both disciplines. Practitioners would appreciate clearer signposting and access to this information, a | Knowledge transfer, good practices | Access to evidence base |
| 24 | Improving national guidance and having stronger policies for place-making and health were ranked as the most important recommendations, while organizing networking events was ranked as the least important recommendation | - Strength of national policy - Knowledge transfer, good practices | Clarity in national policy |
| 26 | Five years on from the NPPF’s introduction of a “less complex” planning system, over 60% of LPAs are still without a Local Plan tested and found sound against national policy, with the majority of those still to get to the starting blocks of a local plan examination. A slowly emerging spatial pattern, illustrated in Figure 2, highlights how plan-making is lagging in some particular areas including authorities surrounding Manchester, Birmingham and London where difficult choices about Green Belt appears to be halting progress. | - Local plan content and plan-making progress - Impact of planning reform processes | Soundness  Cost and delay |
| 26 | Before the introduction of the NPPF the average time from submission to a plan being found sound was 9.7 months. However, since the introduction of the NPPF, this has increased to an average of 16.8 months, with some taking upwards of three years to pass through the examination process. Obviously, prior to the NPPF, | - Local plan content and plan-making progress - Impact of planning reform processes | Soundness |
| 26 | Early plan reviews - With almost a third of plans having to commit to an immediate or early review to be found sound, such mechanisms have been regularly used by Councils and Inspectors alike to get plans across the line. In July 2015, then housing and planning minister Brandon Lewis, highlighted in a Written Ministerial Statement how early reviews might help reduce delays in plan making | Local plan content and plan-making progress | Cost and delay |
| 26 | Increasing planned supply  One explanation for the time it takes to get plans through the examination process is that, following scrutiny, there is often a necessary ‘patching-up’ of evidence which results in different housing requirement figures from that originally proposed. Almost half of plans have had to change their housing targets through the examination process, with the vast majority of those required to increase planned housing numbers, albeit a few plans more recently have seen reductions (Figure 4 | - Evidence base - Local plan content and plan-making process | Use of evidence base |
| 26 | Understanding that there are no ‘silver bullets’ to overcome the barriers associated with building more homes, the White Paper took a considered, deal-making approach – councils, housebuilders and other organisations would receive financial or policy assistance in return for compromise elsewhere | - Stakeholders and statutory consultees | - |
| 28 | 2023 | - Capability, skills and knowledge - Resourcing local planning authorities |  |
| 28 | Decision making throughout the process - The appeal process was perceived as confusing and difficult to navigate for some, especially to the public and to those new to the practice, although procedural guidance is published and available to view on the PINS website this was not referred to in any of the interviews | Decision making | Planning Inspectorate |
| 28 | Only consider evidence presented to them. I t was frequently noted how the PINS will only consider evidence if it is presented directly to them and that this was the responsibility of the case specific officer. It was stated that appellants should not assume that PINS know anything about the available evidence, requiring a systematic and thorough approach to pulling together all available evidence to support a case. - General consensus over certain types of evidence. Across interviews with all three professional groups (public health, planning, and the Planning Inspectorate), there was agreement that certain types of evidence were prioritised over others. Some forms of evidence were perceived to be undisputable and essential to a successful appeal (e.g. reference to Local Policy), while others were seen as ‘anecdotal’, unreliable, and to generally be avoided (e.g. the views of the public). - Local plan/ policy and statistical (data) evidence. Two forms of evidence were highly cited by respondents: reference to the local plan and or planning policy (n = 5) and statistical evidence or quantitative data (n = 6). These forms of evidence were regarded as the ‘gold standard’ and often necessary for an appeal to be successful. For councils based in London, the London Plan was perceived as carrying significant weight in comparison to the local plan which was considered more generic. - Academic, authoritative and expert evidence. Academic, authoritative or expert evidence was likewise cited as useful in a planning appeal. Examples varied from peer-reviewed academic papers (particularly systematic reviews), data from PHE (now OHID), government publications, legislation, administerial statements, relevant authoritative groups or professionals, even comments from The House of Commons. However, this type of evidence was not referred to as frequently, and when referenced, it was often to endorse or complement a prior argument, which would have already been supported by one of the previous two primary sources of evidence | - Evidence base - Capability, skills and knowledge | - Planning Inspectorate - Access to evidence - Type of evidence - Relevance of evidence |
| 29 | Another notable point was that although the Planning Inspectorate was viewed across all professional groups as fair and neutral; the perception from public health was that they didn’t think the inspectorate gave enough weight to public health evidence, or that they had to go out of their way to ‘state the obvious’, in that a new takeaway would be unhealthy and cause harm. On the other hand, Planners and the Inspectorate were keen to point out that planning policy and the appeals process is not designed solely with public health in mind, and that achieving public health objectives is not as simple as limiting the number of HFT | Decision-making process | - Use of evidence - Deliver wider value and benefits |
| 29 | Communication. Communication played a significant role in putting an appeal case together. Cross-department working, knowing who to approach in an LA as well as where to find outside sources of information that could add value to a case (such as academic papers, reports and statistics) were believed to facilitate the process. Absence of working in a multidisciplinary way was perceived to impact on the ability to collate evidence for a case | Partnerships, collaboration, relationships, engagement | - Externalised expertise - Lack of engagement - Access to evidence - Governance |
| 29 | Accessibility of evidence and data. Access to both national and local data was considered important. Health statistics were cited as being central by some. This was also believed to be important even if there were already local plans and policies in place. | Evidence base | Access to evidence |
| 29 | - Storage and updating of information. Having up-to-date information at hand was stated by some as being useful in helping to collate and respond to cases, making it less time consuming to collect. | Knowledge transfer, good practices | Access to evidence |
| 29 | Format of evidence. It was not only important to have this information readily available but it was also important that it was usable. Often it was deemed to be in a format which was tricky to interpret or make sense of, and therefore could be difficult to use | Evidence base | - Type of evidence - Use of evidence |
| 29 | Understanding the importance of health. Understanding the importance of health and the implications of health on the wider planning agenda was considered valuable. Several participants felt this acknowledgement was lacking across professional groups, including planners. There were suggestions for additional training on the topic | - Capability, skills and knowledge - Visioning and outcomes-focused - Training, learning and development | - Health considerations - Professionalism and impact on profession |
| 28 | (1) Local Authorities should ensure that a clean and robust local plan is in place and applied correctly | Local plan content and plan-making process | - |
| 28 | (2) Having adequate time and staff capacity to deal with appeals cases/acknowledging time required to deal with an appeal | - Resourcing local planning authorities - Decision-making |  |
| 28 | (3) Having access to accurate, robust, and up to date local information to use in the appeals case. | Evidence base | Access to evidence |
| 28 | (4) Having firm commitment from elected members and senior management from various professional groups (such as Planning and Public Health). | - Impact of politics and politisation in planning - Leadership | Political commitment |
| 28 | 5) Good lines of communication with relevant local groups & communities interested in the appeal.  (6) Using clear, concise language that is accessible across professional groups." | Partnership, collaboration, relationships, engagement | - Access to evidence - Role of local communities |
| 30 | 2020 | Capability, skills and knowledge  Resourcing of local planning authorities |  |
| 30 | with legislation and current policies | Strength of national policy |  |
| 30 | (the lack of) expertise | Capability, skills and knowledge |  |
| 30 | (different) ‘working cultures’ across different teams, combined with the problem of accessing ‘hard to reach groups’ and/or successfully involving communities in their projects. | Stakeholders and statutory consultees | - Governance - Role of local communities |
| 30 | What kind of skills were needed to overcome barriers/obstacles or oppositions | Capability, skills and knowledge | - |
| 30 | The most important skill deemed essential by our participants to overcome barriers and obstacles to the implementation of healthy placemaking principles was (by far) the ability of planners to collaborate with other professionals (especially public health professionals but also developers). | Partnership, collaboration, relationships, engagement | - Role of private sector - Health considerations - Impact on implementation |
| 30 | Participants also cited ‘leadership’, ‘communication’ and the ability to ‘develop personal knowledge and new skills’ as important factors to overcome challenges | - Leadership - Partnership, collaboration, relationships, engagement | - |
| 30 | Engaging with a variety of stakeholders — ranging from public health ‘leads’ to community leaders and end users — was deemed critical to successful project delivery. In addition, engaging with communities and developers early on in the process was considered a key component of a successful strategy. | Stakeholders and statutory consultees | - Role of local communities - Role of private sector - Frontloading/ upstream |
| 30 | Other key drivers of success mentioned by participants included adequate funding, political interest (in the project/initiative) and alignment with other policy objectives and priorities | - Resourcing of local planning authorities - Impact of politics and politisation in planning | - Funding - Political commitment - Lack of alignment - Deliver wider value and benefits |
| 30 | Our findings suggest that despite trying circumstances (e.g. lack of resources and capacity, lack of expertise and formal partnerships across teams) successful projects have been driven by planning professionals who initiated and developed collaborative, innovative and ‘integrated’ approaches to healthy placemaking | - Partnership, collaboration, relationships, engagement - Resourcing of local planning authorities | Innovative practices |
| 30 |  Effective collaboration and clear communication between planning and public health teams/professionals.   Developing new skills and acquiring expertise in areas cutting across planning and health.   Stakeholder engagement – especially developers and local communities.   Clear and transparent processes | - Capability, skills and knowledge - Partnership, collaboration, relationships, engagement | - Role of local communities - Role of private sector - Professional qualification |
| 31 | Responses to our call for evidence, including interviews with research participants, strongly suggest a real ‘willingness’ from planning professionals to integrate principles of healthy placemaking in their decisions | Decision-making process | Health considerations |
| 31 | More often than not, efforts to collaborate with other teams and to acquire technical expertise on health-related topics in a short space of time were undertaken on an individual and voluntary basis; motivated by sincere commitments to plan for ‘the public good’. | Capability, skills and knowledge | - Deliver wider value and benefits - Professionalism and impact on profession |
| 32 | 2020 | - Resourcing of local planning authorities - Capability, skills and knowledge |  |
| 32 | , the initiation of such policies was driven largely by central government guidance, namely, the NPPF and NPPG (National Planning Policy Guidance), previous Planning Policy Statements (PPS) and related planning legislation. The response suggests that a strong top-down influence drives climate change policy formulation in plan making | Strength of national policy | Clarity in national policy |
| 32 | Relevant local plan policy themes also tended to reflect local experiences of climate change hazards, in particular, extreme flooding events. In part, this focus on experienced flood hazards was because these issues were at the forefront of the minds of the planners writing these policies, as well as elected members and the local public. It was indicated that these were not only issues that had been experienced recently, but they were also hazards that were perceived to have increased most noticeably over the last few years and presented the most significant threat to impeding development and growth | Visioning and outcomes-focused | - Impact on implementation - Professionalism and impact on profession - Role of local communities - Role of elected members |
| 32 | flooding and coastal change was something for which the planners had the most well developed and extensive evidence base. As such, writing strong evidence-based policies was considered to be much easier than for some other climate change hazards, such as the threats from increased summer temperatures and heat waves | Evidence base | - Access to evidence base - Confidence |
| 32 | - At least four respondents, however, acknowledged that the emphasis of central government leadership on climate change adaptation was changing or had changed. Contemporary central government priorities were perceived to be related to economic growth and delivery of new development (such as the current Conservative government’s pledge to raise housing supply to 300,000 per year, on average, by the mid-2020s). This emphasis had somewhat subordinated climate change action in local planning | - Impact of economic focus, viability and costs - Strength of national policy - Leadership | - |
| 32 | At least four of the respondents believed that this national political rhetoric filters down to local level leadership and, subsequently, also influences local elected members’ priorities. Thus, local government imperatives, as with central government, are shaped primarily by the economic development agenda and adaptation to climate change has been relegated to a lower priority position | - Strength of national policy - Impact of economic focus, viability and costs - Leadership | - Political commitment |
| 32 | Whilst respondents generally remarked that their elected members seemed reasonably aware of the need to respond to climate change, three respondents believed that the issue was not a big enough political priority to motivate the elected members to “drive” policy formulation in that area themselves. Two respondents highlighted that the policies have been formulated because of the persistence of a few individuals within the planning department itself, rather than politicians | Leadership | - Political commitment - Professionalism and impact on profession |
| 32 | the effect of public spending cuts on the limited resources of LAs was also recognised. There was a general consensus that things felt like they had gone backwards, with officers recognising a reduced profile for climate change within the LAs, including the loss of in-house knowledge and skills. One individual felt that austerity measures and ensuing restructuring within the department had led to the erosion of knowledge relating to climate change in their team with impacts upon Local Plan development. Another respondent held the same view, stating that there was no longer the budget to fund more specific climate change expertise in house as it “was not a government priority” (Respondent P6) | - Resourcing of local planning authorities - Capability, skills and knowledge | - Funding - Governance - Dedicated posts and capacity |
| 32 | further important factor in the failure of LAs to deliver climate change adaptation as “public goods” through new development was the propensity of developers to challenge such conditions on the basis of economic viability | Impact of economic focus, viability and costs | - Cost and delay - Role of private sector |
| 32 | Viability was expressed as a source of frustration by one planner, who remarked that many adaptations included in new buildings, such as plug sockets being placed above the level of possible flood-water incursion, could save a lot of money in the long term. | Impact of economic focus, viability and costs | Cost and delay |
| 33 | Development management decisions - Development management officers reported that the level of awareness about climate change demonstrated by planning permission applicants was limited (62%) or rare (28%). | - Capability, skills and knowledge - Decision-making process | - |
| 33 | this challenge places a bigger burden on the planning officers to make up the shortfall in adaptation action in two ways. First, through awareness-raising about the need for adaptations in pre-application meetings and, second, through resilience- building through the use of planning conditions to ensure that development proposals are adequately future-proofed against climate change. Additional comments suggested that climate change was often seen as an implicit expectation of developers by the planning department and not something that requires a specific conversation in pre-application discussions. | Decision-making process | - Frontloading/ upstream - Impact on implementation - Role of private sector |
| 34 | Local planning authorities should no longer systematically control what specific activity can take place on individual land plots, nor should they attempt to calculate how much space is ‘needed’ by local households and firms and set policy accordingly. A looser binary zonal system should be introduced instead. This would allow obsolete land uses to be recycled much more quickly and local planning authorities would no longer micro-manage land markets | Local plan content and plan-making process | Focus on process |
| 34 | rather than produce extensive local planning documents, local planning authorities should produce a definitive and limited set of rules detailing what type and form of development is not acceptable in their urban area. Developments would then be permitted as long as they are not forbidden. | Local plan content and plan-making process | Focus on process |
| 34 | This should be a reform programme that cuts across government departments, not least HM Treasury, the Ministry for Housing, Communities & Local Government and the Department for Transport. Planning reform would, after all, be a major structural reform of the economy. | Impact of planning reform processes | Focus on process |
| 35 | need for greater coherence regarding health requirements in local plans and national planning policy and guidance | - Local plan content and plan-making process - Strength of national policy | - Health considerations - Clarity in national policy - Lack of alignment |
| 35 | We suggest that this review could help enhance guidance about conducting HIAs of local plans. This includes (i) incorporating the assessment of policy references to local health priorities and national guidance, (ii) reflecting evidence on determinants of health and (iii) including language that promotes the effective implementation of policy requirements. Similarly, where SEAs/ SAs are applied, LPAs are recommended to adopt greater transparency about the specific health elements that are appraised in the review of local plans | - Use of impact assessment - Evidence base | - Health impact assessment - Health considerations - Use of evidence base - Impact on implementation - Environmental Impact Assessment/ SEA |
| 36 | Furthermore, there were very strong feelings that the targets were distorting planning practice through game playing and a range of unintended consequences, so that the service offered to all users of the planning system became worse overall. There were also suggestions that efficiency alone should not be what matters for planners, and concerns that targets somehow deprofessionalize planners, making them administrators | Strength of national policy | - Focus on process - Confidence |
| 36 | There was actually some significant value to professional work from the time-based targets, which had apparently altered practice and opened-up spaces for institutional actors to further their own interests. This fits with what the institutionalist frame would lead us to expect when a strong new rule structure is implemented, and highlights the importance of a nuanced, empirically informed account of managerial governance | Stakeholders and statutory consultees | - Focus on process - Governance |
| 36 | The abilities of frontline planners to influence the very neoliberal reforms they are enacting might thus be the best guardian against the complete reduction of planning to nothing more than an administrative tick-box exercise where little value is added. | Impact of politics and politisation in planning | - Focus on process - Professionalism and impact on profession |
| 37 | 2019 | Capability, skills and knowledge |  |
| 37 | A professional commitment towards a better environment appeared to be a generative mechanism for sustainability practices and underlying conditions included professional identity, identity as a public sector worker, organisational and team identities, and personal commitment | Capability, skills and knowledge | Professionalism and impact on profession |
| 37 | Professional identity as a generative mechanism of action towards sustainable development offers one ‘lever’ by which to increase efforts, but appears unlikely to be effective in a situation where planners are unclear on their responsibilities in a context of weak policy and the competing influence of multiple stakeholders | - Strength of national policy - Stakeholders and statutory consultees | - Professionalism and impact on profession - Clarity in national policy - External influence |
| 37 | The unexpected finding from the study was that of identity as a public sector worker. As identities as a public sector worker, organisational employee and team member also may act as enabling conditions, the local authority as employer organisation has an important role to play here. Valuing their professional planners, supporting planners’ professional development and recognising their professional judgement may strengthen beneficial effects on sustainability outcomes | Capability, skills and knowledge | - Professionalism and impact on profession - Deliver wider value and benefits |
| 37 | planners themselves have much to gain in developing their profession, strengthening its jurisdiction and showing greater leadership, in order for the demonstrated commitment to the concept of sustainability to act more clearly as a generative mechanism for sustainability practices. | Leadership | - Professionalism and impact on profession |
| 38 | 161 – local plans examined or submitted for examination since the introduction of the NPPF  105 - local plans found sound with 36% of LPAs boasting an up-to-date local plan against the NPPF 30% - of sound plans subject to an ‘early review’, all related at least in part to housing matters  71% - of early reviews subject to a deadline or time limit, with 8 local plan areas already missing that timescale  23 - tools and tests we’ve identified within the Housing White Paper that could help improve plan-making  56% - of LPAs likely to fall foul of the new housing delivery test in November 2017 and face a 20% buffer  3.6 % - aggregate undersupply of housing necessary to meet the household projections of housing requirements in all adopted Plans | Monitoring and evaluation | - Up to date local plan - Soundness |
| 38 | Five years on from the NPPF’s introduction of a “less complex” planning system, over 60% of LPAs are still without a Local Plan tested and found sound against national policy, with the majority of those still to get to the starting blocks of a local plan examination | - Impact of planning reform processes - Complexity of planning regulatory framework | - Up to date local plan - Soundness - Planning Inspectorate |
| 38 | Perhaps most notably, timescales for examining local plans have increased significantly. Before the introduction of the NPPF the average time from submission to a plan being found sound was 9.7 months. However, since the introduction of the NPPF, this has increased to an average of 16.8 months, with some taking upwards of three years to pass through the examination process | Local plan content and plan-making process | - Cost and delay - Planning Inspectorate |
| 38 | Almost half of plans have had to change their housing targets through the examination process, with the vast majority of those required to increase planned housing numbers, albeit a few plans more recently have seen reductions (Figure 4). (Further detailed findings, maps, tables and analysis throughout). | Local plan content and plan-making process | (Less) focus on process |
| 38 | As the NPPF hits half a decade of existence, the plan-led system it advocates – one that proactively seeks to meet needs – continues to show slow progress. Still fewer than four in ten local planning authorities have seen a local plan through examination | Impact of planning reform processes | - Up to date local plan - (Less) focus on process |
| 39 | 2018 | - Resourcing of local planning authorities - Capability, skills and knowledge |  |
| 39 | The analysis illustrates a stark picture of a planning system falling significantly short of delivering the levels of affordable housing required across the country. Combined with the outputs from other routes of delivery (for example grant funded schemes, and those led by housing associations), there is a major shortfall in the delivery of affordable housing.  ■ There is significant variation in outcomes on securing affordable housing across the country, between areas of high and low land values.  ■ Social rented homes – those available at the lowest levels – are not being delivered through the planning system in the vast majority of local authority areas | Local plan content and plan-making process | Impact on implementation |
| 39 | ■ Attendees suggested that there is a need for a spatial plan for England, with a regional approach to housing, including greater devolved powers. | Strength of national policy | Increase regulations/ powers |
| 39 | ■ Councils reported that greater powers are required in planning and place-making in the regions – the GMCA, for example, does not have the ability to call in planning applications, unlike the Mayor of London | Use of (local) planning powers | Increase regulations/ powers |
| 39 | There is an opportunity for councils to take a lead and act as a beacon for delivering high-quality affordable housing | Leadership |  |
| 39 | The survey results showed that the majority of councils (51%) have requirements in their Local Plans for social rented homes – those available at lowest rent for people in greatest need. There is a willingness and ambition among councils to secure these homes by requiring developers to contribute; however, the statistics show that they are clearly not being delivered in practice.  ■ Councils feel that the plan preparation stage is the most influential stage in determining whether or not affordable homes are secured through the planning process (38% said it was the most influential). | Local plan content and plan-making process | - Impact on implementation - Frontloading/ upstream |
| 39 | Around 70% of councils said that they rely substantially on the planning system to deliver the affordable homes that their area needs – indicating the current scale of reliance on planning. | Decision-making process | Impact on implementation |
| 40 | It was agreed by the delegates that the loss of social rent from the definition of affordable housing in the draft revised NPPF was particularly significant for the outcomes that could be secured from the planning system for those in greatest housing need, and they encouraged the TCPA to make this point in the Association’s response to the consultation | Strength of national policy | Clarity in national policy |
| 40 | ■ Council officers feel that planning is simply ‘tasked with too much’ and is being used, according to one officer, as a way of ‘government avoiding their responsibilities’ on investing in affordable housing. At the same time, councils said that as a country we are relying on the development industry to build the affordable homes we need when it is not within their business models to do so. | Resourcing of local planning authorities | Role of private sector |
| 40 | It was reported by councils that changes to permitted development that allow commercial properties to be switched to residential use without requiring full planning permission are having a significant and detrimental impact on the ability of councils to secure enough affordable housing | Impact of planning reform processes | Impact on implementation |
| 41 | Council officers all agreed that viability is the defining factor in deciding whether councils can secure affordable housing. Higher-value sites are much more likely to secure affordable housing than those of lower value. The result is that people in some areas ‘feel they are being left behind’. However, sometimes applications are received in high-value areas with low levels of affordable housing.  ■ Councils are increasingly looking at sophisticated approaches to viability, with viability assessments being an important skills base for emerging planners.  ■ One council officer said that the current policy framework plays into the hands of developers – and that the changes to the final NPPF are unlikely to make a difference to this. | - Impact of economic focus, viability and costs - Capability, skills and knowledge | - Innovative practices - Role of private sector - Commercial and development economics skills |
| 41 | Another council officer believed that national policy is unfocused and does not provide enough clarity on key terms to support councils | Strength of national policy | Clarity in national policy |
| 41 | Council officers believed that they need to able to have more local discretion in deciding what type and tenure of affordable housing is required. | Use of (local) planning powers | Increase regulations/ powers |
| 41 | Councils that have a strong institutional and corporate commitment to securing social and affordable housing, with dedicated staff resources, have greater levels of success | - Resourcing of local planning authorities - Visioning and outcomes-focused | Deliver wider value and benefits |
| 41 | Councils want to do more by themselves in delivering affordable housing – but the Housing Revenue Account borrowing cap and restrictions on the use of Right to Buy receipts have been handicapping them substantially | Use of (local) planning powers | Impact on implementation |
| 41 | ■ Council resources that are invested in negotiations with developers over contributions to affordable housing can be a major burden on wider services, according to one council | Resourcing of local planning authorities | Private sector investment |
| 42 | Many of the respondents to the survey and those interviewed spoke of the reliance that is placed on the planning system to deliver the affordable homes that are needed. This is problematic, especially because of the low levels of affordable housing that are being secured in many areas. Around 70% of the councils who responded to the survey said that they rely significantly on the planning system to deliver the affordable homes required in their area. One attendee at the expert roundtable agreed that planning is being relied on ‘massively’ at the expense of the delivery of other key infrastructure. As a result, councils agree that affordable housing is not seen as an integrated component of strategic planning in the present policy context, but rather as an output in its own right from the planning system and a target to be met | Decision-making process | Impact on implementation |
| 42 | Table 3 shows the proportion of affordable housing need met through planning policies in Local Plans. Importantly, Table 3 highlights the levels of ambition of councils in their Local Plan policies and the feasibility of delivering affordable housing need through planning. There is a huge variation in the extent to which the need for affordable housing is reflected in local planning policy. Analysis of individual Local Plans reveals more about the process of preparing these policies. Evidence explored in this report suggests that even though lower-value areas are generally more affordable, affordability is, nevertheless, still a major concern in low-value areas, and delivery is well below the levels required. Major investment in affordable housing is required in these lowerdemand areas to make up for this shortfall, alongside a planning system that prioritises the delivery of genuinely affordable housing above other outcomes | Local plan content and plan-making process | - |
| 42 | The responses to the project survey provide some important insights into the likely impact of the changes to the NPPF viability test made in the final revised NPPF and the updated PPG on viability. This is an important issue on which that the government has sought to provide clarification in the updated NPPF and PPG. In response, councils on the whole agreed with the principle that viability assessments ought to take place at the plan-making stage. However, they remain unconvinced that viability will not still be argued by developers at the planning application stages, with the wording in the guidance inviting an opportunity for viability to be re-assessed later on | - Impact of planning reform processes - Impact of economic focus, viability and costs - Strength of national policy | - Clarity in national policy - Impact on implementation |
| 43 | One of the attendees at the roundtable said that whether affordable housing is delivered to meet local need comes down to a matter of political will. One council interviewee noted a recent change in attitude at their local authority: whereas previously there was a lack of emphasis on the need for affordable housing, it is now seen as a priority. Another council representative said that ‘there is strong leadership from the council on driving forward housing growth, despite local opposition’. A change in the council’s approach came about as a result of it ‘previously being too reliant on the development industry, and a desire to set the bar higher in terms of quality’ | Leadership | Political commitment |
| 43 | There was consensus during one of the regional seminars that this is an exciting time to be in local government, with new and innovative approaches being taken | Knowledge transfer, good practices | Innovative practices |
| 43 | However, the TCPA research has found that many local authority planning departments are struggling with deregulation, demoralisation and a skills and capacity shortage.34 The cumulative impact on both policy and practice is a loss of confidence in planning for affordable housing. One council officer who was interviewed spoke about the significant resources required to negotiate and renegotiate Section 106 agreements, often for small numbers of affordable housing units, and the adverse impact on the morale of officers of working under a current policy framework that prioritises the needs of developers. | - Resourcing of local planning authorities - Impact of planning reform processes | - Confidence - Role of private sector |
| 43 | Despite the strain on local authority resources that securing affordable housing through the planning system entails, some councils have benefited from investing additional resources into teams that enable this delivery. This has helped to boost skills and provide for creative, entrepreneurial approaches | Knowledge transfer, good practices |  |
| 43 | A strategic programme approach has been taken with housing associations, committing to the longer term. This has fostered good relationships with the housing association sector and allowed flexibility over how developer contributions are spent. The council has also taken the time to create a good working relationship with Homes England and is aiming to adopt a collaborative approach as far as possible to meet local housing need by bringing in a range of partners | - Partnership, collaboration, relationships, engagement - Stakeholders and statutory consultees | Innovative practices  Role of private sector (RSL) |
| 44 | 2021 | - Resourcing of local planning authorities - Capability, skills and knowledge |  |
| 44 | National planning policy has been characterised by stop-start reform over several years. This has regrettably resulted in uncertainty among local authorities and across the planning sector. Contrary to the Government’s objective of facilitating planmaking, the short-term effect of its announcement of proposed planning reforms (6 December 2022) has been to halt the progress of local plans in a number of local authority areas. (Paragraph 14)  2. The Government must see the merit in pausing plans for further reform, in order to allow for a period of stability in which reforms already introduced can be properly implemented, and any lessons from that implementation learned. (Paragraph 15)  3. The Government has not been clear on the timetable for its many planning consultations and when its reforms will be implemented. Nor has the Government sufficiently evaluated the impact of its past NPPF changes to inform its current reform proposals. There is a strong case that the Department should conduct impact assessments of past NPPF changes, which would inform future reform proposals. Given that the Department is currently considering 26,000 responses to the December 2022 consultation, and is conducting at least nine further consultations on planning reform, we do not believe resource constraints should prevent the Department from conducting these impact assessments. (Paragraph 21) | - Impact of planning reform processes - Use of impact assessment | - Uncertainty - Impact on implementation - Cost and delay - Up to date local plan |
| 44 | The Government should urgently conduct and publish impact assessments on all future NPPF changes. It should take a more strategic approach to future consultations, including publishing timelines for the implementation of its proposed reforms. (Paragraph 22). | Use of impact assessment | - |
| 44 | We support the principle of a plan-led system and are sympathetic to the Government’s wish to ensure more local authorities have up-to-date local plans. However, it is difficult to see how the Government will achieve its 300,000 net national housing target by the mid-2020s if local targets are only advisory. The Government has not provided sufficient evidence to demonstrate how the policy of removing mandatory local housing targets will directly lead to more housebuilding. (Paragraph 33 | Strength of national policy | - Up to date local plan - Clarity in national policy |
| 44 | 7. In line with its previous commitment to us, the Government must publish its own comprehensive analysis, as part of its response to the December 2022 consultation, to demonstrate how the proposed changes to the NPPF will facilitate delivering 300,000 net new homes per year, including the evidence base for each of those proposed changes. The response to the December 2022 consultation containing this analysis should be produced by the end of September 2023; it was originally expected in spring 2023. If there are further delays, the Minister for Housing and Planning should write to us to explain why. (Paragraph 35) | Evidence base | - Access to evidence base - Cost and delay |
| 44 | 9. If the NPPF reforms, once they have been implemented, do result in a reduction in housebuilding, the Minister for Housing and Planning should write to us as soon as this becomes apparent, and should explain whether the Government intends to keep the national housing target by making further NPPF revisions, or maintain its policy of advisory local targets at the expense of building 300,000 net new homes per year, or take other action in response. (Paragraph 37) | Impact of planning reform processes | - Impact on implementation - Clarity in national policy |
| 44 | Local planning authority resourcing  17. There continues to be a pressing need for additional resources for local planning authorities to ensure the efficient working of the planning system and to implement the Government’s proposed reforms. The Government must ensure local planning authorities have the specialist skills required to implement proposed reforms. The programme of support offered by the Department—including the measures outlined in correspondence from the Minister for Housing and Planning, and the letter from the Chief Planner to local authorities—does not constitute a comprehensive resources and skills strategy for the planning sector. This does not match the scale of the resourcing challenge which local planning authorities currently face. (Paragraph 79)  18. The Government should publish a comprehensive resources and skills strategy for the planning sector, in line with its commitment to us. The strategy should clearly explain how the resourcing and skill needs of local planning authorities will be met; and should be published before future reforms to national planning policy are implemented. (Paragraph 80). | - Resourcing of local planning authorities - Capability, skills and knowledge | Impact on implementation |
| 45 | 2012 | - Capability, skills and knowledge - Resourcing of local planning authorities |  |
| 45 | Planning regulations were perceived by some authors to be inflexible | - Complexity of planning regulatory framework - Strength of national policy | Health considerations |
| 45 | Concerns were also raised about gaps in the quality and range of the local evidence base supposed to underpin the ‘soundness’ of plans and allowing planning permission | Evidence base | - Access to evidence base - Soundness - Impact on implementation |
| 45 | as well as inadequate scoping processes in plans, resulting in the exclusion of health and well-being as objectives. Health outcomes are rarely used as grounds for refusing planning permission (NICE, 2011). | - Decision-making processes - Use of impact assessment | - Health considerations - Impact on implementation |
| 45 | Our literature review analysis suggests that those responsible for decisions on, and assessments of, planning proposals often view health in narrow terms, focussing on physical environment concerns such as air quality, rather than recognising the role of the social environment and other broader determinants of health. This narrow focus is seen to be primarily a result of a lack of engagement between health and planning professionals, coupled with the rigid boundaries around the development of knowledge between these two professions, different cultures between the various stakeholders, with differing terminologies and languages, priorities and structures (UWE, 2011c). Lack of understanding of the roles that different organisations and individuals hold is also factor in that respect (Tewdwr-Jones, 2011). Furthermore case study research in England shows that spatial planners have a weak knowledge of how they can influence the determinants of health. | Capability, skills and knowledge | - Professionalism and impact on profession - Governance - Professional qualification |
| 45 | Facilitators of good practice Good practice occurs when the health sector takes a pro-active approach to development planning and partnering with local planners. Where partnership exists, it is more advanced in relation to the planning of healthcare facilities – health professionals often do not yet see planning as a core business (Tewdwr-Jones, 2011; NICE, 2011). A number of good practice examples have nevertheless emerged involving the use of “broker” agency such as the London’s Healthy Urban Development Unit (HUDU) advising local authorities and health agencies on the planning of health facilities through the use of legal planning agreements, extracting financial support from developers and bridging the divide between the two sectors. | Knowledge transfer, good practices | - Innovative practices - Better use of local levers - Private sector investment |
| 45 | The Bristol and Plymouth case studies stress the value of joint appointments between health authority and local authority. This has been found to break down silo barriers and greatly assist the integration of health into planning policy and decisions. It can take the form of a jointly appointed director of public health, and a dedicated officer with explicit health and planning responsibilities. There are also real benefits if the health authorities are engaged in the process of plan-making at an early stage, so as to influence the core agenda of the plan. According to the Manchester, Bristol as well as Plymouth case studies, this should also apply to major developments: the public health authority can influence the nature of the initial advice given to applicants. Changes to move the public health function into local authorities in England may be beneficial in this respect. | Capability, skills and knowledge | - Dedicated posts and capacity - Frontloading/ upstream - Impact of public health reforms |
| 45 | Further research is needed to examine how the rhetoric of plans and strategies is implemented on the ground, how planning and other departments prioritise health when defining the purpose and scope of plans and projects. The appraisal processes, in this regard, should be seen as integral to the whole decision-making process and ensure health objectives help shape the options that are considered. We found in Manchester, Plymouth and Bristol that a pre-application HIA, with the health and planning authorities helping with scoping, can enable key issues to be addressed in advance and mitigation incorporated at the outset when it is likely to be much more effective. HIA has been identified as a trigger for mutual learning. | - Decision-making processes - Use of impact assessment - Training, learning and development | - Impact on implementation - Uncertainty - Health consideration - Health impact assessment |
| 45 | The English planning system is at a turning point. Our assessment occurred as the Government was about to radically reform the system and increase the planning remit of local authorities and neighbourhood planning within a broader “Localism” agenda including the simplification of the raft of planning policy statements and guidance into a single document, the National Planning Policy Framework (NPPF). It will be interesting, in time, to examine whether the new system offers better opportunities for planners to consider health outcomes of their decisions or if it will prevent progress being made. | Impact of planning reform processes | - |
| 46 | At the time we carried out our research, the English planning system did not contain any specific planning policy guidance or planning policy statement on health. It has some excellent nonstatutory healthy environment guidance in the fields of urban design, sustainable building design, local transport and street design, open space, green-space and recreation, for instance By Design (DETR and CABE, 2000), the Manual for Streets (DfT, 2007) or the Code for sustainable homes (DCLG, 2009), but the potential health benefits are not always sufficiently explicit and major gaps in official guidance (except at the broad policy level) occur in relation to, for example, accessibility, social inclusion and strategic policy. This is likely to reduce the ability of local authorities to plan healthy urban environments. | Strength of national policy | - Clarity in national policy - Impact on implementation |
| 46 | On one hand, as the UK government promotes more ‘localism’ it might be argued that national guidelines and guidance are no longer appropriate. Even before the new UK 2010 Coalition Government, UK planning regulations required evidence backing standards to be locally based where possible. On the other hand, while Building Regulations and traffic design requirements remain centrally defined, in some other spheres documents recommend levels and set ‘benchmarks’, or specify a process, rather than statutory obligations. So there is still very much a place for national guidelines. | Strength of national policy | - Increase regulations/ powers - Clarity in national policy |
| 46 | This lack of policy guidance or planning policy statement on health has been, to an extent, corrected. The 2012 NPPF defines the key term “sustainable development” (which is intended to be the guiding principle of planning policy) by specifying economic, social and environmental dimensions. The social dimension puts health and well-being centre stage. This at last removes the excuse for inaction by some practicing planners, that health is not a material consideration in planning decisions. The NPPF does, though, lack precision on how to interpret healthy planning and how to monitor achievements on the ground. The National Indicators (used until 2011) to assess and compare local authority performance, included targets relevant to health and well-being. Although they have been abolished, it could be argued that they were useful tools to promote healthy environments, and had important health implications. | - Strength of national policy - Monitoring and evaluation - Impact of planning reform processes | - Clarity in national policy - Health considerations - Impact on implementation |
| 46 | Finally, the new NPPF suggests more joint working between public health and planning, an issue that clearly defined as key in the promotion of healthy planning both by our evidence review and case study research. Local planning authorities when drawing their development plans should work with public health leads and health organisations to understand and take account of the health status and needs of the local population (such as for sports, recreation and places of worship), including expected future changes, and any information about relevant barriers to improving health and wellbeing (CLG, 2012). The move of the public health function into local authorities in 2013 in England offers new opportunities in this respect (DoH, 2010). It will be interesting in time to examine if this policy ambition has the desired results in producing healthier environments | Partnership, collaboration, relationships, engagement | - Impact of public health reforms - Impact on implementation |
| 46 | In addition, the level of integration of health into plans in England depends not so much on the planning system per se as on the leadership, commitment and knowledge of politicians and practitioners involved | - Leadership - Capability, skills and knowledge | - Role of elected members - Political commitment |
| 46 | As our research showed further, the barriers to health integration are organisational and professional silos, ignorance, resources, and a reactive planning regime | - Resourcing of local planning authorities - Complexity of planning regulatory framework - Capability, skills and knowledge | - Governance - Professionalism and impact on profession |
| 47 | 2022 | Capability, skills and knowledge  Resourcing of local planning authorities |  |
| 47 | The findings of this research indicate that the English planning system continues to produce the socially conservative outcomes repeatedly identified in research over the past several decades. It is evident that tackling racial inequalities in housing and meeting the accommodation needs of BAME households are not explicit aims of planning at present, nor are these issues being considered in practice. While in most case study areas planning authorities did consider the needs of ethnic or faith groups in their SHMAs, this did not translate into specific policy aimed at addressing this need. This was due in part to a feeling that planning departments must not be seen to be prioritising the needs of certain groups over others. This naïve approach to ‘equalities’ (i.e., focussing on equality of treatment rather than equality of outcomes) has been identified and challenged by planning researchers for over 40 years, and will continue to reinforce existing disparities and discrimination within planning processes if it remains unchanged. | Visioning and outcomes-focused | - Deliver wider value and benefits - Clarity in national policy - Lack of alignment - Health consideration (equalities) |
| 47 | It must be acknowledged that there are some factors associated with racial inequalities in housing which local planning authorities simply struggle to influence. Firstly, it is difficult for planning departments to improve poor-quality existing housing stock which is often inhabited by BAME families due to the cheaper costs associated with it. Secondly, the placement of new housing developments remains reliant on land being or becoming available in suitable locations. Finally, local planning authorities cannot ultimately control whether the requested numbers of affordable housing will be accepted following a viability assessment, and whether any affordable housing that is delivered is genuinely affordable for the average resident. | Impact of economic focus, viability and costs | Cost and delay |
| 47 | However, despite these limitations, the planning system is a critical tool in addressing racial inequalities in housing as it is the key mechanism for delivering new homes in England and has significant untapped potential in tackling this form of social injustice. It does not follow from the lack of progress on these issues to date that the planning system cannot play a substantial role in addressing social injustices impacting BAME groups in the future, and any improvements made in pursuit of this aim could add real value in an effort to tackle such issues holistically across multiple policy areas. | Visioning and outcomes-focused | Deliver wider value and benefit |
| 47 | To begin with, the National Planning Policy Framework (NPPF) should be updated to include a core focus on tackling racial inequalities and meeting the needs of BAME groups; having this central requirement running through all planning policy would compel local authorities to better address these aims in every aspect of decision-making. This would therefore help to prevent equalities considerations from remaining ‘tick-box’ exercises and would put greater emphasis back onto ‘who’ planning decisions, particularly relating to housing, really affect. | - Strength of national policy | - Clarity in national policy - Impact on implementation - Health consideration (equalities) |
| 47 | Existing tools such as equality impact assessments (EqIAs), strategic housing market assessments (SHMAs) and public consultation requirements could be used more explicitly to pursue the goal of meeting the accommodation needs of BAME households. This would in part depend on Government issuing more robust guidance and/or directives on how local authorities could or should use these tools with this aim in mind, and also on local planning authorities having the staff, skills and resources to collect accurate data on these accommodation needs. EqIAs should be coupled with a statutory requirement to take account of any findings felt to have adverse impacts on any groups with protected characteristics. | - Use of impact assessment - Resourcing of local planning authorities - Monitoring and evaluation | - Clarity in national policy - Better use of local levers - Health consideration (equalities) - Access to evidence base |
| 48 | Finally, public consultation opportunities within planning urgently require improvement and refocussing to include a more diverse range of voices within decision-making processes. Worryingly, it seems that the manner in which consultation processes work at present often reinforces rather than challenges existing power inequalities. In particular, BAME groups and others who are marginalised or on low incomes seem much less likely to engage as they can often lack the spare time to devote to participating, an awareness of how to get involved, or the specialised knowledge needed to understand documents that include language specific to planning | Stakeholders and statutory consultees | - Role of local communities - (Less) focus on process - Externalised expertise - Lack of engagement |
| 48 | Increasing engagement is likely to require dedicated and properly resourced outreach work. Examples were given of how diversity of respondents has increased since consultation processes were held online during the pandemic. In Bradford, consultation processes including local housing providers, BME housing specialists and community faith leaders had positive outcomes as they led to a reference to cultural housing needs being included within the local authority’s housing strategy, and the design of regeneration projects being changed to better meet such cultural needs. Therefore, it is possible for stakeholders to have meaningful involvement in consultation processes if leadership is shown on this issue. | - Capability, skills and knowledge - Partnership, collaboration, relationships, engagement | - Lack of engagement - (Less) focus on process - Better use of local levers - Innovative practices |
| 47 | Mainstream equalities considerations throughout any proposed planning reforms. This would compel local authorities to take account of issues of racial equality in all planning decision-making and therefore prevent these considerations from being simply a tick-box exercise | - Strength of national policy - Decision-making | - Clarity in national policy - Impact on implementation |
| 47 | Expand the National Planning Policy Framework’s presumption in favour of sustainable development to include an aim of striving for racial equality in all planning processes, so that this overarching aim can feed into all aspects of the planning system | Strength of national policy | Clarity in national policy |
| 47 | Resource local planning authorities to keep up-to-date records of housing needs in their area and to prevent a reliance on outdated records in between censuses. | Evidence base | Access to evidence base |
| 47 | Attach a clearer statutory duty to equality impact assessments (EqIAs) that obligates local authorities to act on any findings that reveal adverse impacts on groups with protected characteristics | Use of impact assessment | Increase regulations/ powers |
| 47 | Include specific information of the needs of ethnic and/or faith groups when conducting a strategic housing market assessment (SHMA). By doing so, planning departments would develop a better understanding of the housing needs of their BAME residents when completing their SHMA and this could then provide the evidence needed to pursue a specific focus on the housing needs of BAME groups in planning policy | Evidence base | Increase regulations/ powers |
| 47 | Actively undertake outreach work in order to ensure that the views of BAME communities are included in public consultation opportunities, including producing documents in different languages, and liaising with established community leaders who can act as mediators between the council and the wider community. Whilst this may serve as a good starting point for increasing engagement, councils should not assume that this will be sufficient to ensure all community views are represented; efforts should also be made to promote the inclusion of more marginalised members of BAME communities, such as children and young people, women, and low-income or homeless households. • Continue to utilise new online forms of consultation, which have led to increases in public engagement during the Coronavirus pandemic, whilst also maintaining an awareness of who may be affected by the wider issue of digital exclusion. | - Partnership, collaboration, relationships, engagement - Leadership | - Lack of engagement - Role of local communities - (Less) focus on process - Better use of local levers - Access to evidence base |
| 47 | Work in partnership with any specialist BME housing providers operating locally, and also any housing providers known to house large numbers of BAME or low-income residents, to foster mutual learning and to share data and information on the needs of BAME groups | - Knowledge transfer, good practices - Partnership, collaboration, relationships, engagement |  |
| 47 | • The Royal Town Planning Institute (RTPI) should continue to pursue their “Change” Action Plan3 to increase equality, diversity, and inclusivity in the sector, and should provide guidance for local authorities detailing how they too can increase diversity in the planning profession. This should go beyond junior or entry level roles and seek to ensure that both diversity and the skills needed to take real account of equalities issues are also increased in senior roles and positions of leadership. | - Capability, skills and knowledge - Leadership | - Professionalism and impact on profession - Professional qualification |
| 48 | The RTPI should work with external stakeholders in housing, race equalities, and academia to develop educational resources on ‘race and planning’ which can be accessed by local authorities and planning professionals | - Training, learning and development | - Externalised expertise - Professional qualification |
| 48 | Ensure that planning degree programmes includes teaching on how race equality and other social considerations are relevant to the study of planning, including education on the limits of formal equality of treatment in addressing systemic inequality and disadvantage. • Make efforts to raise the profile of the planning profession amongst a diverse range of potential students from BAME and low-income backgrounds, e.g. by providing targeted bursaries or financial support | Training, learning and development | - Professionalism and impact on profession - Professional qualification - Funding |
| 49 | 2016 | Resourcing of local planning authorities |  |
| 49 | Problems Facing Plan Preparation  S6. We heard an almost unanimous consensus that the principal difficulties affecting plan making are attributed to the following matters, in approximately this order:- • agreeing housing needs; • difficulties with the Duty to Cooperate, including the distribution of unmet housing needs; • a lack of political will and commitment; • a lack of clarity on key issues, particularly SHMAs, strategic planning, Green Belt and environmental constraints; • too many changes – changes of policy, advice and factual changes in forecasts (“moving the goalposts”); | - Impact of strategic planning practices - Impact of politics and politisation in planning - Impact of planning reform processes | - Political commitment - Clarity in national policy - Consistency - Lack of alignment |
| 49 | S7. In addition we heard of concerns with the extent of evidence base requirements, the nature of the examination process, the soundness tests and the consistency of decision making from the Planning Inspectorate | - Evidence base - Decision-making processes | - Access to evidence base - (Less) focus on process - Consistency - Planning Inspectorate |
| 49 | S11. Serious problems are generated by the lack of an agreed approach to SHMAs, which have become one of the most burdensome, complex and controversial components of plan making. We set out detailed recommendations for a shorter, simplified, standard methodology for SHMAs and, in particular for assessing housing need, with the aim of saving very significant time, money and, most importantly, with the intention of removing unnecessary debate from this aspect of plan making. | - Local plan content and plan-making process - Complexity of planning regulatory framework | - Lack of alignment - Cost and delay |
| 49 | S13. Despite the clear test set by paragraph 14 of the NPPF, few authorities compile an assessment of the environmental capacity of their area, making it difficult for Planning Inspectors to apply the NPPF policy in situations where authorities propose to provide fewer homes than the assessed level of need. Whilst we recommend significant reductions in other elements of the local plan evidence base (see below), we propose that a proportionate Assessment of Environmental Capacity should be an important part of plan making. The NPPG should be strengthened to ensure a robust application of the NPPF’s expectation that needs will be met unless the authority can demonstrate that to do so will cause significant adverse effects.  S14. We recognise that the NPPF does not require authorities to meet the full identified need for development in all circumstances, even within the Housing Market Area, if there is insufficient environmental capacity but we encountered significant uncertainty about how the appropriate balance should be struck. We make recommendations to remove that uncertainty and to confirm the legitimacy of applying the tests set out in the NPPF to ensure that needs are met up to the point where the adverse effects of doing so can be shown to outweigh the benefit of meeting the need. | - Evidence base - Strength of national policy | - Type of evidence base - Clarity in national policy - Uncertainty |
| 50 | S16. We received strong representations that the Duty to Cooperate was not effective in ensuring agreement between neighbouring authorities about the distribution of housing needs and that this was one of the most significant constraints to effective plan making. Whilst the NPPF is already clear that it expects the outcome of the Duty to be that housing needs will be met, it is apparent that further measures are necessary.  S17. In order to provide more “bite” our recommendations include revisions to the soundness tests of the NPPF to emphasise the expectation that needs should be met, with authorities who do not plan to meet their own needs identifying how they expect those needs to be satisfied elsewhere. This would necessarily involve authorities applying to their neighbours to meet their unmet needs and, where necessary, engaging in representations on their neighbours’ plans in order to test the capacity of those adjacent authorities to meet unmet needs. Revisions to both the NPPF and the NPPG are necessary for this purpose.  S18. Even with that strengthened position in place, however, local plans are rarely coordinated in time and, whilst the Duty to Cooperate may encourage joint working between pairs of authorities, it is not sufficient in itself to generate strategic planning across wider areas, such as housing market areas. | - Impact of strategic planning practices | - Soundness - Clarity in national policy - Lack of engagement - Lack of alignment |
| 50 | S20. There are several examples of good joint working between authorities and we agree with the RTPI, District Councils Network and others that such joint working should be encouraged. In a number of parts of the country, however, joint working needs to be actively facilitated if housing needs are to be effectively addressed – in the meantime, significant shortfalls in delivery will continue. We recommend that, where authorities have failed to reach sufficient agreement on meeting and distributing housing needs by March 2017, the Government should take and use powers to direct the preparation of a high level Joint Local Plan for the HMA or a suitable geography, such as transport corridors, within a prescribed timetable | - Impact of strategic planning practices - Partnership, collaboration, relationships, engagement | Increase regulations/ powers |
| 50 | S21. From the outset of our appointment, LPEG has been interested in the potential for voluntary joint planning provided by the current round of bids for devolved powers, which cover a large majority of the country. Devolution provides the best opportunity for bottom-up joint planning but bids tend to focus on economic growth rather than housing and we have strongly recommended to Government that it attaches precise conditions to any successful devolution bid, requiring a commitment to plan positively to meet objectively assessed housing needs and a commitment to produce a plan for the combined area. We further recommend that individual authorities within a combined authority area should receive sign off from the combined authority that their emerging plan addresses the Duty to Cooperate before their plan can progress. | Use of (local) planning powers | - Increase regulations/ powers - Better use of local levers |
| 50 | Incentives for Timely Plan Preparation  S22. Many respondents suggested that it is necessary to use one of several “carrots and sticks” to incentivise local authorities to generate timely local plans or local plan reviews. Characteristically, public sector respondents were more likely to suggest that good performance should be rewarded, whilst private sector representatives were more likely to suggest that poor performance should be penalised. We recommend that Government should review the role of financial incentives to stimulate efficient and effective plan making, although we recognise that those matters are already under review and we have concentrated instead on policy and procedural incentives. Authorities bidding for funds should expect to have an up to date plan in place.  S23. In relation to the Government’s recent consultation on sharpening the incentive provided by the New Homes Bonus to encourage plan preparation, we have responded to the consultation setting out strong views that any incentive should not inadvertently condone the poor plan making performance of authorities who have not put in place a post NPPF compliant local plan.  S24. In addition to the Government’s proposals that it may intervene to arrange for local plans to be written in consultation with local people, we recommend that the stimulating effect of that announcement can be enhanced by: Introducing a statutory duty on local authorities to produce and maintain an up to date local plan | Local plan content and plan-making process | - Up to date local plan - (Less) focus on process - Cost and delay - Soundness - Increase regulations/ powers |
| 51 | Policy Changes  S27. Plan making is significantly affected by changes in Government policy. The NPPF sought to bring clarity to Government policy and to end “policy creep”; however, recent years have seen a large number of additional policy statements, new legislation and rapid changes to the NPPG, all of which may be beneficial in their own right but which do have the unintended consequences of destabilising plan making, which needs a solid foundation | - Complexity of planning regulatory framework - Impact of planning reform processes | - Clarity in national policy - Uncertainty - Risk management - Impact on implementation |
| 51 | S28. Planning now needs a period of stability after significant recent reform and we recommend that:- • the NPPF is reviewed only every five years; • the NPPG is only changed periodically (for instance, every 6 months); and • proposed changes to the NPPG are subject to scrutiny by a technical working group before the changes are made in order to reduce the prospect of unintended consequences. | Impact of planning reform processes |  |
| 51 | Local Plan Process  S29. Our work identified a fault in the current local plan making process. At present, the Local Plans Regulations do not allow a local authority to modify a plan in response to public consultation at the first (and only) stage when a local plan is formally published in draft. This creates several difficulties: • local communities feel excluded from the plan making process; • plans may be at risk because consultation may not meet legal requirements; and • as a result, many authorities undertake additional non-statutory stages of consultation, thereby adding significantly to the plan making process. We recommend two particular changes to the current regulations so that: • the first stage of engagement (Regulation 18) should principally enable the community to express their views about their vision for the area and their views on all relevant issues; and • a local authority can change its published plan in response to public consultation without undertaking a further round of plan making.  S30. These simple changes would substantially improve community engagement but also significantly speed plan making.  S31. We have identified a series of further measures which would reduce a number of the more burdensome and unnecessary obligations on the plan maker including:- • a significantly shorter, standardised approach to calculating housing needs (see above); • a much tighter definition of evidence which it is necessary to gather in preparing the plan – limiting evidence to that which is strictly necessary to meet legal requirements; • clear advice that the preparation of a simple Sustainability Statement auditing the local plan against the NPPF would be sufficient to meet the legal requirement for Sustainability Appraisal – thereby dramatically reducing the burden of one of the most time consuming aspects of plan making; • scoping back local plans so that they deal only with strategic issues which cannot be addressed in Neighbourhood Plans and other documents; and • amending the soundness tests so that the local plan need only be “an appropriate strategy”, thereby providing much greater local control over the vision and strategy within the local plan. | - Local plan content and plan-making process - Evidence base - Use of impact assessment | - Role of local communities - Risk management - (Less) focus on process - Cost and delay - Type of evidence base - Soundness |
| 52 | Local Plan Content  S35. Whilst local plans could undoubtedly be shorter (probably around 50 pages) we do not recommend that this is regulated; neither do we recommend the use of template policies. Each local plan should be a distinctive view from its community about the future of its area.  S36. Our appendices draw together (for the first time) a list of requirements for a local plan and a list of the necessary evidence base, to assist plan makers. We identify the scope for a proportionate approach to both. We also provide advice and recommendations that would limit the scope of local plans to strategic issues, thereby creating a clear role for Neighbourhood Plans or (if neighbourhood planning is inactive in an area) for secondary local plan documents and brownfield registers. We also recommend a change to the NPPF to make clear that it is not necessary for authorities to prepare a single local plan, which covers every issue – rather, staged local plans, which defer local issues to Neighbourhood Plans would be appropriate.  S37. Similarly, local plans do not need to repeat development management policies set out in the NPPF. Instead, we recommend a simple formula of words, which can incorporate those policies without repetition. We make detailed recommendations in relation to the way in which local plans can be structured and the way in which they deal with infrastructure and their interface with the Community Infrastructure Levy.  S38. Importantly, however, we particularly recommend that local plans must generate the confidence that they are planning sustainability over the full local plan period (at least 15 years). The simplification of housing issues means that local plans should also engage with those matters of greatest concern to local communities including biodiversity, heritage, place making and quality of life. | Local plan content and plan-making process | - Type of evidence base - Confidence - Deliver wider value and benefits |
| 53 | Presentation, Access and Style  S43. Our report reviews emerging best practice in local plan production, noting that there is no central resource with which plan making authorities can consult.  S44. Recommendations are made for shorter, more publicly accessible plans and for the better use of online technology, with best practice examples highlighted. This includes the approach known as “propositional planning” in which communities are engaged with clear illustrations of the consequences of proposed plans in order to stimulate easier engagement. | Knowledge transfer, good practices | - Innovative practices - Externalised expertise - Role of local communities - Lack of engagement |
| 49 | 29. PINS resources. We recommend that Government undertakes a review of PINs resources in the light of the full scale of recommendations set out in this Report. | - Stakeholders and statutory consultees - Resourcing of local planning authorities | Planning Inspectorate |
| 54 | 2023 | Resourcing of local planning authorities |  |
| 54 | a) The increase in case law and precedents: Over time, challenges and appeals have led to an inflated set of parameters and decisions, each providing new risks of appeal and judicial review of decisions. Under pressure, local planning authorities therefore request more evidence up front at the outline stage ‘as standard’ in anticipation of any potential legal challenge based on former decisions | - Evidence base - Decision-making processes | - Risk management - Planning Inspectorate - Access to evidence base |
| 54 | b) The increased politicisation of planning: Our interviewees agreed that, compared with the 1990s, there is a much higher level of public scrutiny and - on occasion – antipathy to any development in certain areas. This means that planning officers and councillors often ask developers for more evidence, in anticipation of a potential objection, at a much earlier stage than used to be the case. | - Impact of politics and politisation in planning | - Use of evidence base - Frontloading/ upstream |
| 54 | c) The lack of public sector capacity across the decision-making process. The reduction in funding across local planning authorities has been well ventilated, and with it there has been a lack of capacity especially in more specialist roles and a significant turnover in staff. This means that applicants are often forced to consistently ‘start again’ on demonstrating the case for development, often in the face of inconsistent feedback on proposals, leading to delays which can overwhelm the project timeline. | - Resourcing of local planning authorities - Decision making process | - Dedicated posts and capacity (Specialist) - Consistency - Cost and delay |
| 54 | 1) Outline planning permissions needs to return to being outline, rather than being only a nudge down from a detailed approval. The evidence required at the outline permission stage should be simplified and scaled back commensurate with the ‘principle of development’ on the site rather than the detail, allowing for other – legitimate – considerations to be addressed at the detailed stage. Whilst this will not be a return to the simple red line and form approach of the 1980s, this can help reduce the costs and potential barriers to entry that the current burdens provide. | Evidence base | - Type of evidence - (Less) focus on process - Cost and delay - Impact on implementation |
| 54 | 3) Reducing or eliminating fees for smaller sites (of between 10 and 100 units) would reduce the barriers to entry for SME builders to compete on small sites. Ringfencing, or partially ringfencing the remaining planning fees, can help LPAs build their capacity and can ensure decision making incentives are aligned with the Governments ambition to build 300,000 homes each year, as well as addressing ongoing workforce issues in planning teams. | Resourcing of local planning authorities | Planning fees |
| 54 | 4) LPAs need to work with consultees and committee members to ensure that expectations for outline permissions are understood. This could be a practical guide, backed by professional planning organisations, in which consultees and committee members are given the guidance and information needed so they can assess outline permissions more effectively. This would be aimed at reducing the amount of evidence expected at the outline planning stage. | Stakeholders and statutory consultees | - Role of elected members - Externalised expertise - Use of evidence base |
| 55 | 2023 | Resourcing of local planning authorities |  |
| 55 | 3.1. Competing priorities  A diversity of actors are involved in urban development decision-making. We identified competing priorities amongst three key groups: national government (including politicians, civil servants and government agencies); local/regional government (including councillors, officers, and regional bodies); and private sector for property development (including developers, investors, landowners and brokers). Their dominant values and intentions may result in either aligned or conflicting priorities which can influence health outcomes. | - Complexity of planning regulatory framework - Stakeholders and statutory consultees | - Deliver wider value and benefits - Lack of alignment - External influence |
| 55 | .1.1. National government priorities  National government actors had multiple, diverse priorities spanning housing, economy, environment, public opinion and political ideology. Trade-offs that affect health considerations relating to urban development appeared inevitable. | Complexity of planning regulatory framework | - Health considerations - Lack of alignment |
| 55 | Conservative, libertarian ideology was seen as influential in the housing market, where home ownership was promoted over renting, and government had a limited role in providing social housing or funding place-making infrastructure. There was a view that “we need very brave politicians” (AB-250, Scientific advisor) to support healthy development | Impact of politics and politisation in planning | - Political commitment - Health considerations |
| 55 | 3.1.2. Local/regional government priorities  Local/regional government also had multiple, often competing, demands that influenced decisions associated with healthy urban development. These could result in trade-offs involving quantity and quality of housing, social needs, financial demands, public opinion and political concerns. | Complexity of planning regulatory framework | - Lack of alignment - Funding |
| 56 | Financial limitations were discussed as barriers to achieving all desired development outcomes by politicians and officers – it appeared that local government officers had too many issues deemed to be priorities by leaders, without adequate acknowledgement of necessary trade-offs, e.g. commercial, social and political decisions associated with affordable housing could be in tension, especially with limited funding.  Because of reductions in central government funding, and multiple short-term pressures, there was a feeling that “local authorities are now just completely dependent on their business rates and their council tax” (UB-468, local government, public health), therefore development was accepted to secure investment not available from internal budgets (as well as achieving housing targets). | Resourcing of local planning authorities | - Funding - Lack of alignment |
| 56 | Local government agenda, led by politicians, could influence urban development. However, there was reluctance to take potentially controversial stances, particularly for challenging car dominant environments. Community engagement for active transport interventions was described as ‘activation’ for ‘promoting’ active neighbourhoods, with recognition that cultural change takes time. Tension was perceived between neighbouring councils with different priorities, with suggestions that councillors may not care about the health impacts of urban development for residents living outside their administrative area. | Impact of politics and politisation in planning | - Lack of alignment - Lack of engagement - Health considerations |
| 56 | Health was often not expressed as a priority but there was evidence of strong support for healthy place-making by local government officers. It just may not be framed directly as ‘health’, rather “It tends to be talked about in terms of carbon, air quality, congestion externalities rather than health.” (HI-491, regional government, environment and transport). | Visioning and outcomes-focused | Health considerations |
| 56 | 3.1.3. Private sector priorities  Private sector priorities tended to focus on maximising profits, which could conflict with healthy urban development outcomes. However it appeared that some elements of the market (e.g. long-term and institutional investors) were increasingly needing to consider sustainability and wellbeing issues. Developers’ investors for real estate operate in global financial markets and seek “a reasonable risk-adjusted return” on investment (DL-152, private sector, real estate finance).  Seeking short term profit can reduce incentives to create healthy environments which may be more expensive. Prioritising profit may promote gaming behaviours such as land banking or flipping. Investors' interest in health appeared mixed: some property developers expressed that “[investment bankers don't] give a damn about place making or really they don't give two hoots about people's health. They want return on their investment” (MF-177, private sector, property development). However, various interviewees from global financial services companies highlighted that real estate investors, especially institutional investors such as pension funds, increasingly see sustainability and wellness as critical factors that affect their return on investment and consider them very much in their investment decisions in this shareholder society | Impact of economic focus, viability and costs | - Role of private sector - Private sector investment - Health considerations - Deliver wider value and benefits - Risk management - External influence |
| 57 | 3.2.1. ‘Rules’: policies and legislation  Many centralised policies, regulations and laws were discussed that controlled urban development decision-making, alongside local planning policies and frameworks. However, these may be insufficient to enable healthy place-making due to focus on process over outcomes, power asymmetries, perverse financial incentives and lack of enforcement.  Some stakeholders discussed how ‘rules’ sought to control private sector developers who may otherwise produce poor quality developments, but there was criticism of focus on process rather than outcomes. | - Visioning and outcomes-focused - Strength of national policy | - (Less) focus on process - Impact on implementation |
| 57 | Regulations that defined minimum standards were criticised because they were insufficient to ensure healthy urban development. They reportedly could exacerbate inequality as only wealthier places were able to negotiate higher standards.  Some interviewees suggested that regulation, taxation or policy change was needed e.g. to price in health and environment costs upfront; or for finance and credit regulations to influence housing affordability and accountability to stakeholders. Levelling up differences in tax arrangements on home ownership and rental markets, as well as on VAT subsidies for new housing developments, so they aligned with retrofitting, was also suggested. However some thought that regulations were sufficient – they thought implementation of regulations were the problem.  Despite limitations of ‘rules’ to enable healthy development the law was perceived as an important tool to achieve policy objectives, including on politically challenging local issues. Landmark legal cases challenged national legislation and may be a catalyst for societal and legal change. | - Strength of national policy - Decision making process | - Increase regulations/ powers - Clarity in national policy - Impact on implementation - Innovative practices |
| 58 | .2.2. Influential relationships  How the ‘rules’ for urban development were followed could be influenced by formal and informal relationships, with values and motivations affecting how environments associated with health were considered.  In England's discretionary planning system negotiations occur between developers and local authorities. Interviewees said development costs were opaque and negotiations involved “brinkmanship” (SC-355, local government, urban development). Developers were said to “play local authorities off against each other” (QT-613, local government, transport) by threatening to develop in another local authority. An alternative to “the traditional slightly combative situation with developer/council” (MK-132, private sector, property developer/investor) were partnerships between developers and local government which could be beneficial where values aligned. However, one local government interviewee said lack of capacity could restrict such major procurement processes.  Developers appeared to have close connections with many stakeholders across the system. Some interviewees talked about the importance of working with people they know and trust. However, there were accusations of an ‘old boys network’ in some private sector organisations which may limit diversity of views, including lack of representation by young people, women and minority groups. | Partnership, collaboration, relationships, engagement | - Uncertainty - Role of private sector - Professionalism and impact on profession |
| 58 | 3.3.1. Defining and evidencing health  Clarifying what was meant by ‘healthy’ development, using evidence, could increase objectivity for decision-making to prioritise health. Quantitative measures, including framing health in commercial terms, may incentivise some stakeholders.  Health and wellbeing outcomes can be associated with a broad array of environmental factors. Defining ‘healthy’ development appeared a necessary, albeit often missing, step to clarifying how to improve health outcomes. The interview questions did not define ‘healthy’ and some interviewees assumed narrow definitions (i.e. clinical, rather than prevention), whereas others described health as “too broad … because it embraces so many different elements” (GW-402, Local government, housing). Proxies for health, such as environmental conditions, active travel and air quality were described by some interviewees. | Evidence base | - Relevance of evidence base - Type of evidence - Use of evidence base - Health considerations |
| 58 | The lack of clarity about ‘healthy’ development appeared to make it difficult to consider trade-offs e.g. between environmental and health outcomes. The lack of clear epidemiological evidence demonstrating associations between the built environment and health outcomes appeared to limit ability to object to planning proposals. This could result in accusations that comments about health impacts on planning applications were “off-hand, unsubstantiated” (HM-336, local government, urban development). It was thought that health could not be used to object to planning applications because it would not stand up to examination by the government's Planning Inspectorate. Greater objectivity appeared necessary. | Evidence base | - Type of evidence base - Relevance of evidence base - Planning Inspectorate |
| 58 | .3.2. Obtaining resources to focus on health  To enable healthier places interviewees suggested funding priorities needed to shift, particularly by national government since local government resources were limited, bidding for funding could be inefficient, and developers likely unwilling to pay additional costs. Demand may affect the market's willingness to fund these costs.  Some interviewees thought national government should provide additional funding to local authorities, for example via subsidies for affordable housing, especially in poorer areas with high housing needs. However, long-term population benefits, compared to short term costs, did not appear to be incentivised in the current short-term political system. | Resourcing of local planning authorities | Funding |
| 55 | The arrows in Fig. 2 highlight our interpretation of key control mechanisms across stakeholders: policies/legislation (‘rules’), and funding. Political control is also shown to demonstrate relationships, and likely differing priorities, between political and non-political actors in local and national government. The figure highlights that public health and community groups are not clearly connected to these important control mechanisms, suggesting that they have low levels of influence in the system. | Impact of politics and politisation in planning | Governance |
| 59 | Finding 2: Health is rarely featured in policy acting on the urban environment Conceptualizations of health common to public health such as preventing disease, prolonging life and promoting good physical and mental health rarely featured to any substantial extent in either housing or transport documents. We identified scant examples of an explicit emphasis on influencing health outcomes, such as morbidity, mortality or specific health conditions, and a lack of any clear and measurable health targets from implementing urban policies. Similarly, overarching concepts such as public health, health improvement, and health protection were very rarely discussed compared with the dominant and secondary factors. When mentioned, more commonly in transport documents, health was typically referred to briefly and in broad terms, such as stating the desire to improve health or quality of life without providing detail such as targets or specific conditions. For example, in the 2017 “Transport Investment Strategy” the case is made for a transport investment strategy and refers to the importance of safeguarding health with no elaboration on what this means or how it should be delivered.  Current UK housing and transport policies rarely prioritize health outcomes and fail to provide the context through which healthy urban development can be achieved. Future policy acting on urban development will need to consider how to protect against and mitigate the impacts of future infectious disease outbreaks (Megahed & Ghoneim, 2020) and, more widely, the increased attention on the association between health, inequalities and urban development presents an opportunity to go further. | - Strength of national policy - Monitoring and evaluation | - Health considerations - Clarity in national policy |
| 59 | However, urban policy conditions in the UK may not be conducive to taking advantage of this opportunity: government does not prioritize health outcomes, and there is insufficient cross-departmental collaboration on health despite the urban environment’s critical role as a health determinant | - Strength of national policy - Complexity of planning regulatory framework | - Health considerations - Governance |
| 60 | Establishing this cross-government strategy requires buy-in and commitment from the top of government and across key departments and leaders. Evidencing the interactive effects between improving health outcomes and dominant urban policy agendas can help to incentivize this shared accountability | - Leadership - Partnership, collaboration, relationships, engagement | - Governance - Access to evidence base |
| 59 | Recommendation 1: Health needs to be a direct outcome of policy The hierarchy of justifications for the housing and transport policy spaces leave little doubt that national government policy creating urban environments does not prioritize health. Notions of health common to public health as preventing disease, prolonging life and promoting good physical and mental health rarely featured in the documents we analysed. Where it is included, it is an assumed indirect outcome from achieving other dominant policy objectives. This implicit assumption that health benefits will be created through the achievement of other policy objectives, such as reducing air pollution, is a crucial insight. There is little evidence to suggest that policies assuming health benefits as implied outcomes of other objectives will result in anything more than ancillary or ad hoc improvements in health. For example, policy actions to improve environmental sustainability appear to have mixed or little impact on health measures (Swann et al., 2019). | - Visioning and outcomes-focused | - Health considerations - Deliver wider value and benefits |
| 59 | Integrating health and health actors into urban decision-making and including health outcomes to measure policy impact, and not simply numbers of houses being built, is vital to ensure urban development foregrounds good health for all (Wernham & Teutsch, 2015; World Health Organization & UN-Habitat, 2016). Beyond broad rhetoric about the importance of healthy urban environments, policies must include clear and measurable health targets if they are to be effective (Giles-Corti et al., 2022; Lowe et al., 2022). Where health objectives and justifications are not included in policy shaping the urban environment, non-health actors may overlook the health implications of their decisions and opportunities may be missed (Corburn et al., 2014) | - Monitoring and evaluation - Decision-making process | - Impact on implementation |
| 59 | Recommendation 2: Health sensitive policy requires cross-sector, collaborative action. Establishing health front and centre in urban policies and delivering on such objectives will require greater collaborative action across government. Currently, the UK policy environment is insufficiently managing the cross-sector issue of health and even where health is included in urban policies, there is insufficient collaboration between health experts and other policymakers. The following extract from the government’s response to a recent report by the Building Better, Building Back Building Beautiful Commission (2020) that called for the creation of beautiful urban areas illustrates this. On the one hand, it promotes the importance of supporting good health and well-being, yet it omits health stakeholders from the list of important partners and leaders in its delivery:  To fulfil this role, the Secretary of State works collaboratively with many other Cabinet Ministers to ensure successful placemaking, including with the Secretary of State for Transport, the Secretary of State for Environment, Food and Rural Affairs and the Secretary of State for Business, Energy and Industrial Strategy, and with relevant Ministers of State, in particular, the Minister for Housing and Minister of State for Regional Growth and Local Government. (MHCLG, 2021c, p. 33) The WHO emphasizes the need for cooperation between public health, planning and environmental sectors to support better health (Prüss-Üstün et al., 2016), and a lack of partnership working in the planning system has long been recognized as a barrier to integrating health into decision-making (Carmichael et al., 2012; Wernham & Teutsch, 2015). | Partnership, collaboration, relationships, engagement | - Lack of engagement - Externalised expertise - Governance |
| 60 | Recommendation 3: Shared accountability in national policy is needed to embed health in policymaking Increasing shared accountability and collaborative working across areas of national policy on health appears key therefore for supporting the integration of health outcomes in urban policies. This requires a coordinated cross-government approach to establish health prevention in areas of policy outside of the health sector (Iacobucci, 2022; O’Dowd, 2023). | Partnership, collaboration, relationships, engagement | - Governance - Lack of engagement |
| 60 | Embedding a similar “whole of government” approach whereby actors work across departmental boundaries to understand policy co-benefits and other agendas (Ortenzi et al., 2022) for health prevention is critical if health outcomes are to be integrated into urban development policy. This will require leadership at national level underpinned by long-term thinking and commitment to promote the health agenda (Guglielmin et al., 2018; Stahl, 2018) with ownership for the strategy from the Prime Minister and Treasury who have the power to drive this agenda (Dixon & Everest, 2021; Merrifield & Nightingale, 2021). This would not only raise the health agenda in national policy documents but can strengthen the integration of health into other policy areas at local level (Guglielmin et al., 2018). | - Leadership - Strength of national policy | - Governance - Clarity in national policy |
| 60 | Recommendation 4: Emphasizing the interactive effects between health and key urban policy agendas can incentivize stakeholders across government silos. To reach across silos and incentivize a shared pro-health agenda, messages can be reframed to demonstrate how health can help to deliver dominant agendas. Our review demonstrates how health outcomes currently fall underneath other departmental agendas in a hierarchy of priorities for urban policy actors. “Win-win” approaches that emphasize the interactive effects between health, economic and environmental benefits can create the case for investing in health without reducing attention on other agendas (Freiler et al., 2013; Molnar et al., 2016). Reframing how health is important for all policies, rather than in all policies, will help to emphasize mutual benefits across policy areas from improving population health (Greer et al., 2022). For example, supporting a healthier workforce will improve productivity and reduce absence from work, which can help drive economic growth and innovation (British Medical Association [BMA], 2022).Healthier adults are more likely to choose alternative transportation than using their car (Bopp et al., 2013; Naumann et al., 2009), which could help to deliver on agendas on air pollution, transport congestion and decarbonization. These examples illustrate how health is not only a byproduct of delivering other important agendas, but is a critical factor in their success that may incentivize stakeholders who are not primarily concerned with health objectives. | Visioning and outcomes-focused | - Governance - Deliver wider value and benefits |
| 61 | 2019 | Resourcing of local planning authorities  Capability, skills and knowledge |  |
| 61 | 7 Local authorities have struggled to produce up-to-date local plans. As of December 2018, only 44.1% of local authorities had an up-to-date local plan (a plan less than five years old) that sets out their strategies for meeting the need for new homes. All local authorities should have a local plan and identify a five-year supply of land for new homes. Producing local plans can be technically complex, resource-intensive and time-consuming. As of November 2018, the Department – through the Secretary of State – has only challenged 15 local authorities that do not have an up-to-date plan. If a local authority does not show it has a five-year supply of land for housing, it gives developers greater freedoms to build where they want, and a local authority has less control over the location of development. This limited local authority control risks ill-suited developments (paragraphs 1.10 to 1.14 and Figure 6. | Local plan content and plan-making process | - Up to date local plan - Impact on implementation |
| 61 | 8 The Department’s standard method for assessing the need for new homes has weaknesses and as a result will be revised. In 2017, the Department developed a standard method for local authorities to assess the number of new homes needed in their area. Previously, local authorities used a variety of methods to calculate this. The Department’s standard method is based on projections of the growth in the numbers of households, adjusted for the affordability of housing in local areas. It is unclear whether the methodology is consistent with the overall ambition for 300,000 new homes per year by the mid-2020s. The Department intends to revise the methodology to be consistent with ensuring that 300,000 homes are built per year by the mid-2020s (paragraphs 1.15 to 1.19).  9 Local authorities in four out of nine regions have seen an increase in the number of new homes needed in their areas. The standard method has changed the need for new housing when compared with the need assessed previously by local authorities | Strength of national policy | - |
| 62 | 12 According to the Department’s performance measures, local authorities are increasingly processing planning applications within target timescales. The percentage of major residential applications that local authorities determined within the target of 13 weeks or agreed extended period, increased from 47% in 2012-13 to 87% in 2017-18. During the same period, local authorities dealt with more major residential applications. The number increased from 5,244 in 2012-13 to 7,997 in 2017-18. In 2017-18, local authorities approved 81% of major residential planning applications (paragraphs 2.3 and 2.4, Figure 9).  13 However, some of the reported improvement in local authorities’ performance might reflect a greater use of agreed extensions to timescales rather than increased efficiency. | Decision-making process | - Better use of local levers - Impact on implementation |
| 62 | 14 The Planning Inspectorate is slow at determining appeals and acknowledges its performance is unacceptable | Decision-making process | Planning Inspectorate |
| 62 | Constraints on the planning system  21 Total spending by local authorities on planning functions fell 14.6% in real-terms between 2010-11 and 2017-18; local authorities increased their income to avoid further reductions. Between 2010-11 and 2017-18, there was a 37.9% real-terms fall in net current expenditure (expenditure funded by an authority’s own resources) on planning functions. However, increased income generated from sales, fees and charges or transfers from other public authorities meant that total spending on planning reduced in real-terms by 14.6%; from £1.125 billion in 2010-11 to £961 million in 2017-18 (paragraphs 3.2 to 3.5 and Figure 14). | Resourcing of local planning authorities | Planning fees |
| 63 | 22 The Department does not understand the extent of skills shortages in planning. In 2017, the Department pledged to help ensure that the planning system has enough skilled professionals. While local authorities complain of a shortage of planners, data on staff numbers are patchy and the Department does not collate comprehensive data on the extent of this shortage. Research in 2017 indicated that the number of local authority planning staff fell 15% overall between 2006 and 2016. As of the end of 2018, the Department had made some efforts to deal with the shortages of planners by helping to fund a bursary scheme and supporting a bid by the Royal Town Planning Institute for a degree-level planning apprenticeship (paragraphs 3.6 to 3.11).  23 The Planning Inspectorate has failed to recruit the right number of inspectors. Between 2010 and 2018, the Planning Inspectorate experienced a 13% fall in staff numbers, amounting to almost 100 full-time equivalent staff. The Planning Inspectorate does not have detailed workforce plans to show how it will use existing and any newly recruited staff effectively, and deal with future workload pressures | - Capability, skills and knowledge - Resourcing of local planning authorities | - Access to evidence base - Professional qualification - Planning Inspectorate - Externalised expertise - Professionalism and impact on profession |
| 64 | 2023 | Resourcing of local planning authorities  Capability, skills and knowledge |  |
| 64 | The first question asks about barriers that may have limited a council’s use of externally produced evidence in formulating and implementing health-related components of their Climate Action Plans. The most common challenge when using external evidence was finding the time and resources to assess its quality and relevance (n = 14). | - Resourcing of local planning authorities - Evidence base | Relevance of evidence base |
| 64 | lacked the expertise to do this (n = 4) | Capability, skills and knowledge |  |
| 64 | struggled to find evidence that matched their local context (n = 3) | Evidence base | Relevance of evidence base |
| 64 | Other barriers included difficulty accessing the evidence (n = 2 | Evidence base | Access to evidence base |
| 64 | lack of support from senior leaders (n = 1) and lack of local political backing (n = 1). | Leadership | Political commitment |
| 64 | The second question asks about barriers to working with local communities when developing health-related components of a Climate Action Plan. The most common barrier reported was having enough capacity or resources to reach out to them (n = 12). | Resourcing of local planning authorities | - Lack of engagement - Role of local communities |
| 64 | Other barriers included poor relations or trust between communities and councils (n = 4), lack of existing links with communities (n = 1), reluctance of colleagues to involve communities (n = 1) and preference for working with other public sector organisations (n = 1). | Partnership, collaboration, relationships, engagement | - Lack of engagement - Role of local communities |
| 64 | The biggest barrier was not having enough staff and resources to carry out the plan (n = 9). | Resourcing of local planning authorities | Impact on implementation |
| 64 | Other barriers included organizational culture and values or awareness of colleagues including senior leadership (n = 3), difficulties engaging with colleagues from the health- care system (n = 3), | Partnership, collaboration, relationships, engagement | - Governance - Lack of engagement |
| 64 | too little being known about the health impacts of climate change (n = 3) | Capability, skills and knowledge |  |
| 64 | Others said they faced political barriers at a local (n = 2) or national level (n = 1) | Impact of politics and politisation in planning | - Role of elected members - Political commitment |
| 64 | In Fig. 2, the highest ranked barrier to implementation of health/health inequality-related components of climate action plans was insufficient staff and resources | Resourcing of local planning authorities | - |
| 64 | Of the other barriers to implementation, we see that the national political and policy context was the second highest ranked barrier to action. | Strength of national policy | - |
| 64 | The third highest ranked barrier concerned problems internal to the council such as organisational culture and values/awareness of colleagues (including senior leadership). Internal problems are also evident in the fourth ranked option of ’council colleagues working in silos, lack of joining up and collaboration’ | - Partnership, collaboration, relationships, engagement - Stakeholders and statutory consultees | - Governance - Lack of engagement - Lack of alignment |
| 64 | In Fig. 3, first-ranked among facilitators of implementation was ’the need to save money on energy expenditure’, serving as a financial incentive for more urgent action. | Impact of economic focus, viability and costs | Cost and delay |
| 64 | Closely following behind in second place was ’effective collaboration and joined up working’ | Partnership, collaboration, relationships, engagement | - |
| 64 | receiving support in its ranking of fourth place as a facilitator was "the national political and policy context is conducive", suggesting some participants perceive there to be a degree of support from national government and policymakers | Strength of national policy | - |
| 65 | In Fig. 4, of the barriers to use of evidence, ’Competing demands/lack of resources and time required to assess the evidence’ was the highest ranked choice | Resourcing of local planning authorities | - |
| 65 | Many participants believed their local authority lacked the internal expertise to assess the evidence (rank 2), | Capability, skills and knowledge | - |
| 65 | which was com- pounded by the inaccessibility of the evidence itself (rank  3). The sense of confusion and uncertainty over the evidence was added to by the perception that there is a lack of evidence relevant to their locality (rank 4). | Evidence base | Access to evidence |
| 65 | The option ranked in first position by participants was ’greater collaboration between climate and public health teams’ | Partnership, collaboration, relationships, engagement | - |
| 65 | Receiving almost the same overall score, the options in second and third position were ’commitments to adequately staffing and resourcing research capacity’ and ’collaboration with academic partners’ | Resourcing of local planning authorities | Externalised expertise |
| 65 | Figure 6 shows the barriers to working with local com- munities and stakeholders. The first ranked barrier by participants is ’Lack of capacity/resources to engage with those outside of the council’ | Resourcing of local planning authorities | - |
| 65 | The second highest ranked barrier was ’challenges reaching specific communities or demographics’ highlighting the challenges accessing seldom heard communities. The third-highest barrier of ’lack of interest/knowledge within the local community’ may mean many communities are unaware of the value of climate action plans. The next highest ranked barrier ’community-council relations – lack of trust in the council’ may point to unfavourable relations with local communities | Partnership, collaboration, relationships, engagement | Role of local communities |
| 66 | Finances, resources, personnel, and prioritisation of the issues  Financial and resource constraints were highly ranked as a barrier across a number of domains, including implementation, use of evidence and engaging with communities. Conversely, some respondents ranked finance and resources as a key facilitator for the implementation of health/health inequality-related components of local authorities’ climate action plans, use of evidence, and working with local communities. | Resourcing of local planning authorities | - Funding - Use of evidence - Role of local communities |
| 66 | Cultural and organisational readiness for the challenge Barriers related to internal culture and organisation were found to be significant in hindering the implementation of health/health inequality-related components of local authorities’ climate action plans. In contrast, for some other respondents, culture and internal practices were more conducive, with internal culture and organisation identified as key facilitators of implementing health/ health inequality-related components of local authorities’ climate action plans. Regarding facilitators of working with local communities and stakeholders, the second ranked choice was ’working closely with other services that are better connected to local communities’ | Partnership, collaboration, relationships, engagement | - Governance - Externalised expertise |
| 66 | National political and policy context  The national political and policy context is ranked in second position among all barriers to implementation of health/health inequality-related components of climate action plans and thus is perceived as a major hindrance to action by respondents, suggesting some level of unease with wider issues of political will and policy coherence. Ranked fourth of six facilitators of implementation is "the national political and policy context is conducive", suggesting some participants perceived there to be a degree of support from national government and policymakers | - Strength of national policy - Impact of politics and politisation in planning | Political commitment |
| 66 | Collaboration with external partners  To improve access to local community groups, the fourth-ranked facilitator is the ’creation of multi-agency organisations’. This strategy may prove effective in over- coming persistent barriers to co-production, enabling local authorities to work more closely and efficiently with external partners in the pursuit of common goals. In third position among facilitators of use of evidence was ’collaboration with academic partners’. The perception that closer collaboration with academic partners may be beneficial indicates those local authorities with greater access to academic partners, such as those located close to universities or with pre-existing links, may be better positioned to draw on evidence in their climate action plans. | Partnership, collaboration, relationships, engagement | - Governance - Lack of alignment - Externalised expertise |
| 64 | Resource constraints, institutional fragmentation, lack of political support, and competing priorities pose significant challenges for local authorities seeking to implement climate action plans that prioritise health equity | - Resourcing of local planning authorities - Impact of politics and politisation in planning | - Governance - Political commitment - Lack of alignment |
| 64 | To overcome these barriers, local authorities must nurture a culture of innovation, collaboration, and purpose, while also addressing the urgent need to protect vulnerable communities from the health impacts of climate change. This means not just working with external partners, but also collaborating and co-producing with communities to achieve health equity and mitigate the debilitating effect of climate change on public health | - Knowledge transfer, good practices - Visioning and outcomes-focused | - Deliver wider value and benefits - Innovative practices - Role of local communities |
| 64 | Lastly, the overriding and unavoidable conclusion is that local authorities cannot solve these problems on their own - policymakers must provide the resources and support that local authorities need to make a real difference | Resourcing of local planning authorities |  |
| 67 | 2020 | Resourcing of local planning authorities |  |
| 67 | Responsibilities for infrastructure planning and for housing provision are fragmented across various stakeholders, including local authorities, central government bodies, private developers, arms-length bodies, and utilities companies (Graham and Marvin, 2001; National Audit Office, 2019). Although these stakeholders work together to deliver and integrate infrastructure and housing, they may have different motivations and seek different outcomes | Complexity of planning regulatory framework | - Governance - Lack of alignment |
| 67 | Coordination between key stakeholders is therefore essential in order to build successful places | Partnership, collaboration, relationships, engagement | Governance |
| 67 | Interviews with stakeholders confirmed that spatial planning is a key mechanism for effective collaboration between different levels of government. It provides a strategic vision for development, connecting local planning practices with national needs expressed through planning policy (Scottish Government, 2019a, 2019b, Hawkes, 2019). The benefits afforded by spatial planning, a joined-up approach which connects different spatial scales, go beyond delivering housing with infrastructure and could help solve other problems. For instance, it may help to identify ‘left-behind’ areas, enabling replacement of outdated infrastructure or promoting growth within those places (Williams, 2014, RTPI, 2019). Therefore, spatial planning is an effective vehicle for linking infrastructure planning and housing provision and has wide-ranging benefits. | Visioning and outcomes-focused | Deliver wider value and benefits |
| 67 | This research identified the need for a ‘larger than local’ approach to development, but also highlighted the fact that the possibilities for such an approach to strategic planning are somewhat restricted because of the gap left by the removal of the RSSs. The lack of regional guidance causes difficulties in coordinating infrastructure and housing planning effectively across different spatial scales, from national to local. | Impact of strategic planning practices | - |
| 67 | The integration of infrastructure development and investment is not only impacted by the lack of clarity between different aspects of government in their interaction with the private sector (National Audit Office, 2019), but also by the lack of centralised sources of funding for local authorities. Local authorities are more reliant upon developer contributions and source funding from multiple streams in order to fund infrastructure development, meaning that funding is piecemeal (RTPI, 2019). Consequently, the funding for infrastructure is focused more upon the short-term and the immediate returns for the funders. Therefore, in order to provide infrastructure alongside housing in a timely manner, there is a need for a centralised source of funding. | Resourcing of local planning authorities | - Funding - Lack of alignment |
| 67 | Collaboration between the public and private sector can be more efficiently coordinated through the wider use of institutional mechanisms like development corporations, or by the public sector taking a more proactive approach through strategic land assembly. Development corporations enable different public and private sector actors to come together with different resources and work with one another in establishing a common platform for development, e.g. public sector with land and the private sector with developmental expertise and monetary resources. Furthermore, strategic land assemblies are useful since they offer a mechanism to consolidate land under single ownership, enabling private sector developers to collaborate with the public sector and landowners to encourage development. These mechanisms offer a way for the public and private sector to effectively coordinate their activities | Partnership, collaboration, relationships, engagement | - Better use of local levers - Role of private sector |
| 68 | This research identified a range of stakeholders involved in the design, delivery, and maintenance of ‘good quality’ places, including local authorities, statutory bodies, private sector developers, and local communities (Dempsey and Smith, 2014; Beza and Hernandez-Garcia, 2018). The place-making process can be complex, challenging and time consuming because there are multiple stakeholders whose views and needs must be taken into account. Whilst consultation and adherence to Local Plans are designed to enable developers to meet the demands of existing stakeholders, the voice of future occupants of new developments is often missing from the consultation process. The prior experience of developers may mitigate this to a certain extent where they are able to account for which amenities and housing types are needed by people of differing ages, backgrounds and life stages. The design and delivery of ‘good quality’ places is complex, as it has to account for the different needs and wants of multiple existing and future stakeholders. Therefore, there is a real need to dedicate time and resources to facilitate open and inclusive stakeholder engagement. | - Stakeholders and statutory consultees - Partnership, collaboration, relationships, engagement | - Role of private sector - Role of local communities - Externalised expertise - Lack of engagement |
| 68 | Place-keeping is a process which involves stakeholders ranging from asset management companies, private developers, local authorities, local communities and user groups to form long-term management strategies and implement them (Dempsey and Smith, 2014). The influence that relevant stakeholders have in shaping their development thus extends beyond the initial construction of a place into the future | Stakeholders and statutory consultees | - |
| 67 | A layer of strategic planning beyond the local authority level, e.g. a tier of regional planning, is needed to ensure that infrastructure planning and housing provision are in step with one another. | Impact of strategic planning practices | Governance (Strategic planning) |
| 67 | Given the important role of planning in coordinating housing and infrastructure, there needs to be adequate resourcing for local planning departments and investment in the skills of planning officers | Resourcing of local planning authorities | Professional qualification |
| 67 | Effective public and private sector coordination for housing and infrastructure delivery should be facilitated by the wider use of institutional mechanisms (such as development corporations) and a more proactive public sector approach (including the use of strategic land assemblies). | Partnership, collaboration, relationships, engagement | Better use of local levers |
| 67 | There is a need for good practice guidance on consultation to support collaborative approaches to development consultation based on openness and transparency. Developers must commit adequate time and resources to facilitate this. Local residents may need support to engage effectively, particularly in disadvantaged communities | Partnership, collaboration, relationships, engagement | - Role of local communities - Lack of engagement |
| 67 | Industry guidance on good practice that balances the economic, social and environmental aspects of sustainable place-making needs to be developed. There would be value in developing a robust way to assess sustainability in the broadest sense, as well as identifying and disseminating good practice examples. | - Knowledge transfer, good practices - Use of impact assessment | Externalised expertise |
| 67 | To meet targets for carbon emissions that are economically viable and sustainable, developers need to collaborate with government and other actors not only when designing new places, but also in retrofitting existing developments. Achieving net-zero carbon needs sector commitment, technical innovation and resourcing | - Impact of economic focus, viability and costs | - Governance - Role of private sector - Innovative practices |
| 69 | 2017 | Capability, skills and knowledge |  |
| 69 | Policymakers were asked which type of information or evidence they most frequently used and what were the most useful types of information or evidence (figure 1). In both categories, local data were the most frequently cited type of evidence (95% and 80% of respondents, respectively). | Evidence base | - Type of evidence base - Use of evidence |
| 69 | The second most cited source of evidence was the joint strategic needs assessments, selected by 89% as ‘frequently used’ | Evidence base | Type of evidence base |
| 69 | Research-derived evidence was considered useful and used by most participants, especially qualitative research studies and survey data. Systematic review and trial data were also used (47% and 23%, respectively) but were much less popular. | Evidence base | - Type of evidence base - Use of evidence |
| 69 | Survey data and PH surveillance data were both more ‘used’ than that they were considered to be ‘useful’. Several respondents (n = 7) however, commented that these types of evidence were useful for describing problems but did not provide solutions useful for decision actors. | Evidence base | - Type of evidence base - Use of evidence - Relevance of evidence base |
| 69 | Respondents were also asked if there were other types of regularly used evidence not mentioned in the closed categories. Answers provided included professional advice, political and local ‘soft’ in- formation from ‘the field’—meaning knowledge of local personalities, local systems and organizations. | Evidence base | - Type of evidence base |
| 69 | Policymakers described a lack of capacity for analysis, and wanting assistance with analysing local data in more sophisticated ways; for instance to model care pathways to understand what works for different groups; to understand the interplay of local factors and roles of different actors at points in the care pathway; to understand cross- sector and cross-governmental department interventions and how to use more active and rigorous audit loops so that ineffective interventions could be halted. | Capability, skills and knowledge | - Use of evidence base |
| 69 | The most frequently mentioned sources were governmental websites (84%), followed by National Institute for Health and Care (NICE) guidelines (70%). ‘Experts’ and ‘other people’ were both chosen by over 70% of respondents, with ‘Experts’ ranked 4th and other people was chosen, and how it was ranked by participants. | Evidence base | - Access to evidence base - Externalised expertise |
| 71 | 2021 | Capability, skills and knowledge |  |
| 71 | In line with the findings from the GRIP project (5), there was general agreement across participants in the four locations that health evidence could be developed and used more effectively in planning policy | Evidence base | Use of evidence base |
| 71 | A second set of comments was made on the types of evidence people were referring to when talking about health-related evidence | Evidence base | Type of evidence base |
| 71 | A third set of comments concerned the characteristics and attributes of the evidence. The GRIP project (5) highlighted the differences in the interpretation and use of evidence across public health and built environment disciplines, and the need to build on the evidence provided in Spatial Planning for Health (15) to ensure that health evidence is applicable to planning policy and development management. In this research, workshop participants across the four locations expressed the two primary needs and aspirations in this respect. Other aspects concerning the language and interpretation of evidence were also considered and these are addressed in the section on the local resources. | Evidence base | - Type of evidence - Relevance of evidence |
| 72 | Delegates had varying degrees of knowledge and awareness of the available existing health evidence resources, with some admitting not to be aware of such resources. A number of delegates raised concerns over the sheer amount of public health evidence available, the need to distil it down into an essential evidence base, and to make it available, usable, easily accessible and interpretable, especially when those wishing to use it are non-experts and may not possess the skills to interpret and use the evidence, e.g. those involved in the production of an NPD (Gloucestershire). | - Evidence base - Capability, skills and knowledge | - Access to evidence base (Awareness) - Use of evidence |
| 72 | Challenges related to public health and planning being separate professions with different vocabulary, ways of working, policy development processes and gaps in understanding each other’s responsibilities and areas of influence.  As was a key finding in GRIP (5), participants across the four locations recognised that planning and health are largely separate policy domains. They provided examples of the challenges this brings highlighting three areas of disconnect: language and communication, organisational and structural, and documentation. In turn, these challenges can affect the provision of appropriate evidence and the effective use of the evidence provided. It is necessary to recognise that these locations were specifically chosen because they have not received support from PHE or TCPA in the past, so are likely to feel these barriers more acutely than areas that have already developed or begun to develop policies on planning for health | Complexity of planning regulatory framework | - Professionalism and impact on profession - Governance - (Less) focus on process |
| 72 | The lack of shared language between the two disciplines was identified as a challenge: “There is still a big disconnect between the worlds – public health commissioners are not equipped with the knowledge or language”. This is despite public health teams being transferred to local authorities following the Health and Social Care Act (HSCA), 2012, and health being a recurrent theme in the National Planning Policy Framework (NPPF) (10). | - Capability, skills and knowledge - Strength of national policy | - Impact of public health reforms - Clarity in national policy |
| 73 | The lack of shared education and knowledge between public health and planning professions was identified as a challenge. For example, it was also mentioned that the lack of health content in planning degrees “means that data/evidence needs to be presented in a lay fashion for planners, e.g. based on traffic lights” (Worcestershire). Likewise, some public health professionals lack planning knowledge | Training, learning and development | Professional qualification |
| 73 | Other additional challenges concerned the different, and often disjointed, levels of governance within local authorities, the tendency to work in silos, and different working practices in the public health and planning domains | Complexity of planning regulatory framework | - Governance - Lack of alignment |
| 73 | Challenges related to maximising opportunities in the planning process to integrate health and planning.  In different ways, each workshop highlighted areas where the opportunities for planning to deliver better health outcomes were being missed or where the complexity of the planning system was not always well understood by other stakeholders. For example, it was suggested in the Hull workshop that there are opportunities for the Planning Inspectorate to better reflect and implement current thinking or emphasis around health in decision making, a factor also reported in GRIP | Complexity of planning regulatory framework | - Planning Inspectorate - Health Considerations |
| 73 | Challenges related to the stakeholders/communities involved in producing planning policies/documents.  Translating the available health evidence into planning was seen as dependent upon the stakeholders, practitioners and communities involved in the development of planning policies | - Evidence base - Capability, skills and knowledge | - Use of evidence base - Professional qualification |
| 74 | Regulations and controls over other areas of local government, such as licensing and procurement, were perceived as important enablers to improving health outcomes (e.g. reducing obesity). Examples mentioned by participants included conditions on opening hours for hot food takeaways, controls over concentrations of betting shops, payday lenders and casinos: “Licensing is critical, it needs to work alongside and in addition to planning to ensure robustness in approach” (Hull). Participants suggested that regulation should require all new developments to be planned with space to grow food, but the challenges of rural locations were highlighted in this context: “Many rural communities are becoming food deserts despite being very close to where fresh food is actually produced” (YNYER). There is, in the absence of national mandatory standards, perhaps an opportunity for local authorities to set expectations on the use of accreditation systems in new developments in their areas and ensure that these work synergistically with regulations from other areas as planning can only ever solve part of the problem. | - Strength of national policy - Use of (local) planning powers | Better use of local levers |
| 74 | Evaluating the outcomes of planning policy interventions wherever possible (e.g. active travel plans, community gardens, public gyms etc.) and the robustness of the health evidence itself, as well as learning from past experience, were seen as key enablers to translating evidence into planning. Participants in Hull suggested that an independent organisation could be in charge of the monitoring and evaluation task. Linking local authorities’ Authority Monitoring Report (AMR) with health data was also seen as a potentially useful tool to monitor the outcome of interventions. AMRs are an important part of local authority reporting on planning. Although, there are inevitably challenges around attributing causality. | Monitoring and evaluation | - Use of evidence base - Better use of local levers |
| 74 | Helpful ways of working  From a policy process perspective, effective working practices emerged among the key enabling factors. The inclusion of “subject matter experts” (Worcestershire) and “partnership working” were considered contributors to successful policy implementation and achievement of positive health outcomes (Hull). In Gloucestershire, it was considered helpful to involve local organisations such as residents’ associations “to make sure that the views of left-out communities are included in the development of Neighbourhood Plans”. However, it should be noted that those involved in residents’ groups are often also not typical of the areas they represent (19). Other stakeholders to include in the policy process were mentioned, for example parish councils, developers and “community builders”. Examples of good practice noted at the workshops included running joint CPD courses with planning | Capability, skills and knowledge | - Externalised expertise - Role of local communities - Role of private sector - Professional qualification (knowledge) |
| 71 | All those involved in the planning and development process must understand the importance of planning in tackling poor health and health inequalities, including central and local government planning policymakers, and those working in development management, private developers and their consultants. There is variation in understanding and practices in many parts of the process, which is hampering progress in some areas. | Visioning and outcomes-focused | - |
| 71 | Disparities in practice amongst planning inspectors concerning the potential, scope, and ability to integrate enhanced health and wellbeing derived requirements into the planning policy context, particularly with regards the Development Plan creation, examination, and adoption process, is causing uncertainty and a lack of confidence in authorities about how to ensure health is integrated into their policies | Decision-making | - Planning Inspectorate - Health considerations - Uncertainty - Confidence |
| 71 | Elected members should be supported to better understand the relationship between planning and the built environment, and health and wellbeing outcomes;  Stakeholders in different areas of local authorities should be supported to better understand the contribution they can make, for example those involved in highways planning | Stakeholders and statutory consultees | - Role of elected members - Professionalism and impact on profession |
| 71 | There is a role for public health teams in local authorities to enhance planning officers’ knowledge concerning health inequalities, and their relationship with the built environment. Although the roots of planning are in improving health, planners have not always approached planning policy and decision-making in the context of health inequalities in the manner now expected. There is a role for public health teams to strengthen planners’ understanding of health inequalities, for example, through joint CPD, particularly the priorities in their locations, and how planning policies and decision could help reduce these inequalities or, if they are not considered, make them worse | Training, learning and development | - Professional qualification (knowledge) - Health considerations - Deliver wider value and benefits |
| 71 | Planning policy teams could articulate the contribution planning can make to improving health and reducing health inequalities. There is a lack of awareness of the wider determinants of health amongst both professional and community groups, so policymakers need to set this out prominently in relevant policies. It may also be that the word ‘health’ is unhelpful, and that alternative terms, such as wellbeing, may be more readily understood, particularly for Neighbourhood Planning Groups or during consultation activities | Capability, skills and knowledge | - Health consideration - Clarity in national policy - Better use of local levers - Role of local communities |
| 71 | Planning policy teams could develop their understanding of how public health evidence can help them achieve their policy objectives. There are multiple opportunities to make better use of public health evidence in planning and public health teams can support planners to maximise these | Evidence base | - Use of evidence base |
| 72 | Health evidence can be presented to decision makers more effectively, for example, by presenting the consequences of doing nothing to enable healthy planning, the cost savings to other areas of local government, and providing positive examples from other authorities. Public health teams can work with planners to ensure that evidence they produce is more usable for planners, and help with interpreting the evidence. Evidence presented at ward or authority level may hide pockets of health inequalities; reducing the ability to target interventions to where they are most need. Similarly, comparisons with national data are not helpful as there are poor health outcomes at the national level, so using this as a benchmark provides an inaccurately positive picture. Critically, health evidence must be specific and precise to support and underpin planning decision making and the effective application of planning policy | Evidence base | - Access to evidence base - Relevance of evidence base - Type of evidence base - Impact on implementation |
| 72 | Planning policy teams could seek the views of a wide range of stakeholders when interpreting and using evidence. There is a need to recognise that different stakeholders interpret evidence in different ways, and this can bias the ways in which evidence is used. Often planning and public health professionals represent a subset of the population and there is a need to ensure that the voices of all the community are represented, particularly those from marginalised groups who may suffer the greatest health inequalities.  Planning policy and public health teams could draw from a broad range of evidence, including that generated by community groups. Linked to the above, there is a tendency to value quantitative evidence from national datasets more than locally generated data from grassroots organisations. Publications and research documents (e.g. Spatial Planning for Health) are very valuable, but it’s vital that they are supplemented with qualitative and locally generated evidence that presents the lived experiences of local people, particularly the least healthy and most marginalised | Evidence base  Stakeholders and statutory consultees | - Use of evidence base - Type of evidence base - Role of local communities |
| 72 | Public health and planning professionals can work together to develop a shared understanding of the role of planning in improving population health and reducing health inequalities. Differences in the use of evidence, language and practices of the different disciplines need to be recognised and overcome. This is particularly important in understanding how public health evidence can facilitate better planning outcomes, but also in recognising the limits of the planning system and what it can and can’t achieve. This will allow scarce resources to be targeted where they can make the greatest difference | - Evidence base - Resourcing of local planning authorities | - |
| 73 | Public health evidence can help planners use their powers more effectively. The use of local standards (e.g. Live Well Accreditation) or accreditation systems (e.g. Lifetime Homes, Building with Nature) was seen as key to delivering healthy places, and planners have the powers to require these into their local policies. Local public health evidence that sets out the health priorities for the area, and evidence of the relationship between built environment interventions and health outcomes, such as that provided in Spatial Planning for Health, can provide the necessary weight for such standards. | - Use of (local) planning powers - Decision-making process | - Use of evidence base - Better use of local levers |
| 73 | Public health teams should support planners in monitoring and evaluating planning policies. Evaluating the effectiveness of policies and interventions in the built environment is often neglected, but often existing evidence can support this. It is crucial that this is prioritised and robust monitoring and evaluation takes place to test what works and allow other locations to learn from front runners, and target resources effectively | Monitoring and evaluation | Innovative practices |
| 75 | 2018 | - Capability, skills and knowledge - Resourcing of local planning authorities |  |
| 75 | These differences in understandings of evidence can prevent the integration of health into urban and transport planning as options that appeal to individual decision makers acting in specific contexts are chosen over what the hard evidence stipulates. Effective evidence translation is difficult. Whilst research on the relationship between the built environment and health is growing, it is rarely picked up by policymakers or practitioners. Frequently, a ‘gap’ emerges between research on the one hand and those responsible for its implementation on the other. This results in suboptimal decision making in terms of health promotion as the evidence is neglected or misunderstood | - Evidence base - Decision-making | - Relevance of evidence base - Health consideration - Impact on implementation |
| 75 | Finally, research often fails to take into account the complex environments in which decision makers operate. Additional difficulties lie in the different timeframes and pressures of the policymaking process as opposed to the lengthy peer review process of the scientific research sector making further collaboration disjointed (Orton et al. 2011). | - Complexity of planning regulatory framework | - Lack of alignment - (Less) focus on process |
| 75 | 31.2.3 Governance and Politics  Issues around governance and the political context exacerbate the disconnect between the health evidence base and urban and transport planning decision making. Whilst evidence-based public policy in principal draws on robust evidence to determine appropriate courses of policy direction, in reality decisions are often made on political grounds (Davis 2014; Juntti et al. 2009; Phillips and Green 2015). As Davis comments ‘politics and power take the lead in determining what and how evidence is used’ (Davis 2014). Urban and transport planners in local government are accountable to a variety of stakeholder groups in a way that public health practitioners have traditionally not been (Phillips and Green 2015). As part of the decision- making process, urban and transport planners weigh up the interests of various stakeholder groups including politicians, residents, local businesses, internal departments and health services. Often these groups have different competing priorities and interests. | Impact of politics and politisation in planning | - Lack of alignment - External influence - Use of evidence base |
| 75 | The shortism of the political cycle means that an incoming administration often engenders a shift in policy focus in order to distinguish  itself from predecessors. Therefore, whilst one administration may seek to address the upstream determinants of health and encourage the integration of health into urban and transport planning, this policy may stagnate or be reversed with the next administration. Frequently, the extent to which a political leader seeks to address the upstream determinants comes down to personal preferences and understanding of health | Impact of politics and politisation in planning | - Political commitment - Role of elected members |
| 75 | The absence of health as a key consideration in land-use regulation presents a further impediment. Whilst guidance on how to build healthier urban environments is increasing, regulation is weak. The lack of regulation mandating the integration of health into urban and transport planning becomes a more acute issue amidst the political drive for more affordable housing and jobs. | Strength of national policy | Clarity in national policy |
| 76 | In addition, general lack of budget within departments removes incentives to embark on schemes outside of business-as-usual-type activities. For example, underfunded and under-resourced health departments are often under pressure to provide basic needs such as access to medical and sexual health services or care for the elderly. Within this context of limited resources, schemes that address health determinants within the built environment are often considered a secondary priority and therefore neglected. Institutional capacity exacerbates this problem. Cuts to personnel result in a fewer number of staff responsible for the same amount of workload. Therefore, solutions and programmes that imply additional work in the short term or that seem to detract attention from direct and current health concerns face internal opposition. | Resourcing of local planning authorities | - Funding - Governance |
| 76 | In an under-resourced environment, value for money and the business case of different choices become important considerations. Decision makers employ cost-benefit analysis in order to weigh up options. Cost-benefit analysis becomes the proxy upon which decisions are made. Quantifying health outcomes is difficult, and despite advances in metrics such as the Health Economic Assessment Tool, benefits to cost-type evaluations frequently do not sufficiently account for the health risks or benefits (Cavill et al. 2012; Fishman et al. 2015; Rojas-Rueda et al. 2016). Effectively accounting for health benefits of different options is complex leading to suboptimal decision making. Additionally, where the benefit to cost ratio is evident, frequently the positive outcome occurs at a time in the future beyond that stipulated by the funding. Therefore, issues relating to the nature of quantifying the health benefits as well as the timeframes of funding make it difficult for decision makers to choose health-optimising solutions over other policy options. | - Impact of economic focus, viability and costs - Use of impact assessment | - Lack of alignment - Health considerations |
| 76 | 31.3.1 Institutional Context  To start, changes can be made to the structure of the institutional environment in which urban and transport planning decisions are made. Intersectoral collaboration could be encouraged through formal and informal structures. Formal structures that encourage collaboration include joint work programmes across departments, joint funding bids, co-location of health professionals within planning departments and intersectoral advisory boards and task forces | Partnership, collaboration, relationships, engagement | - Governance - Funding - Lack of alignment - Dedicated posts and capacity |
| 76 | Funding a permanent member of staff with intersectoral responsibilities may encourage the inclusion of health into urban and transport planning (Thomas et al. 2009). Suffusing public health teams across departments can break down silos and assist in building understanding, trust and rapport, therefore, encouraging collaboration. Such institutional changes can encourage the integration of health into decision making. | Capability, skills and knowledge | - Dedicated posts and capacity - Health considerations |
| 76 | These positive changes can be compounded by increasing the availability of resources and capacity. Specifically allocating budget for a member of staff with responsibility for the integration of health is likely to be effective (Schwarte et al.2010). In the UK, the co-location of health professionals within Transport for London, the organisation responsible for the direction and delivery of London’s transport system, has driven health up the agenda | Capability, skills and knowledge | Dedicated posts and capacity |
| 76 | Funding for training and education to improve understanding of different professions is also needed. Specifically, training for transport and urban planners around the health impacts of their work and the wider determinants of health is likely to increase collaboration and understanding (World Health Organisation 2017). Such training should be included in the urban and transport planning curriculum for further generations. | Training, learning and development | Professional qualification (knowledge) |
| 76 | 31.3.2 Political Context  In terms of political context, greater engagement is needed with politics on the part of health (Hunter 2015; Kickbusch 2015; Mackenbach 2013). Public health training should equip professionals with the ability to understand and analyse political context, understand complexity of the decision-making environment and frame arguments effectively to influence policy | - Impact of politics and politisation in planning - Training, learning and development | - Lack of engagement - Health considerations |
| 76 | Stronger regulation to integrate health into land-use considerations may assist planners to push back against suboptimal development proposals in terms of health promotion. Whilst guidance on how planners can integrate health is increasing, regulation that mandates the integration of health into urban and transport planning is sparse | Strength of national policy | Increase regulations/ powers |
| 77 | 31.3.3 Evidence  Solutions to overcoming challenges of effective evidence translation in part have their roots in collaboration between policymakers, practitioners and researchers.  Collaboration between policymakers and researchers to co-identify effective and appropriate research questions that directly assist practitioners faced with challenges in the real world is important (Innaever et al. 2002; Oliver et al. 2014). Researchers could also work to better communicate their findings with policymakers and suggest interventions that are actionable. This may also include a costing of the business case for different policy options. In many circumstances, clearly communicated evidence can help combat more emotive arguments and misperceptions. | Evidence base | - Externalised expertise (Academia) - Relevance of evidence base |
| 77 | Effective evidence communication may result from improved collaboration between health professionals and planners as each becomes accustomed to different understandings and terminologies. When political windows of opportunity arise, for example, the presence of strong political will and leadership, pre-prepared and well-communicated evidence can then be pushed through. | - Partnership, collaboration, relationships, engagement - Leadership | - Political commitment - Relevance of evidence base |
| 78 | 2018 | - Capability, skills and knowledge - Resourcing of local planning authorities |  |
| 78 | Many practitioners are not using data and insight to design and create healthy places.  Although some practitioners were aware of the evidence base for creating healthy places, we found that only 27% of practitioners are able to access and use local data to identify local priorities when working on placemaking projects | Evidence base | - Use of evidence base - Access to evidence base |
| 78 | Healthy placemaking interventions can be excluded from design proposals due to the perceived cost to implement them.  Practitioners shared their frustration at not being able to implement healthy placemaking interventions as a result of the perceived cost they bring to the overall project. While contrary to the evidence base to support the economic benefits of healthy placemaking, survey respondents felt market pressures meant healthy placemaking is still seen as a luxury rather than a necessity. | Impact of economic focus, viability and costs | - Cost and delays - Impact on implementation |
| 78 | Very few practitioners can demonstrate impact.  Practitioners that we spoke to said they find it difficult to measure impact, caused by a gap in the resources available to them in explaining and demonstrating how to measure the impact of healthy placemaking interventions. | Monitoring and evaluation | - |
| 78 | The systems, policies and processes of planning and building design and development are not currently supportive towards healthy placemaking.  Some practitioners argued that the existing systems, policies and processes do not foster healthy placemaking interventions to be developed as there is a lack of support. Practitioners felt that there are cultural barriers within the workplace that mean they continue producing designs that exclude elements of healthy placemaking | Strength of national policy  Decision-making process | - (Less) focus on process - Governance |
| 78 | Greater understanding is needed about the effect of the built environment on health.  Our research found that the requirements and expectations of national and local politicians to deliver on other priorities (such as housing supply) would often act as a barrier in enabling practitioners to produce health placemaking intervention, while survey respondents felt that the public are not always aware of the effect of the built environment on health. | Complexity of planning regulatory framework | - Political commitment - Role of local communities |
| 78 | Priorities differ across government departments leading to conflict, confusion and no shared vision on healthy placemaking.  Practitioners discussed the challenges they face from various government departments. Some survey respondents reported that they have been incentivised to develop healthy placemaking interventions through working closely with public health professionals as their priorities are aligned with healthy placemaking interventions. However they argued that differing priorities between local government planning departments and highways authorities prevent the interventions from being developed, which compromises design proposals and planning applications in order to gain approval | - Complexity of planning regulatory framework - Decision-making process | - Governance - Lack of alignment - Impact on implementation |
| 79 | Highways, and guidance on highways, make it difficult to create healthy places.  Built environment practitioners reported that they found it difficult to design and develop areas that support health and wellbeing as a result of restrictions placed by highways guidance and highways authorities. | Stakeholders and statutory consultees | Externalised expertise |
| 79 | The vision for healthy placemaking is clear but this vision does not always translate into delivery of projects on the ground.  Director level and senior level practitioners are more open to adopting healthy placemaking interventions, but this vision doesn’t make its way to people working on projects. Data analysis also found that junior practitioners are more likely to experience barriers and therefore feel prevented from creating healthy places, compared to director and senior level practitioners | - Visioning and outcomes-focused - Capability, skills and knowledge | - Impact on implementation - Professionalism and impact on profession |
| 78 | Case studies and exemplars  ‘How to’ guides | Knowledge transfer, good practices | Innovative practices |
| 78 | Encourage and increase use of evidence | Evidence base | Use of evidence |
| 78 | Measure and demonstrate impact | Monitoring and evaluation | - |
| 78 | General principles framework with localised checklists | Use of (local) planning powers | - |
| 78 | Recognising the economic value of healthy placemaking. Social value | Impact of economic focus, viability and costs | - |
| 78 | Education, learning and training  Continuing professional development  Shared learning | Training, learning and development | Professional qualification (knowledge) |
| 80 | 2020 | Capability, skills and knowledge |  |
| 80 | Meanings of sustainability The participants in general understood sustainability to encompass environment, social and economic aspects and described the challenges in seeking outcomes that satisfied all three dimensions. However there was recognition that balance may be an ideal and in reality, there may be pressure to achieve housing, economic or political targets. | Complexity of planning regulatory framework | - Lack of alignment - Professional qualification (knowledge) |
| 80 | This lack of detail and precision in the term was linked to the participants’ view of the core national policy document (National Planning Policy Framework, NPPF) as not useful, despite its stated objective of placing sustainable development as a core construct. The vagueness of critical concepts meant “it’s a lawyer’s dream, because there’s just so much you can interpret and fight the meaning of” | Strength of national policy | Clarity in national policy |
| 80 | Within a planning regime in which planners’ decisions which are unacceptable to a stakeholder can be challenged in court, this implies that the policy is ultimately ineffective because “it lacks teeth, and it means that, I would say, most planners are too scared to really rely on it as something they can resist [a scheme] on | Decision-making process | - Impact on implementation - Confidence - External influence |
| 80 | For the participants who described the NPPF in more positive terms, reference was made to local plans and it appeared that they used the NPPF to ensure compliance but relied more heavily on the local plan. | Use of (local) planning powers | - |
| 80 | In the absence of clear and strong national policy, “it is just down to the planners, at the end of the day, to weigh everything up” [Gail]. Several noted difficulties in implementation, including the contested definition and weak policy noted above, and one described the complexity of attempting to apply a highlevel concept in practice on small projects: “I think climate change is a really difficult thing to consider for an individual planning application | Decision-making process | - Clarity in national policy - Impact on implementation |
| 80 | Planners were not seen as solely responsible: when asked who they viewed as responsible for delivering a sustainable built environment, the interviewees referred not only to central government but also to local government, council partners and specialist advisers such as Highways England. A few argued that everyone involved in the built environment shared responsibility, from citizens submitting a request for planning, to developers to advisers. The responses indicated a role for planners in sustainable development, alongside other stakeholders in the system. | Stakeholders and statutory consultees | - Externalised expertise - Professionalism and impact on profession - Role of private sector - Role of local communities |
| 80 | The evidence was clear that most (though not all) of the participants pursued elements of a sustainability agenda from a sense of professional or personal commitment. They described fighting for greater biodiversity and protection of the green belt, water efficiency, more sustainable homes, fewer cars and eco-towns. This was evident in responses ranging from strong views on the need to reduce reliance on private cars to a more general aim to defend the natural environment where possible. | Visioning and outcomes-focused | - Professionalism and impact on profession - Deliver wider value and benefits |
| 80 | When asked about the applicability of resilience in their work, there was no initial recognition of the concept by almost half of the participants and in a few cases, there was acknowledgement of lack of knowledge on the topic | Capability, skills and knowledge | Professional qualification (knowledge) |
| 81 | With the exception of this participant [Jack], there was little evidence of activity relating to resilience in planning work currently. Participants noted the absence of national policy and the corresponding gaps in local plans. Some also pointed to the challenge in development management of applying the overarching aims of resilience on a single development | - Strength of national policy - Local plan content and plan-making process - Decision-making process - Stakeholders and statutory consultees | - Clarity in national policy (absence) - Impact on implementation |
| 80 | On climate resilience, there is too little knowledge and awareness for the concept to inform day-to-day planning policy and decisions. This holds important implications for the profession, its professional body and policy makers. | Capability, skills and knowledge | - Professional qualification (knowledge) - Professionalism and impact on profession |
| 80 | Current national legislation is by and large unhelpful to planners in delivering sustainability. The legislation should address directly the difficulties of definition and the inherent tensions in aiming for environmental, economic and social sustainability. Consistency in policy is needed which transparently meets national commitments including to COP21 targets, the Climate Change Act (2008) and the UN Sustainable Development Goals | Strength of national policy | - Increase regulations/ powers - Consistency |
| 80 | Development of the built environment is guided by planners’ expertise and long-term view. If planners nationally are unaware of the predicted impacts of climate breakdown on urban and other development, then the built environment now and into the future is not being prepared for future risks. Led by the professional body (RTPI), and drawing on experts within relevant economic sectors and from academe, it is imperative that the predicted risks to the built environment from climate breakdown and principles around urban resilience are disseminated. Resilience thinking must become embedded in day-to-day work of planners | Capability, skills and knowledge | - Professionalism and impact on profession - Externalised expertise - Impact on implementation |
| 80 | Without a bottom-up drive from planners and their representatives, national policy will not develop in the direction needed. | Stakeholders and statutory consultees | - |
| 80 | With adequate legislation, and knowledge of principles and approaches to resilience, the evidence from this study suggests that planners have the motivation, through their professional identity to serve the public good, to deliver a sustainable and resilient built environment. | Visioning and outcomes-focused | Professionalism and impact on profession |
| 82 | 2018 | Capability, skills and knowledge  Resourcing of local planning authorities |  |
| 82 | ■ First, a period of near-continuous change in the planning system over the past decade has compounded rather than resolved the problems these so-called reforms were designed to remedy. | Impact of planning reform processes | - |
| 82 | Second, in place of short-term tinkering we need to take a long, hard look at the fundamentals – the purpose of planning, how it can best be structured, and how all parties can engage most constructively in the planning process | - Visioning and outcomes-focused - Complexity of planning regulatory framework - Stakeholders and statutory consultees | - Deliver wider value and benefits - Lack of engagement |
| 82 | The inadequate state of strategic planning in England | Impact of strategic planning practices | - |
| 82 | The ‘people and planning’ question and the continued disconnect between communities and the planning process | Stakeholders and statutory consultees | Role of local communities |
| 82 | The state of the public planning service | Resourcing of local planning authorities | - |
| 82 | The growing concerns as to whether planning is fit for the future. Is the system ready for the challenges of climate change, inequality, health, biodiversity loss and technological change? | Complexity of planning regulatory framework | - Deliver wider value and benefits - External influence |
| 82 | The key areas of concern raised in the evidence can be grouped into 12 key themes:  ■ the purpose and objectives of the system;  ■ the degree to which the current system is delivering on its objectives, and particularly the kinds of outcomes it produces for people | Visioning and outcomes-focused | Deliver wider value and benefits |
| 82 | how the balance of planning powers should be distributed between central and local government | Use of (local) planning powers | Better use of local levers |
| 82 | the lack of resources and capacity across the public sector planning service and the confusion surrounding the role of the town planner; | Resourcing of local planning authorities | Professionalism and impact on profession |
| 82 | the economic costs and benefits of planning regulation | Impact of economic focus, viability and costs | Cost and delay |
| 82 | Delivering a clear purpose for the planning system  Recommendation 1: A new legal duty to deliver sustainable development in England  Recommendation 2: A cross-sector compact on the values of Planning | - Visioning and outcomes-focused | - Increase regulations/ powers - Governance |
| 82 | Delivering effective and people-centred planning  Recommendation 3: A new kind of positive and powerful Local Plan  Recommendation 4: Local planning authorities that act as ‘master-developers’ to ensure that Local Plans deliver real change  Recommendation 5: Community powers to plan effectively | - Use of (local) planning powers - Local plan content and plan-making process | - Better use of local levers - Role of local communities |
| 82 | Recommendation 10: A duty to local planning authorities to plan for high-quality and affordable home | Use of (local) planning powers | Increase regulations/ powers |
| 82 | Delivering simplified planning structures  Recommendation 12: A smart structure for planning  Recommendation 13: The use of bespoke delivery bodies to deal with long-term planning problems  Recommendation 14: A new Sustainable Development and Wellbeing Act  Aligning the agencies of English planning  Recommendation 15: A re-purposed National Infrastructure Commission  Recommendation 16: An enhanced role for Homes England | Complexity of planning regulatory framework | - Governance - Increase regulations/ powers - Externalised expertise |
| 82 | Encouraging the creative and visionary planner  Recommendation 21: Attracting, training, developing and supporting the necessary numbers of high-calibre planners  Recommendation 22: A requirement for university planning schools to have a social mandate to support basic outcomes for people  Recommendation 23: Increased professional standing for planners, particularly within local government  Recommendation 24: Introduction of a ‘Do no harm’ obligation in built environment professional codes of conduct | Capability, skills and knowledge | - Professionalism and impact on profession - Professional qualification (knowledge) - Increase regulations/ powers |
| 83 | 2010 | Capability, skills and knowledge |  |
| 83 | Evidence Statement 1: Official guidance to spatial planners makes them aware that positive health outcomes should be a strategic objective to be addressed as part of the agenda for sustainable communities. Guidance does not, however, advise them of how this should be addressed; although there appears to be a general expectation that planners should be engaging with professional expertise from other areas to support their evidence and decisions. | - Decision-making process - Stakeholders and statutory consultees | - Clarity in national policy - Externalised expertise - Access to evidence |
| 83 | Evidence Statement 3: It is important for health outcomes to be recognised as a “material consideration” for them to be taken properly into account in planning decisions. Currently, national guidance does not provide this status. However, there is scope within LDFs for explicit policy reference to health outcomes to endow health outcomes with the status of being material to planning decisions | - Decision-making process - Strength of national policy - Local plan content and plan-making process | - Health considerations - Impact on implementation - Clarity of national policy - Better use of local levers |
| 83 | - Evidence Statement 5: National policy guidance to health professionals does not explicitly require them to engage in spatial planning processes. However, it promotes policies which necessitate action on the determinants of health as an effective means of achieving health outcomes, which implies a role for spatial planning as a potential vehicle for effective interventions | Strength of national policy | - |
| 83 | Evidence Statement 6: Government guidance on JSNAs requires them to cover areas of need for health and wellbeing which are potentially influenced by spatial planning decisions. PCTs are jointly responsible for JSNAs and also members of LSPs, which should address the needs identified in formulating SCSs. PCTs‟ involvement in delivering LAAs should require a degree of coordination with LDFs, which are the spatial manifestation of SCSs. Implicit in these arrangements is a need for the decision-making processes for JSNAs, SCSs, LAAs and LDFs to be coordinated, and hence for an engagement between health and spatial planning. This presents a particular challenge of communication and coordination in twotier local authority structures, where the LAA relates to the county area, and spatial planning is the responsibility of boroughs and districts | - Complexity of planning regulatory framework - Partnership, collaboration, relationships, engagement - Strength of national policy | - Non-planning regulations - Lack of alignment - Lack of engagement - Governance |
| 83 | - Evidence Statement 7: The current set of RSSs provides ample evidence of health being on the agenda of spatial planning at that level. Treatment of health is, however, far from uniform, and particularly with respect to any inclusion of policies setting out expectations for health to be addressed in the subordinate LDFs. Where such policies are included, they do not provide clear guidance on how health issues are to be addressed. The recent Government announcement of an intention to abolish the regional level of spatial planning has seriously reduced the relevance of these findings. However, they do indicate how health might be handled in the future if there were any higher-order strategies able to influence local spatial planning. | Impact of strategic planning practices | Health consideration |
| 83 | Evidence Statement 8: HIAs undertaken on RSSs might have been considering the impact of the intended outcomes of policies, and not the impact of the policies themselves. This may have undermined decisions on policies which relied on the HIAs. | Use of impact assessment | Health impact assessment |
| 83 | Evidence Statement 9: The sample of LDFs demonstrates that the pursuit of health outcomes has been accepted as a legitimate issue to be addressed in local spatial planning. Health features both explicitly and implicitly in objectives and policies. Promoting healthier communities was a theme found in both prosperous and deprived areas, and better access to healthcare provision was a common objective or policy. While most LDFs acknowledged that policies and other factors impact on health, the implied causal links between policies and outcomes were generally neither strong nor explicit. The respective PCTs appeared to have been engaged to varying degrees in core strategy preparation but relating to the practicalities of providing facilities, rather than to policies promoting healthier lifestyles or addressing health inequalities. | Local plan content and plan-making process | - Access to evidence - Lack of engagement |
| 83 | - Evidence Statement 12: Spatial planners‟ policies to address health outcomes appear to have but a weak foundation in specific knowledge and understanding of how they can influence the determinants of health. Many spatial planners would welcome better information and guidance on how to address health outcomes, and particularly if this were embodied within formal national policy guidance. This was less the case for transport planners, who appeared much more comfortable and confident that guidance provided a sound basis for taking health fully into account in their decisions. There was little evidence of planners being either aware of guidance from NICE and the DH, nor any belief that this would be of relevance to them. | Capability, skills and knowledge | - Clarity in national policy - Confidence - Relevance of evidence base - Access to evidence base |
| 83 | C- Evidence Statement 13: Despite the requirement for LDFs to comply with RSS policies, local priorities and not RSS expectations appear to be the main driver in directing LDF policies towards health outcomes | Local plan content and plan-making process | - |
| 83 | - Evidence Statement 14: Local sustainable community strategies (SCSs) normally exert a strong influence on the broad objectives and priorities to which local spatial plans are directed, and this has been one of the key reasons for health outcomes being picked up in LDFs. Nonetheless, the impact of SCS prioritisation for health appears to be limited when little guidance is provided regarding what role and outcomes are actually expected from spatial planning. There appears sometimes to be an even stronger disconnection between the SCS of a county and the spatial planning being undertaken by the lower-tier planning authorities. This is an issue which is only partially addressed through the latter‟s own SCSs, particularly if the main focus of the respective PCT‟s engagement in local strategic partnerships (LSPs) is at county level | Complexity of planning regulatory framework | - |
| 84 | - Evidence Statement 15: PCTs do not normally view engagement in spatial planning as core business. In organising their staffing structures around core business, some PCTs were found to have inadvertently created situations in which no member of staff recognised a responsibility for liaison with spatial planners. This was particularly marked where two-tier local government structures led to the PCT‟s focus lying in cooperation with the county council as social care authority, while spatial planning is a district function | Partnership, collaboration, relationships, engagement | - Lack of engagement - Dedicated posts and capacity (none) - Governance |
| 84 | - Evidence Statement 16: An extremely varied knowledge of spatial planning was found among health professionals, even among individuals designated as responsible for liaison with the district authorities in their spatial planning function. The better working relationships between PCT and planning authority tended to be found where responsibility for this in the PCT involved someone with a good understanding of spatial planning | Capability, skills and knowledge | - Professional qualification (knowledge) - Dedicated posts and capacity |
| 84 | Chapter 6: Interaction between Health Professionals and Planners - Evidence Statement 18: The case studies revealed a very wide range of different arrangements for collaboration between PCTs and local authorities. Although some were specifically designed to facilitate health input into spatial planning, it was far more common that this was but one part of more broadly-based arrangements. No strong correlation was found between the type of arrangement adopted and success in integrating health into planning, and factors such as political priorities and the knowledge of individual practitioners‟ appear to be of at least equal importance. In particular cases the creation of a public health capability within a local authority‟s structure provides a ready health input into spatial planning, potentially avoiding reliance on resources being made available from within the PCT‟s structure. - Evidence Statement 19: Where health professionals are engaging in spatial planning, the representatives of the two professions appear to be learning about one another‟s cultures, and hence reducing the scope for misunderstanding and friction between one another. Nonetheless, there is still some way to go: some differences in language, culture and approach remain, and hence issues may yet arise. - Evidence Statement 20: Although several practitioners made reference to there being problems of cooperation between health and planning relating to their respective time horizons, little evidence was found to support this being a real issue | Capability, skills and knowledge | - Governance - Dedicated posts and capacity |
| 84 | Chapter 7: Matters for Consideration if Spatial Planning is to aim at Health Outcomes  - Evidence Statement 21: Unless there is real political prioritisation of health outcomes among members, a council‟s approach to health in spatial planning is likely to be little more than tokenism. This is irrespective of apparent prioritisation in a SCS. Conversely, if there is real political backing for the pursuit of health outcomes, it is possible to initiate a comprehensive corporate approach to the pursuit of health that should automatically incorporate spatial planning as an intrinsic element. | Impact of politics and politisation in planning | - Political commitment - Governance |
| 84 | - Evidence Statement 22: Planners are manifestly conscious of the expectation that their policies are based on sound evidence, particularly the need for them to stand up to challenge at a public enquiry or examination. Nonetheless, they appear to be quite content to pursue policies promoting health outcomes despite the lack of specific evidence that the policies in question will themselves lead to better health. They acknowledge that policies which lead to greater provision with open space, cycling routes and safe pedestrian routes will not automatically produce healthier behaviour among the target population. At the same time, they clearly recognise the evidence that more use of these facilities should improve health, and therefore trust that others‟ actions complementary to their policies can help the population to make healthy choices in their behaviour | Evidence base | - Access to evidence - Use of evidence base |
| 84 | - Evidence Statement 23: JSNAs are generally not satisfying the needs of planners for data inputs to their processes, and they are not providing a solid foundation for joint working with PCTs. While some planners are disappointed by their lack of involvement in Joint Strategic Needs Assessments (JSNAs), others had never heard of them. Evidence was offered in the studies suggesting that health outcomes had hardly ever been used as grounds for refusing planning permission. Examples were also provided of the difficulty encountered when seeking to use developer contributions for health facilities. These problems reflected the difficulties encountered in establishing robust evidence linking health impact to a particular development, and also the unanswered question of when health constitutes a material consideration in planning decisions | Evidence base | - Non-planning regulations (health levers) - Health consideration - Impact on implementation - Relevance of evidence base - Lack of engagement |
| 84 | - Evidence Statement 24: Planners generally believe that health outcomes have hardly ever been used as grounds for refusing planning permission, and that to seek to do so would probably result in failure. They also report problems encountered when seeking to use developer contributions for health facilities. These limitations on action reflect partly the difficulty of establishing robust evidence that links specific health outcomes to any particular development. They also follow from serious doubts whether health would be considered at appeal to be a material consideration in a planning decision. Related to this, there appears to be a common belief that, if national planning guidance were formally to establish health as a material consideration, this would both remove these doubts and also enable evidence of health impact to be handled on the basis of reasonable probability rather than absolute proof. | Decision-making process | - Health consideration - Impact on implementation - Relevance of evidence base - Clarity of national policy |
| 84 | - Evidence Statement 26 The use of HIAs is not universal, reflecting varying perceptions of whether the cost and time involved can be justified by the benefit. An example was found of “mini-HIAs” being developed - a simpler process requiring far less resource - which might offer a more beneficial approach. | Use of impact assessment | - Health impact assessment - (Less) focus on process |
| 84 | Evidence Statement 27: A growing interest was found in the local development of supplementary planning guidance within a LDF to entrench health issues more firmly into development management. This is seen as presenting an opportunity to provide much more detailed guidance than in a core strategy. Significantly, it is also recognised as presenting an opportunity to establish health as a material consideration in planning decisions, even without national guidance. | Use of (local) planning powers | - Better use of local levers - Impact on implementation - Health consideration |
| 84 | Evidence Statement 28: There appears to be very little meaningful monitoring of spatial planning policies which are aimed at health outcomes: the implementation of policies is not always being monitored, nor the direct effect of the policies in terms of their impact on health in accordance with what was intended. Generally, only broad health indicators are being monitored - primarily those used for LAAs – and planners appear to be well aware that these cannot be linked directly to the effects of their policies. | Monitoring and evaluation | - |
| 85 | 2017 | Capability, skills and knowledge  Resourcing of local planning authorities |  |
| 85 | Shared vs sole responsibilities  DsPH and planners had different conceptions of who is responsible for public health. All the DsPH believed in a broad, shared responsibility, citing various actors including the council, planning, the national government, employers and individuals. When planners were asked about responsibility for obesity, physical activity and community nutrition, they saw a clearer link between planning and physical activity than with community nutrition, primarily citing sustainable transport, path provision and recreation, though one identified takeaways. One planner said, ‘At the end of the day, it’s down to the individual to address their obesity (Planner 4)’.  These differences in views on responsibility for health would suggest a difference of understanding amongst some in the local government about the complex causes of obesity and how to address inequalities. However, one planner did point out that it is health professionals’ responsibility to ‘make the link’ between planning and health ‘because I don’t think people from planning would start from a health improvement stance’. This has implications for leadership and leadership roles | - Visioning and outcomes-focused - Leadership | - Lack of alignment - Professional qualification (knowledge) |
| 85 | Joined up vs fragmented practice DsPH were able to provide many examples of how they are joining up with various organisations and taking whole systems approaches in efforts to promote healthy lifestyles for their communities. For community nutrition, working with environmental health officers, one local government provided health awards for local education authorities, nurseries, schools and takeaway restaurants. It also provided a range of community pro- jects such as urban farming and allotments, specifically in deprived areas and with people with mental health issues. Planners indicated working with other organisations, but to a lesser extent. | Partnership, collaboration, relationships, engagement | - Lack of alignment - Governance - Externalised expertise |
| 85 | Relationship building was identified as key to bridging the divide between planning and public health. Planner 1 felt that planning was ‘reactive’ when it came to incorporating public health into their work, but was starting to see signs of planning being more ‘proactive’, with reference to using the Takeaway Toolkits (Greater London Authority 2012) for example | Partnership, collaboration, relationships, engagement | - |
| 85 | DsPH reported more and intense experiences of changes since the restructuring of the NHS in April 2013, whereas planners seemed to have experienced changes more due to the introduction of the National Planning Policy Framework (Department for Communities and Local Government 2012) in March 2012. DsPH also did not notice a major change in planners’ responsibilities as a result of the restructuring, though again, planners demonstrated better awareness of their potential to impact health and well-being, especially around licensing of alcohol and food outlets. | Impact of planning reform processes | - Impact of public health reforms - Better use of local levers - Professional qualification (knowledge) |
| 85 | DsPH identified ‘huge economic constraints’ as a result of the restructuring, for example cutting back on spending in park maintenance, and actually expecting ‘neighbourhoods to look after those kinds of spaces themselves … to find the drivers in the community for self-management’ (DPH 2). | Resourcing of local planning authorities | - Funding - Role of local communities |
| 86 | Planners looked for evidence to support policy-making (e.g. case studies of good practice). They would consult the Planning Policy Guidance Notes, the Local Government Association (LGA 2017) and local ‘statistics’ on obesity. Some planners explained that they did not feel confident in the area of community nutrition and physical activity, and expressed little if any interest in obtaining further training in these areas, indicating they would contact individuals who did know. Planner 2 for example felt the integration of public health into local authorities had helped raise awareness in these areas, and DPH 1 confirmed this observation | Capability, skills and knowledge | - Type of evidence base - Access to evidence base - Confidence - Impact of public health reforms |
| 86 | All participants appeared to use personal contacts for their main sources of evidence. DsPH consulted col- leagues in leadership roles such as other DsPH and man- agers, those in roles responsible for keeping up to date with evidence and those who take a particular interest, looking for information on planning rules around spatial planning, how national policy determines local policy. DsPH kept up to date by consulting public health profiles, public health websites (e.g. NICE, PHE), pub- lic health intelligence services and sharing good prac- tice with colleagues outside of the region. Most DsPH seemed confident in the areas of community nutrition and physical activity, often citing evidence through the interviews, e.g. the role environments play in health; how to reduce gaps in inequalities. For example, DPH 1 felt their staff ‘got’ issues of obesity, so rather than  training, they required the means to implement the evidence base. | Evidence base | - Access to evidence base - Use of evidence base - Type of evidence base |
| 86 | Conflicting vs shared priorities  DsPH recognised that planners have large pressures in other areas besides health, such as concerns for sustainability and climate change. Economic regeneration as a driver in urban development is understandably prioritised in the north of England. Economic regeneration, however, can run counter to health priorities in areas desperate for any inward investment and development. DPH 2, for example, explained how in their area the local government was allowing the development of new properties in more affluent areas, which provided more profit for developers, but widens inequalities as the housing shortage is still a major issue in the more deprived areas. Planner 4 raised a similar issue regarding the establishment of new takeaways, whereby such development was ‘more to do with retail rather than health’. For example, in this case, despite being within 400 m of a school, planning inspectors approved the development of takeaways because they demonstrate investment and job creation. | Impact of economic focus, viability and costs | - Lack of alignment (priorities) - Private sector investment |
| 86 | Formal vs organic responsibilities Planners are able to refer to the National Planning Policy Framework to change policy regulations and practice in relation to public health and obesity prevention, but it is considered ‘very high level and not very easy to interpret’. One planner cited the development of a Supplementary Planning Document to provide the ‘how to’ details. Speaking about the need for health assessments for individual planning applications, one senior planner said:… | - Strength of national policy - Use of impact assessment | - Clarity in national policy - Better use of local levers |
| 86 | Rather than providing training for planners on health and obesity, DPH 3 believed public health needs to connect better with planners and build relationships by which the two areas can innovate new approaches together. DPH 1 identified a ‘critical leadership role put- ting it all together’. | - Knowledge transfer, good practices - Leadership | - |
| 86 | The DsPH also appeared to think outside of their daily professional practices and towards larger, more long- term changes. DPH 1 said on the role of planning with regard to public health, ‘it’s one where there’s a lot of potential, but with one or two honourable exceptions in different parts of the country, untapped potential’. DPH 3 said: I think it’s useful to have a kind of whole area approach… I get the impression if you had a local champion that was really passionate about all this would probably achieve more than any number of kind of well-meaning strategies and policies … Some of it is about opening people’s eyes to what’s possible, isn’t it? | - Capability, skills and knowledge - Knowledge transfer, good practices | - Professionalism and impact on profession - Innovative practices |
| 85 | Given participants identified much value in organic modes of working together, this would require increased capacity including leadership (champions were cited). While the importance of ‘boundary-spanning’ leader- ship has been identified (Hunter and Perkins 2012) and reports have suggested strong leadership in this area is needed (Town and Country Planning Association 2016), our research suggested that such leadership was still lacking | Leadership | Governance |
| 85 | However, again, given budget cuts to local authorities in England, the issue may be a lack in leadership skills and capacity to carry out these less formalised, ‘non-core’ aspects local government staffs’ roles. Similarly, with respect to the anticipation that the relocation of public health to local government in England could improve impact, the current austerity measures somewhat limit local government ability to fund preventative measures, e.g. maintaining green spaces (Hall 2015). In some countries, public health may have always been within local government, but there is a silo between the professions. | Resourcing of local planning authorities | - Funding - Lack of alignment - Impact of public health reforms |
| 85 | Political will is also critical. In France, the EPODE programme (the largest global childhood obesity prevention programme) observed that local authorities needed to engage with the programme on a voluntary basis and there needed to be local political will for success (Borys et al. 2012). However, for progress to be made, not only do individuals and civil society have a role but regulatory action from governments is required (Roberto et al. 2015). Roberto et al. (2015) describe examples where the ‘Health in all Policies’ approach has been used including South Africa, Southern Australia and Victoria (Australia). However, they conclude that despite increased attention to health and obesity, the response is not adequate | - Impact of politics and politisation in planning - Strength of national policy | Impact on implementation |
| 85 | Based on our findings, we recommend health and well-being be explicit learning outcomes of planning education, in England, the U.K. and internationally. In 2010, the RTPI undertook consultation on planning education and this was made by a number of senior planning academics (Townshend 2010). One DPH in the present study pointed out that planners do not need to be experts in public health, but they do need to be aware of their contribution. The Town and Country Planning Association (2016) suggest that local authorities provide shared training across public health and planning as well as offer secondments | Training, learning and development | - Professional qualification (knowledge) - Deliver wider value and benefits |
| 85 | Future research could focus on these areas of leadership identified by the McKinsey Institute (2014), as well as an exploration into the extent to which public health leaders use the full suite of influences available to them. For example, they might re-profile public health budgets away from orthodox intervention treatments into supporting policies that affect the wider determinants of health; this could include incentivising and financially supporting planning teams | - Capacity of local planning authorities - Leadership | - Better use of local levers - Funding - Non-planning regulations |
| 88 | 2019 | Resourcing of local planning authorities  Capability, skills and knowledge |  |
| 88 | Local plans are required by planning legislation and should be the key way that local authorities demonstrate how they will help meet the need for new homes in their areas. However, local authorities are often under-resourced and under-staffed and struggling to produce plans as they can be technically complex, time consuming and resource intensive | - Resourcing of local planning authorities - Complexity of planning regulatory framework | Cost and delay |
| 88 | As of December 2018, only 143 (42%) of local authorities had an up to date local plan, 149 (44%) had a plan that was more than five years old, and 46 (14%) had no plan at all. Despite these significant gaps, the Department has made limited use of its powers to intervene in local authorities who have not produced a local plan. In November 2017, the Secretary of State wrote to 15 of the local authorities who did not have a local plan and in January 2019, made more direct interventions in two local authorities. But these figures barely scratch the surface of the significant number of local authorities which either have no plan at all or a very old one. | Local plan content and plan-making process | Up to date local plan |
| 88 | Recommendation: By the end of 2019, the Department should write to us detailing what additional interventions it will make when local authorities fail to produce local plans. These interventions should include a range of ‘carrot and stick’ measures of support and penalties. | Local plan content and plan-making process | - Impact on implementation - Increase regulations/ powers |
| 88 | The time taken by the Planning Inspectorate to determine housing appeals, which increased from 30 to 38 weeks in the five years from 2013 to 2018, is delaying the building of new homes, hampering progress on targets and creating uncertainty for local authorities and communities. The Department knows the Inspectorate needs to significantly improve its performance. In June 2018, the Secretary of State commissioned a review of how the Inspectorate deals with appeal inquiries. The Department expects the Inspectorate will develop an action plan promptly in response to the review’s findings and has also agreed a performance recovery business plan of £13 million to help the Inspectorate improve | Decision-making process | - Planning Inspectorate - Cost and delay - Uncertainty - Impact on implementation |
| 88 | Recommendation: By the end of 2019, the Department should set out for us detailed actions and milestones for the Planning Inspectorate’s performance improvements across the full range of all its services | Decision-making process | Planning Inspectorate |
| 88 | Local authorities may lack the skills to negotiate contributions from developers through section 106 agreements and there is little transparency of these negotiations. In two tier authorities there is the added risk of insufficient co- ordination between them to achieve the maximum contribution | - Capability, skills and knowledge Partnership, collaboration, relationships, engagement | - Commercial and development economics skills - Governance |
| 88 | Local authorities can also use the Community Infrastructure Levy to get contributions from developers, but as of January 2019 only 47% of local authorities had implemented the Community Infrastructure Levy. Implementing the Community Infrastructure Levy is complex, time consuming and yields small returns in areas of low land value. The Department is aware of these shortfalls and introducing and consulting on several reforms to section 106 and the Community Infrastructure Levy which aim to simplify the process, bring more transparency and help prevent developers reducing contributions using viability arguments. Some of these reforms will need legislative changes and could take several years to be put in place. | Complexity of planning regulatory framework | - Private sector investment - Focus on process - Increase regulations/ powers |
| 88 | There are concerns about poor quality of the build of new homes and that of office accommodation converted into residential accommodation through permitted development rights. The Department is focusing on the quality and safety of high-rise residential buildings after the Grenfell fire. It does not have a specific programme to address concerns about the quality of new builds. It has some initiatives which aim to improve the quality of design of new homes, including revising the Department’s design guide, although these do not address the quality of the final build. | - Impact of planning reform processes - Strength of national policy | Clarity in national policy |
| 89 | 2021 | Resourcing of local planning authorities |  |
| 89 | We are concerned about the lack of detail in respect of the proposed reforms to the planning system, which has made it very difficult to assess the possible practical implications of many of the reforms. The Government should consult on the details of proposed reforms to prevent unintended consequences and harms resulting from them. Given the complexity of the issues, and the possibility that its contents will differ from the proposals contained in the White Paper, the Planning Bill announced in the Queen’s Speech should be brought forward in a draft form, and be subject to pre-legislative scrutiny | Impact of planning reform processes | - Impact on implementation - Uncertainty |
| 89 | We welcome the Government’s proposal that having an up to date Local Plan should be a statutory requirement on local authorities. We also welcome the proposal that Local Plans should be more focused and shorter. But we do not agree that the 30-month timeframe proposed for the development of Local Plans is enough to ensure high quality | Local plan content and plan-making process | Up to date local plan |
| 89 | The Government must ensure that statutory consultees have time to comment on Local Plans. The Government should consider a staggered roll-out of the new types of Local Plans across the country. It should be permissible and straightforward to undertake quick updates of Local Plans every two years, including with appropriate time for public consultation. The Government should consider the case for confirming that the National Grid is a statutory consultee in new Local Plans. (Paragraph 45) | Stakeholders and statutory consultees | - Up to date local plan - Externalised expertise |
| 89 | Increasing the speed at which Local Plans are developed and updating them will be resource hungry. The Government needs to clarify how such needs can be met and what resources will be applied to local authorities to enable them to achieve these ambitious timescales | Resourcing of local planning authorities | Cost and delay |
| 89 | The duty to cooperate between local authorities has operated imperfectly. However, we heard strong agreement there needed to be more cooperation between local authorities and that sub-national planning was a weakness of the current system. The Government should only abolish the duty to cooperate when more effective mechanisms have been put in place to ensure cooperation. Whilst the duty to cooperate remains in place, the Government should give combined authorities the statutory powers to oversee the cooperation of local authorities in their area. Longer-term reforms could include greater use of joint plans, of plans overseen by mayors and combined authorities, and of development corporations. The Government should seek to apply the lessons from successful strategic plans devised by local authorities in certain parts of the country in devising more effective mechanisms for strategic planning. | Impact of strategic planning practices | - Lack of engagement - Increase regulations/ powers - Governance |
| 89 | The Government must commission research about the extent of public involvement in the planning system. This should precede the collection from local authorities and publishing of statistics about public involvement in Local Plans and in individual planning applications. Such research would give a clearer picture of the current situation and, in particular, at which point in the process people are most engaged. (Paragraph 76)  11.We support enhancing public involvement with Local Plans. However, figures cited by the Minister suggest that far more people are involved at the point when individual planning applications are considered than at the local plan stage, and this was backed up by the evidence we have received. We also fear that people will resort to legal measures if they cannot comment upon and therefore influence an individual planning proposal. Therefore, all individuals must still be able to comment and influence upon all individual planning proposals. (Paragraph 77) | Stakeholders and statutory consultees | - Role of local communities - Lack of engagement |
| 89 | It is disappointing that local councillors were not mentioned in the White Paper. They have a key role to play in both Local Plans and individual planning applications We recommend that the Government set out how the valuable role of local councillors will be maintained in the planning system (Paragraph 78). | Impact of politics and politisation in planning | Role of elected members |
| 89 | 13.We welcome the greater use of digital technology in the planning system. But we recognise the need to ensure those lacking access can know about and participate in the planning process. The Minister suggested that the existing statutory notices on local newspapers and on lampposts would become a matter of discretion for local authorities. We do not agree with this approach. It risks creating a postcode lottery as to whether such notices continue. This would disadvantage those residing in financially stretched councils and those moving into local authorities where such practices have been discontinued. The existing statutory notices should be retained for all local authorities, to be used alongside technology. We propose the use of virtual participation in planning meetings continue alongside in-person meetings after the COVID-19 restrictions have been lifted. We also propose that local authorities should experiment with novel ways of engaging the public with the wider planning system, for instance through the use of citizens assemblies. (Paragraph 88) | Stakeholders and statutory consultees | - Access to evidence base - Innovative practices |
| 89 | The housing formula  14.We support the principle of using a standard method that applies across the country. We recognise there has been criticism of the current standard method for not promoting levelling up by reducing the targets for future homes below the numbers currently being delivered. It also does not directly consider brownfield sites nor environmental and other constraints on developable land in a particular area. (Paragraph 110)  15.We think the Government’s abandonment of its proposed formula for determining housing need is the correct decision. There remains a need for additional information about how the Government’s revised approach, announced in December 2020, might work in practice. This is especially important given the proposed urban uplift for 20 urban centres The Government should: | Strength of national policy | Clarity in national policy |
| 90 | 18.It is our view is that the pace of completing planning permissions is too slow, and that carrots and sticks are needed to quicken the pace. The Government should produce a strategy for increasing the extent of multi-tenure construction on large sites in line with the Letwin Review’s recommendations. It should explore the greater use of Development Corporations that are transparent and accountable, alongside incentivising the use of smaller sites and SME builders. We also recommend introducing, in the first instance, time limits for the completion of construction and non-financial penalties where those limits are exceeded without good cause. The Government should set a limit of 18 months following discharge of planning conditions for work to commence on site. If work has not progressed to the satisfaction of the Local Planning Authority then the planning permission may be revoked. An allowance of a further 18 months should be allowed for development to be completed, after which the local authority should be able, taking account of the size and complexity of the site, and infrastructure to be completed by other parties, to levy full council tax for each housing unit which has not been completed. (Paragraph 129) | Decision-making process | - Better use of local levers - Role of private sector - Increase regulations/ powers |
| 90 | 19.We support ensuring that the additional housing being built includes affordable and social housing. There should also be support and encouragement for local authorities to deliver specialist housing, particularly for elderly and people with disabilities. The Government should create a C2R class for retirement communities to ensure clarity in the planning process. There should be a statutory obligation that Local Plans identify sites for specialist housing. We repeat our recommendation in our 2020 social housing report that the Government should publish annual net addition targets for the following tenures over the next ten years: social rent, affordable rent, intermediate rent and affordable homeownership. (Paragraph 136) | Local plan content and plan-making process | Increase regulations/ powers |
| 90 | 20.We heard concerns about the Government’s First Homes programme, especially its potential impact on the provision of other forms of affordable housing. First Homes has an important part to play in delivering homeownership, and we hope that the Government has learnt the lessons of the failure of the Starter Homes programme and the need for the 25% price reduction to remain in perpetuity. But the Government must also ensure that its First Homes programme does not reduce incentives for other types of affordable housing—in particular the delivery of shared ownership properties or social housing. We recommend that the Government lay out its timetable for when First Homes will become available. To reflect the needs for different types of affordable housing in different areas, local authorities should have discretion over what proportion of houses built under Section 106 agreements must be First Homes. (Paragraph 139) | Impact of planning reform processes | - |
| 91 | Omissions  22.We agree that the Government’s proposals omitted important issues that should be considered in any changes to the planning system. This was particularly true of the lack of consideration of non-housing issues. Different aspects of the planning system cannot be compartmentalised in this way. Housing cannot be treated in isolation from wider infrastructure, economic, leisure, and environmental activities and considerations. Therefore, in advance of a Planning Bill, the Government should include within consultations the expected impact of its proposed reforms to the planning system on:  The ‘levelling up’ agenda including the promotion of employment  The economic recovery from the COVID-19 pandemic  The high street  Addressing climate change and creating sustainable development  Bolstering sustainable transport  The delivery of commercial and industrial property, including leisure facilities, mineral extraction, and energy networks  Policies on social exclusion and on particular groups including Gypsy and Traveller Communities  The environment—in particular the proposed reforms to environmental impact assessments, the designation of protected areas and species, and the proposals for a net gain in biodiversity in the Environment Bill currently going through Parliament (Paragraph 148) | - Visioning and outcomes-focused - Strength of national policy | Deliver wider value and benefits |
| 91 | Land capture and the funding of infrastructure  23.We were disappointed that very little progress has been made in implementing the recommendations of our predecessor committee’s report into land value capture. The Government’s response to our social housing report did not engage with our renewed recommendations about reforming the Land Compensation Act 1961, and the promised consultation in the response for autumn 2020 has not appeared. We call upon the Government to act upon the whole range of recommendations in our predecessor committee’s Land Value Capture report. (Paragraph 154)  24.The Government must clarify how it will replicate the binding nature of Section 106 agreements and which parts of the approach will be retained. If they cannot be easily replicated, especially without creating additional complexity, then we recommend retaining Section 106 agreements. (Paragraph 161) | Impact of planning reform processes | Private sector investment |
| 91 | Resources and skills  26.There is a clear need for additional resources for local planning authorities and this was reflected in evidence from a wide range of sectors. The reduction in their funding is slowing down the workings of the planning system. The Government’s proposed reforms will require additional specialist skills, for example in areas such as design, on top of the existing resource pressures faced by the planning system. The Royal Town Planning Institute estimated that £500 million over four years was needed in additional funding. We therefore welcome the additional funding provided at the Comprehensive Spending Review, and the Minister’s assurance that this is only the start. The pressures on the system will only increase if the Government proceeds with its reforms, including the thirty-month timeframe for Local Plans, at the same time as LPAs have to continue to operate the current system. The Ministry should now seek to obtain a Treasury commitment for an additional £500 million over four years for local planning authorities. Providing this certainty of funding should precede the introduction of the Planning Bill. (Paragraph 185)  27.The Government’s reforms require an increase in planning staff, especially those with specific specialist skills, such as design. These skills gaps will need to be filled if the planning system is to be improved. The Government must undertake and publish a resources and skills strategy in advance of primarily legislation, to clearly explain how the various skill needs of the planning system will be met. (Paragraph 186) | - Resourcing of local planning authorities - Capability, skills and knowledge | - Funding - Cost and delay - Professional qualification (knowledge) - Certainty - Impact on implementation |
| 91 | Design and beauty  28.The Government’s focus on beauty, whilst laudable, must not detract from other important aspects of design. The Government must ensure that its national design code, advice for local authorities about local design codes, and other aspects of design policy reflect the broadest meaning of design, encompassing function, place-making, and the internal quality of the housing as a place to live in, alongside its external appearance. Given the problems with defining beauty, and to ensure a wider approach to design, there should also not be a ‘fast track for beauty’ | Visioning and outcomes-focused | - |
| 91 | Many discussions about beauty and design are very localised, concentrating a specific site, building or street. We do not think these discussions can be incorporated into Local Plans covering an entire local authority. Therefore, the Government must clarify how the public will be able to offer views about developments at this small scale. This is doubly significant given the Government’s proposed reduction in the opportunities for people to comment on individual planning proposals. (Paragraph 203 | - Stakeholders and statutory consultees - Local plan content and plan-making process | Role of local communities |
| 92 | 2020 | Capability, skills and knowledge  Resourcing of local planning authorities |  |
| 92 | Stakeholders were influenced indirectly by academic research, which informed national dialogue and organisational concern about levels of physical inactivity and health impacts. Participants generally understood that there is strong evidence of health benefits of physical activity, which they described as ‘common sense’. | Evidence base | Access to evidence base |
| 92 | Health Impact Assessments’22 conducted by developers were often not required in local planning policy or were reportedly weak due to lack of skills and enforcement mechanisms | Use of impact assessment | - Health impact assessment - Professional qualification (knowledge) - Impact on implementation |
| 92 | Evidence for a solution—knowing what works  Evidence for solutions to identified problems or needs was available within guidance material, based on academic evidence from evaluations and case studies, for example from Public Health England and the Town and Country Planning Association.23,24 This was particularly accessed by urban planners, developers and public health practitioners who understood the value of ALI for health and wanted workable solutions. However, some developers complained that health evidence struggled to reach non-health sectors and one transport planner described guidance for cycling infrastructure as ‘sporadic’ and ‘ad hoc’. | Knowledge transfer, good practices | - Type of evidence base - Access to evidence base - Relevance of evidence base |
| 92 | Sometimes health benefits of ALI were used to justify decisions post hoc. For example transport planners, who prioritised tackling congestion, acknowledged health benefits of walking and cycling infrastructure to support such investment over roads; developers justified spending on greenspaces to investors with research about impact on house prices,25 and sometimes used health evidence to justify less road construction, which was expensive, affecting profits | Decision-making | - Health consideration - Use of evidence base |
| 92 | Limitations of evidence  A lack of clear evidence of ALI impacts made it difficult for public health practitioners and developers to know what to promote. Urban planners focused on outputs rather than outcomes, for example that the construction of cycle routes was completed rather than whether routes would be well used. Councillors were reluctant to try new designs based on examples from other places, which did not appear contextually relevant, and were fearful of seemingly wasting resources on apparently ‘risky’ solutions, which could be politically damaging. This was particularly a problem where good-practice demanded a step change in quality from the status quo and opposition from car drivers or restricting house building were concerns. | Evidence base | - Relevance of evidence base - Role of elected members |
| 92 | Developers were also reluctant to invest in walking and cycling infrastructure in areas with apparent low local demand because they did not believe it would increase house prices.  Economic effects of ALI were rarely considered because financial savings from health benefits of ALI did not directly affect local government budgets; therefore, many councillors were sceptical of its value. Also, cost–benefit analysis was difficult to use in the planning system because urban planners negotiate financial contributions from developers, without monetising potential benefits. | Impact of economic focus, viability and costs | - Private sector investment - Use of evidence base |
| 92 | Influential individuals  Public health stakeholders could be influential, firstly as knowledge brokers sharing evidence about the health effects of ALI and providing practical solutions, but potentially also acting as leaders, building strong relationships to inspire decision-makers to raise up health in their consciousness and motivate them to argue for ALI. Where public health practitioners had a defined planning role, urban planners described them as ‘passionate’ and a ‘force of nature’ and participants explained that they broke down silos to motivate stakeholders across sectors, creating mutual benefits with other sectors’ outcomes, including air quality, noise, flooding, biodiversity, congestion, social cohesion, crime and house prices.  Urban planners met most regularly with developers and negotiated with multiple stakeholders who were said to push their own agendas. ALI could be difficult to achieve because of other demands and no defined minimum standards, but urban planners could influence designs if knowledgeable and motivated; however, they lacked specialist health understanding | Capability, skills and knowledge Leadership | - Access to evidence base - External influence - Deliver wider value and benefits - Dedicated posts and capacity - Professional qualification (knowledge) |
| 92 | The value of early involvement  Most stakeholders understood that early engagement with developers, before planning applications were submitted, provided the greatest opportunity to influence ALI designs, and some were frustrated that LA urban planners involved them too late. It therefore appeared that LA urban planners needed to either understand the health impacts of a scheme themselves, which they struggled with, or be able to bring in other sources of knowledge and influence via public health practitioners. | Stakeholders and statutory consultees | - Lack of engagement - Frontloading/ upstream - Health considerations - Access to evidence base - Externalised expertise |
| 92 | Limited by policies  Stakeholders discussed a lack of national level standards and policies for ALI, which restricted quality. Participants said that local policies generally supported healthy developments but wording was vague without specifications for walking and cycling infrastructure and only quantities of open space required per population, not quality. Stakeholders described tensions between ALI and competing demands, including national planning and transport policies, which promoted house building,26 and transport assessment methods, which focused on road traffic analysis rather than ‘fluffy active travel stuff’ (LA transport planner). | Strength of national policy | Clarity in national policy (lack) |
| 92 | So whilst some planners and developers wanted to be innovative, they were restricted by local policies, for example, specifying a minimum number of car parking spaces per house | Knowledge transfer, good practices | - |
| 92 | It seemed that local policies were important to set minimum standards for developments, which LA urban planners could then use to hold developers to account. Without defined policies, stakeholders said developers would only provide the minimum that they could get away with, unless they saw financial value in doing more.  Participants talked about difficulties in producing policies, which risked being unpopular to car drivers as councillors feared public backlash if congestion increased as a result of new development. So whilst some planners and developers wanted to be innovative, they were restricted by local policies, for example, specifying a minimum number of car parking spaces per house. | Impact of economic focus, viability and costs | - Better use of local levers - Risk management |
| 93 | Watering down good designs  Even when ALI was initially well designed, participants described situations where plans could later change because minimum design standards were lacking—developers might try to reduce costs, plans were not enforced or concerns about crime led to watering down designs. Sometimes, the impracticality of plans became apparent too late, for example discovering that a football pitch was located on a slope, resulting in its purpose being changed.  Safety auditors often recommended changes to walking and cycling infrastructure because of safety concerns, and developers agreed to these changes to improve their chances of receiving planning permission and to ensure that the LA would take on long-term management of roads. Whilst public health practitioners also considered accident risks, they were more likely to take an holistic view. Finally, some participants were frustrated by schemes where walking and cycling routes were built after all houses were completed, apparently for cost reasons, because people then got ‘into bad habits’ (Greenspaces stakeholder) and therefore were less likely to use them. | Impact of economic focus, viability and costs | - Role of private sector - External influence - Cost and delay |
| 93 | Not enough resources  Most participants were concerned that LA urban planners were under resourced to engage with the right people, learn about best practice and ensure that health was adequately considered. Limited resources for monitoring and evaluation also restricted learning about effectiveness. Some stakeholders wanted to work more with public health, including master-planning developers, to get feedback on designs (in contrast to volume housebuilders whom participants said had no concern for health). However, most LAs in England did not have a public health practitioner dedicated to urban planning. | - Resourcing of local planning authorities - Monitoring and evaluation | - Health consideration - Dedicated posts and capacity |
| 94 | 2014 | Resourcing of local planning authorities  Capability, skills and knowledge |  |
| 94 | Planners had two main roles within the programme: ﬁrstly to assist projects that required planning permission, and secondly to develop existing planning policy (primarily Local Development Frameworks) that considered local population health needs. When discussing the development of planning policy, the principle of “health prooﬁng” was often invoked. This relates to taking appropriate action to mitigate against potential barriers to performing healthy behaviours and the inclusion of health indicators in new and existing built infrastructure developments. For example, in the following quotes, one planner and one key local actor explained the concept of “health-prooﬁng” in relation to promoting general health and physical activity within their Local Development Framework.  Health prooﬁng was enabled through the additional resources afforded by the HT programme (e.g. funding for specialised healthy urban planners) and knowledge transition through professional relationships with health practitioners. The concept of “health prooﬁng”, either implicitly or explicitly, was adopted in ﬁve out of the six towns to describe their local approach to planning's role in public health. This suggests some commonality in the conceptual interpretation of the role of planning within planning-related interventions and policies with the overall pro- gramme. | - Local plan content and plan-making process | - Health consideration - Impact on implementation - Dedicated posts and capacity - Innovative practices - Funding |
| 94 | Towns that speciﬁcally allocated funding towards employing a planner maintained that they were able to develop health initiatives that may have been otherwise overlooked or not been possible. As part of their role, planners became central contacts for other sectors concerned with health, which helped develop partnership working and assisted in acquiring information and evidence to support the inclusion of health within the wider planning agenda. The value of funding planners through the programme was noted by a senior planner and programme developer | Resourcing of local planning authorities | - Funding - Dedicated posts and capacity - Governance - Access to evidence base |
| 94 | It was acknowledged that there was some scepticism among planners and other professionals (including those in public health) about what planning can practically add to public health practice. However, as the quotes below illustrates, by being receptive and open and developing an understanding of what is possible, partnerships could develop that may be beneﬁcial to health.  Indeed the Healthy Towns programme increased the focus on multi-sector partnership working and helped accelerate the creation of joint agendas that may have otherwise taken longer to naturally develop. As the following planners describe, during their time working on the programme, relationships with other departments were developed and strengthened and further assisted by board meetings that attracted stakeholders from a range of policy sectors. | Partnership, collaboration, relationships, engagement | - Governance - Lack of engagement |
| 94 | It was acknowledged among planners that although their profession can inﬂuence public health, at times there is superﬁcial understanding from those outside of the profession about how this can best be achieved. Although planning and public health may have similar objectives in developing and sustaining healthy communities, the approach adopted by each sector is different. One of the main drivers for this can be attributed to different working practices, where planners routinely operate within structures that facilitate long-term planning (ten or more years). In the following extract, one planners discusses how it is now largely recognised that planners and health professionals should work in unison, but how there are still resistance to this concept by some professionals in each sector | Complexity of planning regulatory framework | - Governance - Lack of alignment |
| 95 | Furthermore differences in language and working practices adopted within the health and planning sectors can hinder the commitment to collaborative working. As one programme manager explained, there is a learning curve that must be appreciated to aid understanding of different roles, the barriers that can exist, and how collaborative working can move forward to support healthy communities. | Capability, skills and knowledge | Impact on implementation |
| 95 | Issues arose in one town, whereby local political activity had an inﬂuence on the development of local planning policy. This occurred despite an increased interest in the integration of planning and health within the local authority. As one planner explained, the political uncertainty around future national policy change that could potentially inﬂuence local systems in the future meant local planning policy development stalled | - Impact of politics and politisation in planning - Impact of planning reform processes | - Uncertainty - Cost and delay |
| 95 | The Healthy Towns programme was implemented at a time when changes were taking place within local planning policy. The quotes below illustrate that the additional resource and heightened interest which accompanied the programme provided an opportunity for public health stakeholders and planners to ensure that health was embedded in long term planning strategy documents to a greater degree than it may otherwise not have been. | - Resourcing of local planning authorities - Impact of planning reform processes | External influence |
| 95 | Additionally, two of the towns were being developed as growth points (a government initiative to develop local areas to support enlarged populations), which provided a new opportunity to inﬂuence new housing and community developments that could notably include health actions. The following quote from a planner provides a concrete example of the beneﬁts of aligning policies across public sector agendas to maximise population health outcomes. | Use of (local) planning powers | Health consideration |
| 95 | Though the relationship between healthy communities and sustainable planning has long been recognised (Barton and Tsourou, 2000), within current planning practice the public health potential of planning has yet to be fully realised. The Healthy Towns programme provided a space for planners to become involved in tackling the environmental determinants of obesity. The programme also provided an opportunity to develop and nurture relationships between planners and public health practitioners, which might have otherwise been neglected. These closer working relationships helped to develop a reciprocal appreciation of working cultures and practices within each other's sector and acted as an “open” channel between local public health and planning departments. The Healthy Towns programme thus provided an opportunity to revive the historical link between the planning and public health professions (Northridge et al., 2003); the resulting improvements in collaboration and knowledge translation between the two sectors would additionally strengthen such a link (Chapman, 2010; Corburn, 2010; Dannenberg et al., 2011 | Partnership, collaboration, relationships, engagement | - Innovative practices - Health consideration |
| 95 | The Healthy Towns programme presented an opportunity to accelerate the integration of planning into public health (including obesity prevention). On the whole, planners referred to how their role could impact the overall health agenda, as opposed to focusing directly on obesity. Planners were considered central to the development of local planning policies and built infrastructure developments that could support healthy living | Visioning and outcomes-focused | - Innovative practices - Deliver wider value and benefits |
| 95 | One way planners achieved this was through articulating the concept of the “health prooﬁng” of local planning documents (such as Local Development Frameworks), which involved placing health at the centre of major spatial planning decisions. Enabling the consideration of public health in planning policy is an achievable outcome for planners who wish an increased focus on health, and to inﬂuence public health (Barton, 2009; Corburn, 2010; Carmichael et al., 2013). | - Local plan content and plan-making process - Decision-making process | - Health consideration - Impact on implementation |
| 96 | Health prooﬁng’ was considered a positive and tangible ‘out- come’ by programme stakeholders, largely because of the perceived long term implications for community health. While health prooﬁng was considered a positive outcome that supports sustain- able population health improvement, it is not a ‘health’ outcome per se. Therefore the development of ‘good’ planning policy should be regarded (and valued) as an appropriate outcome in the initial steps on a programme's causal pathway, as opposed to the actual health impact of the policy (Ogilvie et al., 2011). | Visioning and outcomes-focused | Better use of local levers |
| 96 | 4.4. The need for policy alignment  The Healthy Towns programme was implemented at a time when change was taking place within local planning, predominantly the updating of Local Development Frameworks and Health Impact Assessments. This fortuitous timing provided the catalyst for getting health onto the local planning agenda; without this the planning elements of the programme may have been less successful. This indicates a need for a more managed and formal policy alignment between health and town planning which could ensure more integrated working practices between the two sectors. | Impact of planning reform processes | - Health impact assessment - Up to date local plan - External influence (timing) - Lack of alignment |
| 96 | Indeed, there is a need for more guidance and a national policy that supports easier cross-sector joint working and collaboration (Burns and Bond, 2008; Corburn, 2010). | Strength of national policy | Clarity in national policy |
| 96 | Ideally the town planning and public health sectors should be working towards a set of working practices that includes a shared language and interdisciplinary working (Dannenberg et al., 2011). | Partnership, collaboration, relationships, engagement | - |
| 96 | The data suggest that, if planning policy changes coincide with signiﬁcant changes or initiatives taking place within public health, then there are opportunities for closer alignment. The recently released National Planning Practice Guidance is a positive step towards this goal (DCLG, 2014). | Strength of national policy | - External influence - Clarity in national policy |
| 96 | Currently the two sectors work to different agendas, timeframes and frameworks. For example, there are more robust systems in place for the long-term sustainability of planning policies (e.g. Local Plans) that span longer periods of time (15 þ years). Although towns had shown how this could be achieved, more exploration needs to take place into how changes to political and administrative practices could reconnect public health and planning to support aligned working practices (Corburn, 2010). | - Impact of politics and politisation in planning - Complexity of planning regulatory framework | Governance |
| 94 | In particular, the ‘health prooﬁng’ of planning policy demonstrates a positive outcome of what can be achieved when health and planning agendas are aligned. It is, however, important that ‘health prooﬁng does not become the primary outcome of planning policy, but part of a longer term developmental pathway | Local plan content and plan-making process | - |
| 94 | There is a need for health practitioners to better understand the regulatory systems within which planners work (e.g. in relation to the control of fast-food outlets) and in order to have realistic expectations about how planning can impact on community health. This would be assisted through a process of knowledge exchange between the two sectors | Knowledge transfer, good practices  Capability, skills and knowledge | Professional qualification (knowledge) |
| 94 | While this case study supports the inclusion of planning in current government health policy and of health in government planning policy (see NPPF and NPPG), work still needs to be done in terms of developing relationships between sectors and encouraging policy alignment across health and planning. Recent cross- sector public health initiatives provide an opportunity for closer working practices between public health practitioners and those who can inﬂuence the planning process, but timing is crucial in order that ensure agendas in the two sectors are properly aligned | - Partnership, collaboration, relationships, engagement - Complexity of planning regulatory framework | - Lack of alignment (policy) - External influence |
| 97 | 2016 | Resourcing of local planning authorities |  |
| 97 | The review in figures: •139 LPs examined or submitted for examination •86 LPs found sound  •25 LPs withdrawn on the basis of soundness concerns  •31% of LPAs have an up-to-date LP •1 in 15 LPs fail the duty to cooperate •16 plans found sound with a housing requirement below objectively assessed need  •19% amount by which planned supply exceeds household projections in aggregate within up-to-date LPs •21 LPAs most 'at risk' of Government intervention •Figures exclude London as well as single issue/focused review Plans which do not address housing numbers | - Local plan content and plan-making process - Monitoring and evaluation | - Up to date local plan - Soundness |
| 97 | Since the introduction of the NPPF a total of 139 plans have been examined or submitted for examination outside of London (Figure 3)1. Of these, circa two thirds have been found sound, albeit almost a third have been found sound requiring an immediate or early review. | - Impact of planning reform processes - Local plan content and plan-making process | - Up to date local plan - Soundness |
| 97 | Nine of those failed to meet the legal test of the duty to cooperate, with a key contributing factor in most of these relating to cross boundary housing issues | Impact of strategic planning practices | - |
| 97 | Successfully meeting needs?  Although plan progress has been slow, it is clear that the Plan-led system under the NPPF is starting to deliver positive plans which can meet housing needs. As illustrated in Figure 4, based on the 84 adopted plans in place since the NPPF, in total housing requirements have exceeded the relevant 2012-based household projections (the starting point for considering objectively assessed housing needs) by 19%. This provides an overarching barometer that sound plans are likely to set housing targets that will help the country meet its housing needs | Local plan content and plan-making process  Impact of planning reform processes | - |
| 97 | Planned supply increasing through examination  This relative success in meeting needs has, however, come through upwards attrition on housing numbers during the examination process. Many objectors to plans have argued for higher housing to meet objectively assessed needs,  with almost half of plans having to increase their housing targets to be found sound (Figure 5). This debate itself, whilst serving to achieve the important outcome of plans which meet housing needs, has undoubtedly led to many delays in getting plans in place | - Local plan content and plan-making process - Decision-making process | - Planning Inspectorate - Cost and delay |
| 97 | An impending remedy - local plans expert group  The problems with Local Plans have been on the Government’s radar and prompted Ministers to establish the Local Plans Expert Group (LPEG) in September 2015. LPEG gathered evidence from over 140 respondents and produced its report in March 2016. One of NLP’s Senior Directors -  Matthew Spry - was an advisor to LPEG, with a particular focus on the group’s recommendations in respect of housing. LPEG identified a number of key barriers to plan-preparation and was tasked with simplifying the system “with the aim of slashing the amount of time it takes for local authorities to get them in place”. | - Impact of planning reform process - Local plan content and plan-making process | Cost and delay |
| 98 | LPEG’s recommendations should remove some of the main blockages to plan preparation, and an increased volume and pace of plan-making  activity can be expected over the next 18 months. However, local plans will continue to need to reconcile competing priorities, so a boost to the process, whilst importantly solving the existing causes of delay, will shift the focus of activity to addressing real planning conundrums, particularly in high growth, high constraint locations, but working to a tighter timescale. | - Local plan content and plan-making process - Complexity of planning regulatory framework | External influence |
| 98 | Local plan intervention  Government has set out a commitment to take action to get plans in place and ensure plans have up-to-date policies. As part of this Government has set out that where local planning authorities have not produced a Local Plan by ‘early 2017’ then Government will intervene to arrange for a plan to be written | Local plan content and plan-making process | Up to date local plan |
| 98 | Evidencing environmental and development capacity  Para 14 of the NPPF sets out a clear test that objectively assessed housing needs should be met, unless the adverse impacts of doing so would significantly and demonstrably outweigh the benefits or specific policies in the NPPF indicate development should be restricted. Since the introduction of the NPPF, just 16 Local Plans have been found sound where the Inspector explicitly cited that constraints or adverse impacts justified a housing requirement below OAN. Illustrated in Figure 8, these have predominantly been within areas subject to the types of constraints listed within NPPF footnote 9 and furthermore have mostly been in the south of England. NLP’s experience is that the ‘flex’ in assessing OAN has meant some of the debate over housing need has been a proxy for discussions on environmental capacity, but this flex will end if LPEGs ‘stipulated’ approach is accepted. The debate will move to a proper discussion of the capacity for development | Complexity of planning regulatory framework | - Up to date local plan - Planning Inspectorate - Soundness |
| 99 | Looking to a streamlined system, LPEG recommend all Local Plans should be submitted alongside evidence providing a proportionate Assessment of Environmental Capacity. Where LPAs seek to justify a housing requirement below housing need for their own area by reference to capacity constraints, they will need clear and coherent evidence demonstrating that an area could simply not carry the level of development necessary to meet housing needs. This may be through a more exhaustive search for unconstrained land supply, such as in Brighton and Hove, where the Inspector only found the plan sound having directed the Council during suspension of the examination to “leave no stone unturned” in trying to find additional suitable sites. Alternatively it may be through evidence on environmental impacts, landscape sensitivity, habitats regulation assessment or other constraints, brought together to clearly show why development cannot be accommodated. The NPPF sets the bar high in evidencing insurmountable constraints and, even where it is demonstrable, such approaches will need effective cooperation across the housing market area to ensure any unmet needs are met in less constrained areas. | - Impact of strategic planning practices Evidence base | - (Less) focus on process - Access to evidence base - Use of evidence base |
| 99 | Effective cooperation to meet needs The duty-to-cooperate has never been applied as a duty-to-agree. However, it is this very characteristic which is a perceived weakness, with the duty insufficient in and of itself to generate strategic planning across housing market areas to ensure needs are met. As shown by the numbers on cooperation (see box above), numerous authorities have prepared successful joint local plans, but this is still the exception rather than the rule and there are still unmet needs being left unaddressed (see the Sussex Case Study box below). | Impact of strategic planning practices | - |
| 100 | Addressing this, LPEG set out recommendations on how Local Plans and LPAs can ensure a more effective land supply, including allocating reserve sites as a 20% buffer to the housing trajectory and a new approach to five year land supply similar in nature to the Welsh system of Joint Housing Land Availability Studies. This new approach to five year land supply would involve a process for annual monitoring reports to be signed off by an independent examiner, with the result being treated as the definitive calculation for the following year.  Whilst not removing the debate from Local Plans entirely, it should give confidence that Plans can be more effective in maintaining a five year land supply across the plan period but it will necessitate proper engagement in the new process by all interested parties on an annual basis. There will also be a need for land supply to get to grips with the patchwork of mechanisms for ‘allocating’ sites for development, with most LPAs expecting to have a mix of strategic allocations, sites on brownfield registers (benefitting from Permission in Principle), Neighbourhood Plans and, if required, Site Allocations Plans. Housing Implementation Strategies will play a more important role in piecing this all together. | Use of (local) planning powers | Confidence |
| 100 | Resourcing to deliver  Local Authority budgets are being squeezed, with planning and development activities being hit disproportionately. Local Authority planning budgets have more than halved in real terms over the past six years (Figure 9), highlighting the resource challenges planning departments are facing in getting Local Plans adopted. LPEG’s recommendations for streamlined Local Plans, with more concise Plans and a more succinct and focussed evidence base, will help resource constrained planning teams to produce effective Local Plans in a period of budgetary constraints | Resourcing of local planning authorities | - Funding - Type of evidence base |
| 100 | A streamlined evidence base  The implications of LPEG’s recommendations will be a shifting of the debate and focus from need to delivery. Whilst there will still be a need for SHMAs, a streamlined and standardised OAN process mean new evidence will need to focus on if and how those housing needs are met. This means the new evidence typologies for planning for housing needs will include:  Geographies of Housing Need - understanding functional market areas and where unmet needs will manifest itself across and beyond the HMA.  Development Constraints - how much development can an area carry including analysis of NPPF footnote 9 constraints, environmental capacity, landscape constraints and other development constraints.  Infrastructure Capacity - what infrastructure is needed to support growth.  Green Belt Reviews - robustly reviewing green belt boundaries consistent with achieving sustainable development and meeting needs.  SHLAAs - rigorous evidence which leaves no stone unturned in finding suitable sites, including informing brownfield registers.  Spatial Approach - an evidenced strategy for where and why development is proposed justified against the NPPF through the Sustainability Appraisal (SA).  Agreed Strategy for Unmet Needs - an agreed solution/strategy across an HMA to address any unmet needs arising. For example this might be best achieved through a joint or aligned plan or alternatively a memorandum of understanding on cross-boundary housing needs. | - Evidence base - Use of impact assessment | Type of evidence base |
| 97 | Slow progress continues to be the defining characteristic of Local Plan production, with less than a third of LPAs boasting an up-to-date Local Plan adopted since the introduction of the NPPF four years ago. The reasons for such leisurely plan- making are numerous, but it is clear that planning for housing needs is adding to the time spent both producing and examining Local Plans. | Local plan content and plan-making process | - Cost and delay - Up to date local plan - Planning Inspectorate |
| 97 | Our review of Plan making progress, the issues being faced and the likely future changes to the system in a streamlined process has led us to conclude that all parties engaged in achieving sound Local Plans will need to get to grips with a different set of evidential hurdles focussed around:  • evidencing development constraints, including environmental and deliverability constraints and ensuring Green Belt reviews, where applicable, are undertaken more effectively;  • ensuring a more robust and deliverable land supply; and  • evidence on spatial strategy and distributing unmet housing needs as the duty-to- cooperate is given more bite | Evidence base | - Use of evidence base - Type of evidence base - (Less) focus on process |
