## Supplemental_Coding Framework for "Challenges and opportunities in reforming the planning system for England: a rapid review of literature and lessons for the future of planning for health"

**Coding strategy – thematic analysis of eligible articles (n=46) - Initial codes and emerging recurring themes**

| **Key words (challenge/ barrier)** | **Key words (opportunity)** | **Code / labels (initial first level)** | **Code / labels (explained)** | **Code / labels (second level)** |
| --- | --- | --- | --- | --- |
| **System (node)** |  |  |  |  |
| - No national health policy requirement | - Evaluation practice | 1. Strength of national policy | Strength, quality and consistency in national policy, clarity, regulations | 1. Increase regulations/ powers 2. Repeal regulations 3. Clarity in national policy 4. Non-planning regulations 5. Better use of local levers 6. Up to date local plan 7. Impact of public health reforms 8. Deliver wider value and benefits 9. Uncertainty 10. Consistency 11. Risk management 12. Impact on implementation 13. Cost and delay 14. Funding 15. External influence 16. Lack of alignment 17. Lack of engagement 18. Innovative practices 19. Frontloading/ upstream 20. Confidence 21. (Less) focus on process 22. Governance (coordination, institutional) 23. Role of elected members 24. Political commitment 25. Role of private sector 26. Private sector investment 27. Role of local communities 28. Environmental impact assessment 29. Health impact assessment 30. Use of evidence base 31. Relevance of evidence 32. Type of evidence 33. Access to evidence base 34. Soundness 35. Professionalism and impact on profession 36. Planning fees 37. Professional qualification (knowledge) 38. Commercial and development economics skills 39. Salary level 40. Health consideration 41. Externalised expertise 42. Dedicated posts and capacity 43. Planning Inspectorate |
| - Regulatory | - Better use of local planning powers | 2. Impact of planning reform processes | + progress, public health reforms |  |
| - Health in assessment tools such as EIA/ SEA | - Capture public value | 3. Complexity of planning regulatory framework | Competing priorities, lack of alignment between planning and non-planning systems |  |
| - Local plans not addressing issues | - Long term vision in local plans | 4. Impact of strategic planning practices | Abolishing RSSs, duty to cooperate |  |
| - Too focused on system and plans | - Consistency/ certainty | 5. Use of impact assessment |  |  |
| - Complexity of regulatory framework | - Use of design reviews | 6. Use of (local) planning powers |  |  |
| - Inflexibility of regulations | - Speed of decisions | 7. Local plan content and plan-making process |  |  |
| - Removal of National Indicators | - HIA and health in IAs | 8. Decision-making process | Delivery, implementation, PINS |  |
| - PDR housing | - Authority Monitoring Reports | 9. Monitoring and evaluation |  |  |
| - Absence or out of date LPs | - National policy and guidance |  |  |  |
| - Planning conditions | - Simplify process (small sites, SMEs) |  |  |  |
| - Delays in decision-making | - Use of SPDs |  |  |  |
| - Planning reform processes | - Stronger national policy |  |  |  |
| - Duty to cooperate |  |  |  |  |
| **Context (node)** |  |  |  |  |
| - Partnership, collaboration, relationships | - Leadership (public, private, community) | 10. Visioning and outcomes-focused | Experiences, purpose of planning, benefits of planning |  |
| - Politics | - Prioritisation | 11. Leadership |  |  |
| - Economic focus/ viability | - Role of government in policy setting | 12. Evidence base |  |  |
| - Evidence base | - PH moving to LG in 2013 | 13. Partnership, collaboration, relationships, engagement | Accountability, joining up, governance , delivery arrangements, consultation practices |  |
| - Health in decisions/ valuations | - Planning Reform | 14. Impact of politics and politisation in planning | National and local politicisation in planning, politicians, elected members |  |
| - Land control | - Role of developers | 15. Impact of economic focus, viability and costs |  |  |
| - Education and communication | - Professionalisation | 16. Stakeholders and statutory consultees | Stakeholders and statutory consultees, including private sector, users, communities |  |
| - Reinventing the wheel | - Inclusive public consultation |  |  |  |
| - Use of targets | - Planning fees |  |  |  |
| - Multiple objectives | - Monitoring and metrics |  |  |  |
| - Threat of legal challenge | - Central repository |  |  |  |
| - Corporate strategies (SCS) |  |  |  |  |
| **Capability (node)** |  |  |  |  |
| - Local authority capacity | - Training (councillors) | 17. Resourcing of local planning authorities | local planning authorities, morale, resourcing wider professions, funding, activities, shortage |  |
| - Resources and budget (Capital, staff etc) | - Training (future professionals) | 18. Capability, skills and knowledge | + Professional commitment, diversity, funded posts, confidence |  |
| - Narrow understanding of health | - Joint/ dedicated posts | 19. Knowledge transfer, good practices |  |  |
| - Understanding of wider outcomes/ knowledge | - Soft skills and knowledge | 20. Training, learning and development | Education |  |
| - Attitudes | - DPH, public health teams |  |  |  |
| - Awareness of evidence | - Planning Inspectorate |  |  |  |
| - Learning and sharing good practice | - Statutory consultees |  |  |  |
| - Responsibilities | - Positive communication |  |  |  |
