## Supplemental_Search Strategy for "Challenges and opportunities in reforming the planning system for England: a rapid review of literature and lessons for the future of planning for health"

**Supplementary. Search strategy**

A systematic search of Ovid Medline, Embase, Scopus, and Web of Science was conducted in July 2024 using a predefined & validated search strategy. Results which were published between April 2010 and March 2024 which looked at factors which impacted the planning systems within England were eligible for inclusion. Literature searches retrieved 6931 results. After de-duplication and screening, 47 results remained.

**Information Sources**

We conducted electronic searches for eligible results on the following databases: Ovid MEDLINE(R) ALL <1946 to July 22, 2024>, Ovid Embase <1974 to 2024 July 22>, Scopus, OVID Social Policy & Practice <202406>, and Web of Science which were all ran on 21 Jul 2024. Additionally a grey literature search of Google Scholar([www.scholar.google.com/](http://www.scholar.google.com/)), Google ([www.google.com](http://www.google.com)), and TRIP Database Pro ([www.tripdatabase.com](http://www.tripdatabase.com)) identified a further 378 results. Citation searching using Citationchaser identified an additional 306 unique results.

All 6391 records were imported into Endnote Library. Deduplication was done using Dedudclick and Endnote. After deduplication there were 5461 unique results. Following deduplication, all records were imported into Rayyan Systematic Review software. After title and abstract screening – 63 unique records remained, of which 61 were retrieved for eligibility screening.

**Search Strategy**

Full details of these searches are available in supplementary material **X** An initial scoping search was ran on OVID Medline & Scopus (May 25 2024) based on key terms and key papers shared by the principal author. Potential key terms were identified from the titles, abstracts & data records of the scoping search results. Search terms were also identified from the and checked using the PubMed PubReMiner word frequency analysis tool. Given the exploratory nature of the scope a traditional PICO (Population, Intervention, Comparison, Outcome) strategy development was deemed not effective – so the research question was broken down into major themes: English planning system, issues that may impact operation, and health/public health impacts. During the scoping phase, it was decided to remove the health/public health impacts as a theme. During the scoping phase, it was decided to use frequency operators around planning terms due to the ubiquitous and homographic nature of planning and planner within the evidence base. This approach was agreed at peer review stage. The search was validated by testing whether it retrieved the shared key papers and 2 systematic reviews identified during the scoping phase. The papers indexed in OVID Medline were found and the non-Medline indexed papers were found within the Scopus strategy. The OVID Medline search strategy was peer reviewed by a UKHSA Knowledge & Evidence Specialist. Peer review involved proofreading the syntax and spelling used within the search and the overall structure but did not make use of the PRESS Checklist with agreed upon changes being made across all database search strategies. Grey Literature search strategies were based on the key free text terms used in the validated searches. Given the scope was focused on England’s planning system only, a validated UK only search filter was used on OVID Medline & OVID Embase (Ref 1), with translated unvalidated versions used on Web of Science & Scopus. The search date range was limited to 1 Apr 2010 – 31 Mar 2024

Ovid MEDLINE(R) ALL <1946 to July 22, 2024>

1	planning.ti. or planning.ab. /freq=3	67620

2	planner*.ti. or planner*.ab. /freq=3	893

3	"land use".ti. or "land use".ab. /freq=2	9946

4	spatial.ti. or spatial.ab. /freq=3	109443

5	exp social planning/ or city planning/ or environment design/ or urban renewal/ or Planning Techniques/	27992

6	((environment* or city or town or urban or community or neighbo?rhood) adj (design* or plan* or develop*)).tw,kw.	15571

7	or/1-6	221233

8	Local Government/	3981

9	((local or county) adj2 (Government or council or authorit*)).tw,kw.	12452

10	government/ or federal government/ or government agencies/ or state government/ or government programs/	55008

11	(government or parliament or byelaw*).tw,kw.	121210

12	or/8-11	174263

13	(Engl* adj2 (planning or spatial)).tw,kw.	140

14	((national or regional or local) adj2 (planning or spatial) adj2 (policy or plan or guidance or strategy or report or review or system or authority)).tw,kw.	154

15	13 or 14	293

16	7 and 12	6154

17	15 or 16	6403

18	Attitude/ or motivation/ or drive/	136440

19	(Enabling or enabler* or motivator* or motivation or disincentive* or incentive* or encourage* or barrier* or obstacle* or hinder* or demotivator* or perspective* or attitude* or belief* or views or opinion* or influenc* or qualitative or facilitator or drive* or block or challenge).tw.	4860204

20	Change Management/ or knowledge management/ or exp management audit/ or organizational culture/ or exp organizational innovation/ or exp organizational objectives/ or Knowledge/ or Knowledge Discovery/ or Program Evaluation/ or Implementation Science/	163805

21	("cost?effective*" or effective* or efficac* or benefit* or success* or outcome* or evaluation* or impact* or engagement or up?take or opportunity).tw,kw.	9609458

22	(knowledge or (lesson adj2 learn*) or (what adj2 work*)).tw,kw.	991626

23	(delivery or implementation or enforcement).tw,kw.	978063

24	"planning constraint".tw,kw.	18

25	((consult* or stakeholder* or practitioner*) adj3 (view* or opinion* or decision or block* or challeng*)).tw,kw.	9334

26	((local or national or regional or strategic) adj2 (planning or policy or review or report or strategy or consult*)).tw,kw.	25390

27	((spatial or planning or building) adj2 (committee or portal or department or framework* or strategy or system* or board or policy or policies or vision* or authorit* or consult* or review or inquir* or capability or process* or commission* or application* or organi?ation or regulation* or rule*)).tw,kw.	48539

28	("Local Plan" or "Local Plans").tw,kw.	90

29	("development management" or "local development order" or "Article 4").tw,kw.	274

30	(AONB or "Area* of Outstanding Natural Beauty").tw,kw.	9

31	("plan making" or plan-making or "National Planning Policy Framework Local Development Framework" or "neighbourhood plan*" or (development adj1 document*)).tw,kw.	221

32	or/18-31	13299816

33	17 and 32	4916

34	exp Great Britain/	396779

35	(national health service* or nhs*).ti,ab,in.	294857

36	(english not ((published or publication* or translat* or written or language* or speak* or literature or citation*) adj5 english)).ti,ab.	130443

37	(gb or "g.b." or britain* or (british* not "british columbia") or uk or "u.k." or united kingdom* or (england* not "new england") or northern ireland* or northern irish* or scotland* or scottish* or ((wales or "south wales") not "new south wales") or welsh*).ti,ab,jw,in.	2571191

38	(bath or "bath's" or ((birmingham not alabama*) or ("birmingham's" not alabama*) or bradford or "bradford's" or brighton or "brighton's" or bristol or "bristol's" or carlisle* or "carlisle's" or (cambridge not (massachusetts* or boston* or harvard*)) or ("cambridge's" not (massachusetts* or boston* or harvard*)) or (canterbury not zealand*) or ("canterbury's" not zealand*) or chelmsford or "chelmsford's" or chester or "chester's" or chichester or "chichester's" or coventry or "coventry's" or derby or "derby's" or (durham not (carolina* or nc)) or ("durham's" not (carolina* or nc)) or ely or "ely's" or exeter or "exeter's" or gloucester or "gloucester's" or hereford or "hereford's" or hull or "hull's" or lancaster or "lancaster's" or leeds* or leicester or "leicester's" or (lincoln not nebraska*) or ("lincoln's" not nebraska*) or (liverpool not (new south wales* or nsw)) or ("liverpool's" not (new south wales* or nsw)) or ((london not (ontario* or ont or toronto*)) or ("london's" not (ontario* or ont or toronto*)) or manchester or "manchester's" or (newcastle not (new south wales* or nsw)) or ("newcastle's" not (new south wales* or nsw)) or norwich or "norwich's" or nottingham or "nottingham's" or oxford or "oxford's" or peterborough or "peterborough's" or plymouth or "plymouth's" or portsmouth or "portsmouth's" or preston or "preston's" or ripon or "ripon's" or salford or "salford's" or salisbury or "salisbury's" or sheffield or "sheffield's" or southampton or "southampton's" or st albans or stoke or "stoke's" or sunderland or "sunderland's" or truro or "truro's" or wakefield or "wakefield's" or wells or westminster or "westminster's" or winchester or "winchester's" or wolverhampton or "wolverhampton's" or (worcester not (massachusetts* or boston* or harvard*)) or ("worcester's" not (massachusetts* or boston* or harvard*)) or (york not ("new york*" or ny or ontario* or ont or toronto*)) or ("york's" not ("new york*" or ny or ontario* or ont or toronto*))))).ti,ab,in.	1855549

39	(bangor or "bangor's" or cardiff or "cardiff's" or newport or "newport's" or st asaph or "st asaph's" or st davids or swansea or "swansea's").ti,ab,in.	75143

40	(aberdeen or "aberdeen's" or dundee or "dundee's" or edinburgh or "edinburgh's" or glasgow or "glasgow's" or inverness or (perth not australia*) or ("perth's" not australia*) or stirling or "stirling's").ti,ab,in.	272970

41	(armagh or "armagh's" or belfast or "belfast's" or lisburn or "lisburn's" or londonderry or "londonderry's" or derry or "derry's" or newry or "newry's").ti,ab,in.	36309

42	or/34-41	3295991

43	(exp africa/ or exp americas/ or exp antarctic regions/ or exp arctic regions/ or exp asia/ or exp australia/ or exp oceania/) not (exp great britain/ or europe/)	3445712

44	42 not 43	3090452

45	33 and 44	589

47	limit 45 to dt=20100401-20240331	273

Embase <1974 to 2024 July 22>

<https://ovidsp.ovid.com/ovidweb.cgi?T=JS&NEWS=N&PAGE=main&SHAREDSEARCHID=6inkWFmboYXMMVQmz2NrrL09kpNW4rWmh2L5aG9G2GTu6g0li7RtvUt8BtXlAHrtL>

1	planning.ti. or planning.ab. /freq=3	84103

2	planner*.ti. or planner*.ab. /freq=3	1046

3	"land use".ti. or "land use".ab. /freq=2	10488

4	spatial.ti. or spatial.ab. /freq=3	117802

5	exp city planning/ or ("land use"/ and *Planning/) or exp "land use planning"/ or environment design/ or urban renewal/ or Planning Techniques/	443372

6	((environment* or city or town or urban or community or neighbo?rhood) adj (design* or plan* or develop*)).tw,kw.	15833

7	or/1-6	655228

8	((local or county) adj2 (Government or council or authorit*)).tw,kw.	15679

9	*government/ or federal government/ or government agencies/ or state government/ or government programs/	179674

10	(government or parliament or byelaw*).tw,kw.	149019

11	or/8-10	278768

12	(Engl* adj2 (planning or spatial)).tw,kw.	90

13	((national or regional or local) adj2 (planning or spatial) adj2 (policy or plan or guidance or strategy or report or review or system or authority)).tw,kw.	173

14	12 or 13	261

15	7 and 11	28553

16	14 or 15	28772

17	exp *attitude/ or cognitive bias/ or consumer attitude/ or cultural bias/ or employee attitude/ or exp optimism/ or pessimism/ or respect/ or exp risk attitude/ or exp motivation/	579828

18	(Enabling or enabler* or motivator* or motivation or disincentive* or incentive* or encourage* or barrier* or obstacle* or hinder* or demotivator* or perspective* or attitude* or belief* or views or opinion* or influenc* or qualitative or facilitator or drive* or block or challenge).tw.	5953191

19	Change Management/ or knowledge management/ or Knowledge/ or Knowledge Discovery/ or exp organizational culture/ or exp organizational innovation/ or exp organizational objectives/ or Program Evaluation/ or Implementation Science/	989652

20	("cost?effective*" or effective* or efficac* or benefit* or success* or outcome* or evaluation* or impact* or engagement or up?take or opportunity).tw,kw.	12906977

21	(knowledge or (lesson adj2 learn*) or (what adj2 work*)).tw,kw.	1239657

22	(delivery or implementation or enforcement).tw,kw.	1301080

23	"planning constraint".tw,kw.	38

24	((consult* or stakeholder* or practitioner*) adj3 (view* or opinion* or decision or block* or challeng*)).tw,kw.	12160

25	((local or national or regional or strategic) adj2 (planning or policy or review or report or strategy or consult*)).tw,kw.	34444

26	((spatial or planning or building) adj2 (committee or portal or department or framework* or strategy or system* or board or policy or policies or vision* or authorit* or consult* or review or inquir* or capability or process* or commission* or application* or organi?ation or regulation* or rule*)).tw,kw.	63458

27	("Local Plan" or "Local Plans").tw,kw.	112

28	("development management" or "local development order" or "Article 4").tw,kw.	380

29	(AONB or "Area* of Outstanding Natural Beauty").tw,kw.	13

30	("plan making" or plan-making or "National Planning Policy Framework Local Development Framework" or "neighbourhood plan*" or (development adj1 document*)).tw,kw.	313

31	or/17-30	17737599

32	16 and 31	20802

33	exp United Kingdom/	479506

34	("national health service*" or nhs*).ti,ab,in,ad.	505195

35	(english not ((published or publication* or translat* or written or language* or speak* or literature or citation*) adj5 english)).ti,ab.	65749

36	(gb or "g.b." or britain* or (british* not "british columbia") or uk or "u.k." or united kingdom* or (england* not "new england") or northern ireland* or northern irish* or scotland* or scottish* or ((wales or "south wales") not "new south wales") or welsh*).ti,ab,jx,in,ad.	3884631

37	(bath or "bath's" or ((birmingham not alabama*) or ("birmingham's" not alabama*) or bradford or "bradford's" or brighton or "brighton's" or bristol or "bristol's" or carlisle* or "carlisle's" or (cambridge not (massachusetts* or boston* or harvard*)) or ("cambridge's" not (massachusetts* or boston* or harvard*)) or (canterbury not zealand*) or ("canterbury's" not zealand*) or chelmsford or "chelmsford's" or chester or "chester's" or chichester or "chichester's" or coventry or "coventry's" or derby or "derby's" or (durham not (carolina* or nc)) or ("durham's" not (carolina* or nc)) or ely or "ely's" or exeter or "exeter's" or gloucester or "gloucester's" or hereford or "hereford's" or hull or "hull's" or lancaster or "lancaster's" or leeds* or leicester or "leicester's" or (lincoln not nebraska*) or ("lincoln's" not nebraska*) or (liverpool not (new south wales* or nsw)) or ("liverpool's" not (new south wales* or nsw)) or ((london not (ontario* or ont or toronto*)) or ("london's" not (ontario* or ont or toronto*)) or manchester or "manchester's" or (newcastle not (new south wales* or nsw)) or ("newcastle's" not (new south wales* or nsw)) or norwich or "norwich's" or nottingham or "nottingham's" or oxford or "oxford's" or peterborough or "peterborough's" or plymouth or "plymouth's" or portsmouth or "portsmouth's" or preston or "preston's" or ripon or "ripon's" or salford or "salford's" or salisbury or "salisbury's" or sheffield or "sheffield's" or southampton or "southampton's" or st albans or stoke or "stoke's" or sunderland or "sunderland's" or truro or "truro's" or wakefield or "wakefield's" or wells or westminster or "westminster's" or winchester or "winchester's" or wolverhampton or "wolverhampton's" or (worcester not (massachusetts* or boston* or harvard*)) or ("worcester's" not (massachusetts* or boston* or harvard*)) or (york not ("new york*" or ny or ontario* or ont or toronto*)) or ("york's" not ("new york*" or ny or ontario* or ont or toronto*))))).ti,ab,in,ad.	3046574

38	(bangor or "bangor's" or cardiff or "cardiff's" or newport or "newport's" or st asaph or "st asaph's" or st davids or swansea or "swansea's").ti,ab,in,ad.	125534

39	(aberdeen or "aberdeen's" or dundee or "dundee's" or edinburgh or "edinburgh's" or glasgow or "glasgow's" or inverness or (perth not australia*) or ("perth's" not australia*) or stirling or "stirling's").ti,ab,in,ad.	419304

40	(armagh or "armagh's" or belfast or "belfast's" or lisburn or "lisburn's" or londonderry or "londonderry's" or derry or "derry's" or newry or "newry's").ti,ab,in,ad.	58669

41	or/33-40	4751628

42	(exp "arctic and antarctic"/ or exp oceanic regions/ or exp western hemisphere/ or exp africa/ or exp asia/) not (exp united kingdom/ or europe/)	3690075

43	41 not 42	4478131

44	32 and 43	2436

45	limit 44 to dd=20100401-20240331	1068

**Scopus**

( ( TITLE-ABS-KEY ( engl* OR local OR county OR town OR city OR urban OR authority OR government OR "land use" ) W/2 ( spatial OR plann* ) ) OR ( EXACTKEYWORD ( "urban planning" OR "land use" ) ) ) AND ( ( ( AFFIL ( "national health service*" OR nhs ) OR TITLE-ABS-KEY ( "national health service*" OR nhs ) OR TITLE-ABS-KEY ( english AND NOT ( ( published OR publication* OR translat* OR written OR language* OR speak* OR literature OR citation* ) W/5 english ) ) ) ) OR ( AFFIL ( gb OR "g.b." OR britain* OR ( british* AND NOT "british columbia" ) OR uk OR "u.k." OR united AND kingdom* OR ( england* AND NOT "new england" ) OR northern AND ireland* OR northern AND irish* OR scotland* OR scottish* OR ( ( wales OR "south wales" ) AND NOT "new south wales" ) OR welsh* ) OR TITLE-ABS-KEY ( gb OR "g.b." OR britain* OR ( british* AND not "british columbia" ) OR uk OR "u.k." OR united AND kingdom* OR ( england* AND NOT "new england" ) OR northern AND ireland* OR northern AND irish* OR scotland* OR scottish* OR ( ( wales OR "south wales" ) AND NOT "new south wales" ) OR welsh* ) ) OR ( AFFIL ( bath OR "bath&apos;s" ) OR TITLE-ABS-KEY ( bath OR "bath&apos;s" ) ) OR ( AFFIL ( ( birmingham AND NOT alabama* ) OR ( "birmingham&apos;s" AND NOT alabama* ) ) OR TITLE-ABS-KEY ( ( birmingham AND NOT alabama* ) OR ( "birmingham&apos;s" AND NOT alabama* ) ) ) OR ( AFFIL ( bradford OR "bradford&apos;s" OR brighton OR "brighton&apos;s" OR bristol OR "bristol&apos;s" OR carlisle* OR "carlisle&apos;s" ) OR TITLE-ABS-KEY ( bradford OR "bradford&apos;s" OR brighton OR "brighton&apos;s" OR bristol OR "bristol&apos;s" OR carlisle* OR "carlisle&apos;s" ) ) OR ( AFFIL ( cambridge AND NOT ( massachusetts* OR boston* OR harvard* ) ) OR TITLE-ABS-KEY ( cambridge AND NOT ( massachusetts* OR boston* OR harvard* ) ) ) OR ( AFFIL ( "cambridge&apos;s" AND NOT ( massachusetts* OR boston* OR harvard* ) ) OR TITLE-ABS-KEY ( "cambridge&apos;s" AND NOT ( massachusetts* OR boston* OR harvard ) ) ) OR ( AFFIL ( ( canterbury AND NOT zealand* ) OR ( "canterbury&apos;s" AND NOT zealand* ) ) OR TITLE-ABS-KEY ( ( canterbury AND NOT zealand* ) OR ( "canterbury&apos;s" AND NOT zealand* ) ) ) OR ( AFFIL ( chelmsford OR "chelmsford&apos;s" OR chester OR "chester&apos;s" OR chichester OR "chichester&apos;s" OR coventry OR "coventry&apos;s" OR derby OR "derby&apos;s" ) OR TITLE-ABS-KEY ( chelmsford OR "chelmsford&apos;s" OR chester OR "chester&apos;s" OR chichester OR "chichester&apos;s" OR coventry OR "coventry&apos;s" OR derby OR "derby&apos;s" ) ) OR ( AFFIL ( durham AND NOT ( carolina* OR nc ) ) OR TITLE-ABS-KEY ( durham AND NOT ( carolina* OR nc ) ) OR AFFIL ( "durham&apos;s" AND NOT ( carolina* OR nc ) ) OR TITLE-ABS-KEY ( "durham&apos;s" AND NOT ( carolina* OR nc ) ) ) OR ( AFFIL ( ely OR "ely&apos;s" OR exeter OR "exeter&apos;s" OR gloucester OR "gloucester&apos;s" OR hereford OR "hereford&apos;s" OR hull OR "hull&apos;s" OR lancaster OR "lancaster&apos;s" OR leeds* OR leicester OR "leicester&apos;s" ) OR TITLE-ABS-KEY ( ely OR "ely&apos;s" OR exeter OR "exeter&apos;s" OR gloucester OR "gloucester&apos;s" OR hereford OR "hereford&apos;s" OR hull OR "hull&apos;s" OR lancaster OR "lancaster&apos;s" OR leeds* OR leicester OR "leicester&apos;s" ) OR AFFIL ( ( lincoln AND NOT nebraska* ) OR ( "lincoln&apos;s" AND NOT nebraska* ) ) OR TITLE-ABS-KEY ( ( lincoln AND NOT nebraska* ) OR ( "lincoln&apos;s" AND NOT nebraska* ) ) OR AFFIL ( ( liverpool AND NOT ( new AND south AND wales* OR nsw ) ) OR ( "liverpool&apos;s" AND NOT ( new AND south AND wales* OR nsw ) ) ) OR TITLE-ABS-KEY ( ( liverpool AND NOT ( new AND south AND wales* OR nsw ) ) OR ( "liverpool&apos;s" AND NOT ( new AND south AND wales* OR nsw ) ) ) OR AFFIL ( london AND NOT ( ontario* OR ont OR toronto* ) ) OR TITLE-ABS-KEY ( london AND NOT ( ontario* OR ont OR toronto* ) ) OR AFFIL ( "london&apos;s" AND NOT ( ontario* OR ont OR toronto* ) ) OR TITLE-ABS-KEY ( "london&apos;s" AND NOT ( ontario* OR ont OR toronto* ) ) OR AFFIL ( manchester OR "manchester&apos;s" ) OR TITLE-ABS-KEY ( manchester OR "manchester&apos;s" ) OR AFFIL ( ( newcastle AND NOT ( new AND south AND wales* OR nsw ) ) OR ( "newcastle&apos;s" AND NOT ( new AND south AND wales* OR nsw ) ) ) OR TITLE-ABS-KEY ( ( newcastle AND NOT ( new AND south AND wales* OR nsw ) ) OR ( "newcastle&apos;s" AND NOT ( new AND south AND wales* OR nsw ) ) ) OR AFFIL ( norwich OR "norwich&apos;s" OR nottingham OR "nottingham&apos;s" OR oxford OR "oxford&apos;s" OR peterborough OR "peterborough&apos;s" OR plymouth OR "plymouth&apos;s" OR portsmouth OR "portsmouth&apos;s" OR preston OR "preston&apos;s" OR ripon OR "ripon&apos;s" OR salford OR "salford&apos;s" OR salisbury OR "salisbury&apos;s" OR sheffield OR "sheffield&apos;s" OR southampton OR "southampton&apos;s" OR st AND albans OR stoke OR "stoke&apos;s" OR sunderland OR "sunderland&apos;s" OR truro OR "truro&apos;s" OR wakefield OR "wakefield&apos;s" OR wells OR westminster OR "westminster&apos;s" OR winchester OR "winchester&apos;s" OR wolverhampton OR "wolverhampton&apos;s" ) OR TITLE-ABS-KEY ( norwich OR "norwich&apos;s" OR nottingham OR "nottingham&apos;s" OR oxford OR "oxford&apos;s" OR peterborough OR "peterborough&apos;s" OR plymouth OR "plymouth&apos;s" OR portsmouth OR "portsmouth&apos;s" OR preston OR "preston&apos;s" OR ripon OR "ripon&apos;s" OR salford OR "salford&apos;s" OR salisbury OR "salisbury&apos;s" OR sheffield OR "sheffield&apos;s" OR southampton OR "southampton&apos;s" OR st AND albans OR stoke OR "stoke&apos;s" OR sunderland OR "sunderland&apos;s" OR truro OR "truro&apos;s" OR wakefield OR "wakefield&apos;s" OR wells OR westminster OR "westminster&apos;s" OR winchester OR "winchester&apos;s" OR wolverhampton OR "wolverhampton&apos;s" ) OR AFFIL ( worcester AND NOT ( massachusetts* OR boston* OR harvard* ) ) OR TITLE-ABS-KEY ( worcester AND NOT ( massachusetts* OR boston* OR harvard* ) ) OR AFFIL ( ( "worcester&apos;s" AND NOT ( massachusetts* OR boston* OR harvard* ) ) OR ( york AND NOT ( "new york*" OR ny OR ontario* OR ont OR toronto* ) ) OR ( "york&apos;s" AND NOT ( "new york*" OR ny OR ontario* OR ont OR toronto* ) ) ) OR TITLE-ABS-KEY ( ( "worcester&apos;s" AND NOT ( massachusetts* OR boston* OR harvard* ) ) OR ( york AND NOT ( "new york*" OR ny OR ontario* OR ont OR toronto* ) ) OR ( "york&apos;s" AND NOT ( "new york*" OR ny OR ontario* OR ont OR toronto* ) ) ) ) OR ( AFFIL ( bangor OR "bangor&apos;s" OR cardiff OR "cardiff&apos;s" OR newport OR "newport&apos;s" OR st AND asaph OR "st asaph&apos;s" OR st AND davids OR swansea OR "swansea&apos;s" ) OR TITLE-ABS-KEY ( bangor OR "bangor&apos;s" OR cardiff OR "cardiff&apos;s" OR newport OR "newport&apos;s" OR st AND asaph OR "st asaph&apos;s" OR st AND davids OR swansea OR "swansea&apos;s" ) OR AFFIL ( aberdeen OR "aberdeen&apos;s" OR dundee OR "dundee&apos;s" OR edinburgh OR "edinburgh&apos;s" OR glasgow OR "glasgow&apos;s" OR inverness OR ( perth AND NOT australia* ) OR ( "perth&apos;s" AND NOT australia* ) OR stirling OR "stirling&apos;s" ) OR TITLE-ABS-KEY ( aberdeen OR "aberdeen&apos;s" OR dundee OR "dundee&apos;s" OR edinburgh OR "edinburgh&apos;s" OR glasgow OR "glasgow&apos;s" OR inverness OR ( perth AND NOT australia* ) OR ( "perth&apos;s" AND NOT australia* ) OR stirling OR "stirling&apos;s" ) OR AFFIL ( armagh OR "armagh&apos;s" OR belfast OR "belfast&apos;s" OR lisburn OR "lisburn&apos;s" OR londonderry OR "londonderry&apos;s" OR derry OR "derry&apos;s" OR newry OR "newry&apos;s" ) OR TITLE-ABS-KEY ( armagh OR "armagh&apos;s" OR belfast OR "belfast&apos;s" OR lisburn OR "lisburn&apos;s" OR londonderry OR "londonderry&apos;s" OR derry OR "derry&apos;s" OR newry OR "newry&apos;s" ) ) ) AND ( ( ( TITLE-ABS-KEY ( "plan making" OR plan-making OR "National Planning Policy Framework" OR "Local Development Framework" OR "neighbourhood plan*" OR ( development W/1 document* ) ) ) OR ( TITLE-ABS-KEY ( "development management" OR "local development order" OR "Article 4" ) ) OR ( TITLE-ABS-KEY ( aonb OR "Area* of Outstanding Natural Beauty" ) ) OR ( TITLE-ABS-KEY ( "Local Plan" OR "Local Plans" ) ) OR ( TITLE-ABS-KEY ( ( local OR national OR regional OR strategic ) W/1 ( planning OR policy OR review OR report OR strategy OR consult* ) ) ) OR ( TITLE-ABS-KEY ( ( spatial OR planning OR building ) W/2 ( committee OR portal OR department OR framework* OR strategy OR system* OR board OR policy OR policies OR vision* OR authorit* OR consult* OR review OR inquir* OR capability OR process* OR commission* OR application* OR organi?ation OR regulation* OR rule* ) ) ) OR ( TITLE-ABS-KEY ( ( consult* OR stakeholder* OR practitioner* ) W/1 ( view* OR opinion* OR decision OR block* OR challeng* ) ) ) ) AND ( ( TITLE-ABS-KEY ( engl* OR local OR county OR town OR city OR urban OR authority OR government OR "land use" ) W/2 ( spatial OR plann* ) ) OR ( EXACTKEYWORD ( "urban planning" OR "land use" ) ) ) AND PUBYEAR > 2009 AND PUBYEAR < 2026 ) AND PUBYEAR > 2009 AND PUBYEAR < 2025 AND ( LIMIT-TO ( AFFILCOUNTRY , "United Kingdom" ) )

**Web of Science**

Search link: <https://www.webofscience.com/wos/woscc/summary/014b780e-aaaf-449f-a3c9-00b66e0ac0b5-f1f2404c/relevance/1>
