## Supplemental_CASP scores for "Challenges and opportunities in reforming the planning system for England: a rapid review of literature and lessons for the future of planning for health"

**Supplementary. Summary of CASP Qualitative Appraisal Checklist scores**

| **Reference** | **Q1** | **Q2** | **Q3** | **Q4** | **Q5** | **Q6** | **Q7** | **Q8** | **Q9** | **Total** |
| --- | --- | --- | --- | --- | --- | --- | --- | --- | --- | --- |
| (Adams et al., 2016) | 1 | 2 | 1 | 0 | 0 | 0 | 0 | 0 | 2 | 6 |
| (Airey and Doughty, 2020) | 2 | 0 | 0 | 0 | 1 | 1 | 0 | 0 | 0 | 4 |
| (All Party Parliamentary Group for SME House Builders, 2020) | 2 | 2 | 2 | 2 | 2 | 2 | 1 | 1 | 2 | 16 |
| (Bates et al., 2023) | 2 | 2 | 2 | 1 | 2 | 0 | 1 | 2 | 2 | 14 |
| (Black et al., 2021) | 2 | 2 | 2 | 2 | 2 | 2 | 2 | 2 | 2 | 18 |
| (Bristow, 2021) | 2 | 2 | 2 | 1 | 2 | 1 | 2 | 2 | 2 | 16 |
| (Callway et al., 2023) | 2 | 2 | 2 | 2 | 2 | 2 | 1 | 2 | 2 | 17 |
| (Carmichael et al., 2012) | 2 | 2 | 2 | 1 | 2 | 0 | 1 | 2 | 2 | 14 |
| (Carmichael et al., 2013) | 2 | 2 | 2 | 1 | 2 | 0 | 0 | 2 | 2 | 13 |
| (Carmichael et al., 2019) | 2 | 2 | 2 | 1 | 2 | 0 | 0 | 1 | 2 | 12 |
| (Clifford, 2016) | 2 | 2 | 2 | 1 | 2 | 0 | 0 | 1 | 2 | 12 |
| (Clifford, 2022) | 2 | 2 | 2 | 1 | 1 | 0 | 0 | 0 | 0 | 8 |
| (Comptroller and Auditor General, 2019) | 2 | 1 | 2 | 1 | 1 | 2 | 1 | 1 | 2 | 13 |
| (Denning-Johnson and Coleman, 2016) | 1 | 1 | 1 | 2 | 1 | 0 | 0 | 1 | 2 | 9 |
| (Design Council and Social Change UK, 2018) | 2 | 2 | 2 | 2 | 2 | 0 | 0 | 2 | 2 | 14 |
| (Dodd et al., 2023) | 2 | 2 | 2 | 2 | 2 | 0 | 0 | 1 | 2 | 13 |
| (Henderson et al., 2015) | 2 | 2 | 2 | 1 | 2 | 0 | 0 | 1 | 2 | 12 |
| (HBF et al., 2020) | 2 | 2 | 2 | 1 | 1 | 1 | 0 | 2 | 2 | 13 |
| (HBF et al., 2021) | 2 | 2 | 2 | 1 | 1 | 1 | 0 | 2 | 2 | 13 |
| (HBF et al., 2022) | 2 | 2 | 2 | 1 | 1 | 1 | 0 | 2 | 2 | 13 |
| (HBF et al., 2024) | 2 | 2 | 2 | 1 | 1 | 1 | 0 | 2 | 2 | 13 |
| (Committee of Public Accounts, 2019) | 1 | 0 | 0 | 1 | 1 | 0 | 0 | 1 | 1 | 5 |
| (Housing Communities and Local Government Committee, 2021) | 2 | 2 | 1 | 1 | 1 | 0 | 0 | 1 | 1 | 9 |
| (Levelling Up Housing and Communities Committee, 2023) | 1 | 1 | 1 | 2 | 1 | 0 | 0 | 0 | 1 | 7 |
| (Ige-Elegbede et al., 2021) | 2 | 2 | 2 | 2 | 2 | 2 | 2 | 2 | 2 | 18 |
| (Keeble et al., 2020) | 2 | 2 | 2 | 2 | 2 | 2 | 2 | 2 | 2 | 18 |
| (Lake et al., 2017) | 2 | 2 | 2 | 2 | 2 | 0 | 2 | 2 | 2 | 16 |
| (Le Gouais et al., 2020) | 2 | 2 | 1 | 2 | 2 | 0 | 2 | 2 | 2 | 15 |
| (Le Gouais et al., 2023) | 2 | 2 | 2 | 2 | 2 | 2 | 2 | 2 | 2 | 18 |
| (Lichfields, 2017) | 2 | 2 | 1 | 2 | 2 | 2 | 0 | 1 | 1 | 13 |
| (Lichfields, 2023) | 2 | 1 | 1 | 1 | 1 | 0 | 0 | 1 | 2 | 9 |
| (Local Plans Expert Group, 2016) | 1 | 1 | 0 | 0 | 1 | 0 | 0 | 1 | 0 | 4 |
| (Maidment-Blundell, 2020) | 2 | 2 | 2 | 2 | 2 | 0 | 0 | 2 | 2 | 14 |
| (May Goodwin et al., 2014) | 2 | 2 | 2 | 2 | 2 | 0 | 2 | 2 | 2 | 16 |
| (Murtagh et al., 2019) | 2 | 2 | 2 | 1 | 2 | 1 | 2 | 2 | 2 | 16 |
| (Murtagh et al., 2020) | 2 | 2 | 2 | 2 | 2 | 0 | 0 | 2 | 2 | 14 |
| (Nathaniel Lichfield & Partners, 2016) | 2 | 1 | 2 | 2 | 1 | 0 | 0 | 2 | 2 | 12 |
| (O’Malley et al., 2023) | 2 | 2 | 2 | 2 | 2 | 1 | 2 | 2 | 2 | 17 |
| (Oliver and de Vocht, 2015) | 2 | 2 | 2 | 2 | 2 | 0 | 0 | 1 | 2 | 13 |
| (Ricci et al., 2021) | 2 | 2 | 2 | 2 | 2 | 2 | 2 | 2 | 2 | 18 |
| (Riley and de Nazelle, 2018) | 2 | 2 | 0 | 0 | 0 | 0 | 0 | 0 | 0 | 4 |
| (Royal Town Planning Institute, 2020) | 2 | 1 | 2 | 1 | 2 | 0 | 0 | 1 | 2 | 11 |
| (Smith, 2018) | 2 | 2 | 2 | 2 | 2 | 1 | 1 | 2 | 2 | 16 |
| (Strategic Solutions, 2010) | 2 | 2 | 2 | 1 | 2 | 0 | 0 | 2 | 2 | 13 |
| (Raynsford Review, 2018) | 0 | 1 | 0 | 0 | 1 | 0 | 0 | 1 | 1 | 4 |
| (Young and Essex, 2020) | 2 | 2 | 2 | 2 | 2 | 1 | 1 | 1 | 1 | 14 |
